# Identification of potential core genes in hepatoblastoma via bioinformatics analysis

**DOI:** 10.1101/2020.12.22.20248756

**Authors:** Basavaraj Vastrad, Chanabasayya Vastrad, Iranna Kotturshetti

## Abstract

Hepatoblastoma is the childhood liver cancer. Profound efforts have been made to illuminate the pathology, but the molecular mechanisms of hepatoblastoma are still not well understood. To identify the candidate genes in the carcinogenesis and progression of hepatoblastoma, microarray dataset GSE131329 was downloaded from Gene Expression Omnibus (GEO) database. The differentially expressed genes (DEGs) were identified, and pathway and Gene Ontology (GO) enrichment analysis were performed. The protein-protein interaction network (PPI), module analysis, target gene - miRNA regulatory network and target gene - TF regulatory network were constructed and analyzed. A total of 996 DEGs were identified, consisting of 499 up regulated genes and 497 down regulated genes. The pathway and Gene Ontology (GO) enrichment analysis of the DEGs include proline biosynthesis, superpathway of tryptophan utilization, chromosome organization and organic acid metabolic process. Twenty-four hub genes were identified and biological process analysis revealed that these genes were mainly enriched in cell cycle, chromosome organization, lipid metabolic process and oxidation-reduction process. Validation of hub genes showed that TP53, PLK1, AURKA, CDK1, ANLN, ESR1, FGB, ACAT1, GOT1 and ALAS1 may be involved in the carcinogenesis, invasion or recurrence of hepatoblastoma. In conclusion, DEGs and hub genes identified in the present study help us understand the molecular mechanisms underlying the carcinogenesis and progression of hepatoblastoma, and provide candidate targets for diagnosis and treatment of hepatoblastoma.

## Introduction

Hepatoblastoma is a highly complex and uncommon malignant type of liver cancer appears in infants and children [Kremer et al 2014], and accounting for just over 1% of pediatric cancers [Spector and Birch, 2012]. Despite significant advancements in treatments, including surgery [Busweiler et al 2017], chemotherapeutic [Hiyama et al 2016] and liver transplantation [Cruz Jr et al 2013], the survival rate of patients with hepatoblastoma has not sufficiently improved. The high rates of recurrence and metastasis in patients with hepatoblastoma require the critical advancement of novel diagnostic strategies and therapeutic agents to enhance patient prognosis. As the molecular mechanisms of hepatoblastoma tumorigenesis and development are not yet fully understood, there remains to be a number of unsolved issues in the diagnosis and treatment of hepatoblastoma. Therefore, it is important to identify new biomarkers and pathways linked with tumorigenesis and patient prognosis, in order to help resolve the suitable basic molecular mechanisms, and to help discover novel diagnostic and prognostic markers, and therapeutic targets.

Altered expression and mutations of genes, and signalling pathways are associated in the advancement and development of hepatoblastoma. Genes such as β-catenin [Koch et al. 1999], Axin [Miao et al. 2003], SOCS-1 [Nagai et al. 2003], lipin 1 [Ishimoto et al. 2009] and MT1G [Sakamoto et al. 2010] were linked with development of hepatoblastoma. Signalling pathways such as PI3K/Akt, ERK and p38 signaling pathways [Cui et al. 2016], MAPK signaling pathways [Yuan et al. 2013], notch signalling pathway [Aktaş et al. 2010], Wnt/β□catenin signalling pathway [Cui et al. 2020] and EGFR-ASAP1 signaling pathway [Ranganathan et al. 2016] were responsible for progression hepatoblastoma. Hepatoblastoma remains a require disease to treat, and more investigations are required to develop the understanding of the underlying molecular mechanisms to diagnose genes and signaling for the progression of new therapies. Therefore, the pathogenesis of hepatoblastoma warrants further studies.

With the accelerated advancement of microarray technology, some high throughput platforms for analysis of gene expression are extensively used to examine the differentially expressed genes (DEGs) during cancer progression [Ma et al. 2019]. Many gene expression profiling studies on hepatoblastoma have been implemented using microarray technology and identified many up and down regulated genes linked with progression of hepatoblastoma [Liu et al. 2018; Adesina et al. 2009].

Therefore, the purpose of the current investigations was to download and analyze expression profiling dataset of human samples from the Gene Expression Omnibus (GEO) database, and to identify DEGs between hepatoblastoma tissues samples and noncancerous liver tissue samples. Subsequently, pathway and Gene Ontology (GO) enrichment analysis were carried out. Protein□protein interaction (PPI) network analysis, module analysis, target gene - miRNA regulatory network and target gene - TF regulatory network were used to establish the molecular pathogenesis underlying carcinogenesis and progression of hepatoblastoma. Finally, the hub genes were further verified based on UALCAN, cBio Portal, The Human Protein Atlas, receiver operator characteristic (ROC) curve analysis, RT-PCR analysis and immune infiltration analysis. The results may provide new diagnostic and prognostic biomarkers, and therapeutic target molecules in hepatoblastoma metastasis.

## Materials and methods

### Access to public data

The GEO (http://www.ncbi.nlm.nih.gov/geo) is a wide platform for accumulate genomic or microarray data [Edgar et al. 2002]. Expression profiling datasets [GSE131329] were obtained from GEO. The GSE131329 dataset was tested based on the GPL6244 platform [HuGene-1_0-st] Affymetrix Human Gene 1.0 ST Array [transcript (gene) version], including 53 hepatoblastoma tissues samples and 14 noncancerous liver tissue samples.

### Data pre-processing

The GSE131329 gene expression profiles were preprocessed using affyPLM software (https://www.bioconductor.org/packages/release/bioc/html/affyPLM.html) in the Release (3.10) package [Heber and Sick, 2006]. The original microarray data were transformed into gene symbols according to annotation information of the array platform. If certain probes coincided with the same gene, the moderate scores were determined as the gene expression value of these probes. Robust multiarray average (RMA) in R affy package [Gautier et al. 2004] was used to normalize the matrix.

### DEGs analysis

The DEGs in hepatoblastoma samples compared with noncancerous liver samples were diagnosed using the Limma package (https://www.bioconductor.org/packages/release/bioc/html/limma.html) [Ritchie et al. 2015] in R Bioconductor. Finally, |log foldchange (FC)| > 1.07 for up regulated genes; |log foldchange (FC)| < -1.25 for down regulated genes and PLvalue <0.05 were used as the threshold values for considering the DEGs.

### Pathway enrichment analysis of DEGs

BIOCYC (https://biocyc.org/) [Caspi et al. 2016], Kyoto Encyclopedia of Genes and Genomes (KEGG) (http://www.genome.jp/kegg/pathway.html) [Kanehisa et al. 2018], Pathway Interaction Database (PID) (https://wiki.nci.nih.gov/pages/viewpage.action?pageId=315491760) [Schaefer et al. 2009], REACTOME (https://reactome.org/) [Fabregat et al. 2018], GenMAPP (http://www.genmapp.org/) [Dahlquist et al. 2002], MSigDB C2 BIOCARTA (http://software.broadinstitute.org/gsea/msigdb/collections.jsp) [Subramanian et al. 2005], PantherDB (http://www.pantherdb.org/) [Mi et al. 2017], Pathway Ontology (http://www.obofoundry.org/ontology/pw.html) [Petri et al. 2017] and Small Molecule Pathway Database (SMPDB) (http://smpdb.ca/) [Jewison et al. 2014] are a bioinformatics resource that links genomes to metabolic pathways. Pathway enrichment analysis for DEGs (up and down regulated genes) was performed by using ToppGene (ToppFun) (https://toppgene.cchmc.org/enrichment.jsp) [Chen et al 2009]. P-value < 0.05 was chosen as the cut-off criteria for significant pathway enrichment analysis.

### GO enrichment analysis of DEGs

ToppGene (ToppFun) (https://toppgene.cchmc.org/enrichment.jsp) [Chen et al 2009] was used to perform Gene Ontology (GO) (http://www.geneontology.org/) [Lewis, 2017] enrichment analyses. P-value < 0.05 was chosen as the cut-off criteria for significant GO enrichment analysis.

### Construction of the PPI network and module analysis

The PPI network was constructed based on all the DEGs (up and down regulated genes) using IID (Integrated Interactions Database) from a well known online server (http://iid.ophid.utoronto.ca) [Kotlyar et al 2019], which integrates different PPI databases such as Biological General Repository for Interaction Datasets (BioGRID) (https://thebiogrid.org/) [Oughtred et al 2019], IntAct Molecular Interaction Database (https://www.ebi.ac.uk/intact/) [Orchard et al 2014], the Molecular INTeraction database (MINT) (https://mint.bio.uniroma2.it/) [Licata et al 2012], InnateDB (https://www.innatedb.com/) [Breuer et al 2013], Database of Interacting Proteins (DIP) (http://dip.doe-mbi.ucla.edu/dip/Main.cgi) [Salwinski et al 2004], Human Protein Reference Database (HPRD) (http://www.hprd.org/) [Keshava Prasad et al 2009] and Biomolecular Interaction Network Database (BIND) (http://bond.unleashedinformatics.com/) [Willis and Hogue, 2006]. A united score of >0.4 was determine as the threshold value for constructing the PPI network. The PPI network was visualized using Cytoscape software (version 3.7.2, http://cytoscape.org/) [Shannon et al 2003]. Five topological characteristics of the genes in the PPI network, including node degree [Przulj et al 2004], betweenness centrality [Nguyen et al 2011], stress centrality [Shi and Zhang, 2011], closeness centrality [Li et al 2018] and clustering coefficient [Wang et al 2012], were calculated using the Network Analyzer plugin in Cytoscape software (http://apps.cytoscape.org/apps/NetworkAnalyzer). In addition, PEWCC1 (http://apps.cytoscape.org/apps/PEWCC1) in Cytoscape software was used to analyze the most significant module, with the threshold value of 5 [Zaki et al 2013].

### Construction of target genes - miRNA regulatory network

The miRNAs (microRNA) that can regulate the up and down regulated genes were predicted using the miRNet database (https://www.mirnet.ca/) [Fan and Xia, 2018] with default significant parameters. Only the interaction relationships can be predicted by ten common algorithms, including TarBase (http://diana.imis.athena-innovation.gr/DianaTools/index.php?r=tarbase/index) [Vlachos et al. 2015], miRTarBase (http://mirtarbase.mbc.nctu.edu.tw/php/download.php) [Chou et al. 2018], miRecords (http://miRecords.umn.edu/miRecords) [Xiao et al. 2009], miR2Disease (http://www.mir2disease.org/) [Jiang et al. 2009], HMDD (http://www.cuilab.cn/hmdd) [Huang et al. 2019], PhenomiR (http://mips.helmholtz-muenchen.de/phenomir/) [Ruepp et al. 2010], SM2miR (http://bioinfo.hrbmu.edu.cn/SM2miR/) [Liu et al. 2013], PharmacomiR (http://www.pharmaco-mir.org/) [Rukov et al. 2014], EpimiR (http://bioinfo.hrbmu.edu.cn/EpimiR/) [Dai et al. 2014] and starBase (http://starbase.sysu.edu.cn/) [Li et al. 2014], were included to construct the target gene- miRNA regulatory network using the Cytoscape software.

### Construction of target genes - TF regulatory network

The TFs (transcription factors) that can regulate the up and down regulated genes were predicted using the NetworkAnalyst database (https://www.networkanalyst.ca/) [Zhou et al 2019] with default significant parameters. Only the interaction relationships can be predicted by ENCODE (http://cistrome.org/BETA/) [Wang et al 2013],were included to construct the target gene-TF regulatory network using the Cytoscape software.

### Validation of hub genes

UALCAN (http://ualcan.path.uab.edu/analysis.html) [Chandrashekar et al 2017] is a website that offers an online validation of survival biomarkersand analyzes the overall survival (OS) of patients with high and low expression of certain genes. In our investigation, up and down regulated hub genes were detected, and a survival curve was drawn. The log-rank p-value was calculated. And the UALCAN [Chandrashekar et al 2017] platform was applied to further verify the expression level of up and down regulated hub genes between hepatoblastoma and normal samples from The Cancer Genome Atlas (TCGA) portal. Next, UALCAN [Chandrashekar et al 2017] platform was applied to further verify the expression level of up and down regulated hub genes from normal to all stages of hepatoblastoma from The Cancer Genome Atlas (TCGA) portal. Mutaion analysis of up and down regulated hub gens was performed by cBioPortal online platform (http://www.cbioportal.org) [Gao et al 2013]. We also evaluated the protein expression of up and down regulated hub genes by using the human protein atlas (HPA, www.proteinatlas.org) [Uhlen et al 2010] database considering that gene expression was not always consistent with its protein level. Receiver operating characteristic (ROC) analysis was conducted by using the generalized linear model (GLM) in machine learning algorithms [Robinet al. 2011] to show the potential diagnostic and prognostic value of up and down regulated hub genes. P<0.05 was considered to indicate a statistically significant difference. Up and down regulated hub genes was quantified using real-time PCR. Total RNA was isolated from cultured Hep G2 and normal liver cells using a TRI Reagent® (Sigma, USA). RNA was then reverse transcribed into cDNA according to the instructions of a FastQuant RT kit (with gDNase; Tiangen Biotech Co., Ltd.). RT-PCR was performed using a QuantStudio 7 Flex real-time PCR system (Thermo Fisher Scientific, Waltham, MA, USA). The reaction conditions were as follows: predenaturation at 95 °C for 10 min and 40 cycles of denaturation at 95 °C for 10 sec, annealing at 60 °C for 20 sec and extension at 72 °C for 34 sec. β-actin was used as the internal reference for up and down regulated hub genes. The primers for up and down regulated hub genes are listed in Table 1. The 2^-ΔΔ^ technique was engaged to measure the ratio of the relative expression of a target gene in the experimental group to that in the control group with the following formulas: ΔΔCt = ΔCt_experimental group_ - ΔCt_control group_ and ΔCt_target gene_ - ΔCt_internal reference_. Ct was the amplification cycle. Three separate experiments were conducted [Livak and Schmittgen, 2001]. The immune infiltration analysis for up and down regulated hub genes have been conducted with the scatter plot in TIMER (https://cistrome.shinyapps.io/timer/) [Li et al. 2017] is a RNA-Seq expression profiling database from The Cancer Genome Atlas (TCGA) portal. Immune infiltration analysis was used to check the immune infiltrates (B cells, CD4+ T cells, CD8+ T cells, neutrophils, macrophages, and dendritic cells) across hepatoblastoma.

**Table 1.**
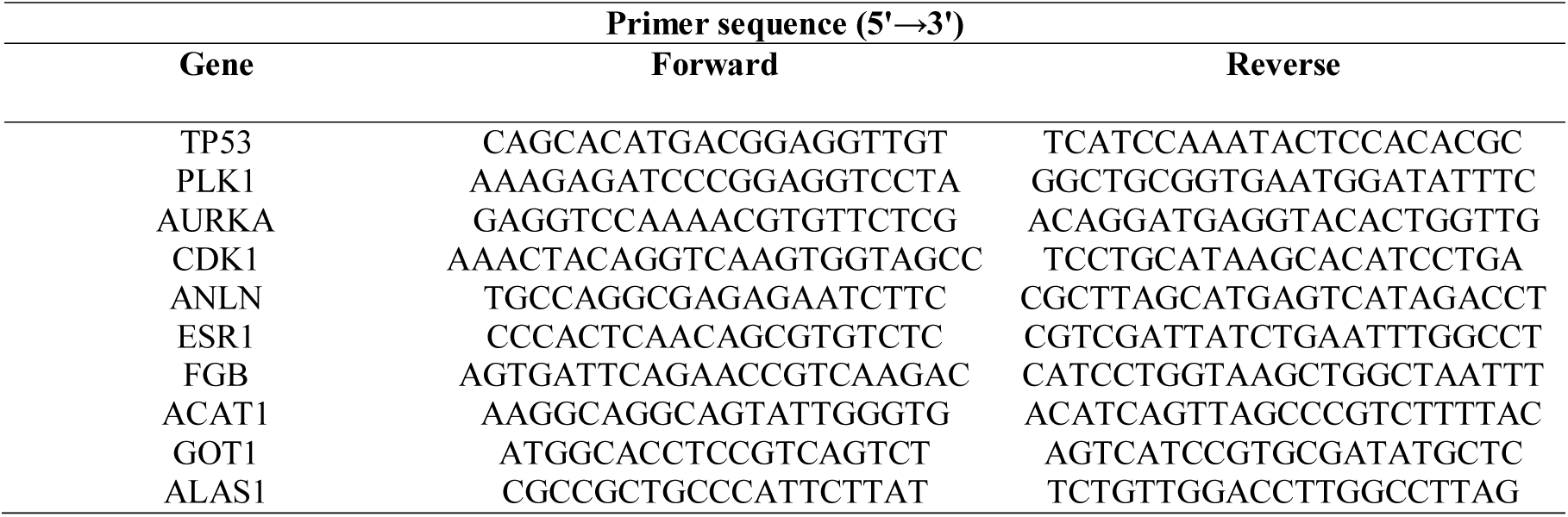
Primers used for quantitative PCR

## Results

### Data pre-processing

Gene expression profile (GSE131329) was collected from the GEO database. A total of 67 samples included 53 hepatoblastoma tissues samples and 14 noncancerous liver tissue samples. The dataset was then performed normalization and batch effect correction (Fig. 1A and 1B).

**Fig. 1.**
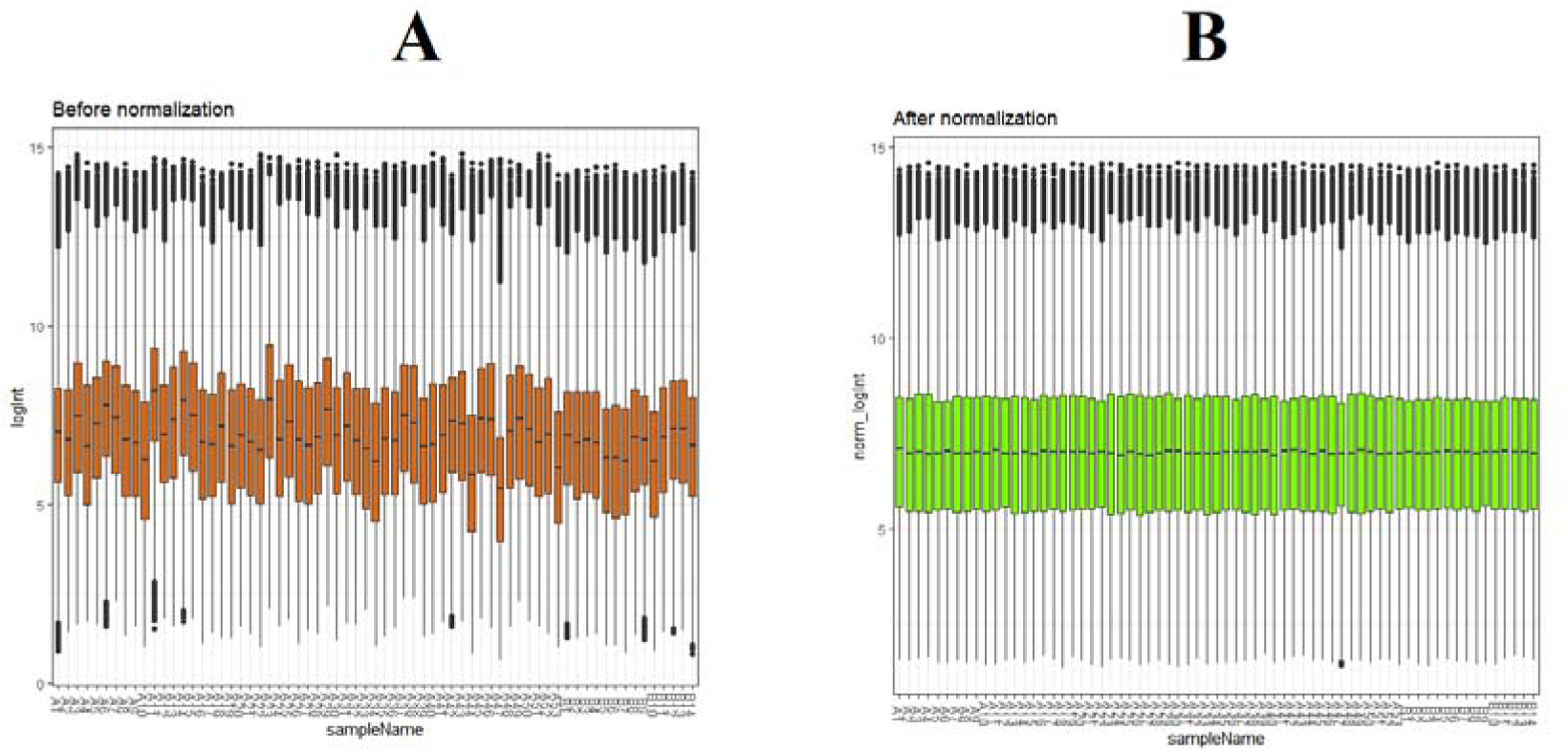
Box plots of the gene expression data before normalization (A) and after normalization (B). Horizontal axis represents the sample symbol and the vertical axis represents the gene expression values. The black line in the box plot represents the median value of gene expression. (A1 – A53 = hepatoblastoma tissues samples; B1 – B14 = noncancerous liver tissue samples)

### DEGs analysis

The series from each chip was analyzed separately using limma in R software, and finally, the DEGs, using adjusted P value < 0.05 and log foldchange (FC)| > 1.07 for up regulated genes; |log foldchange (FC)| < -1.25 for down regulated genes as the cut-off criteria, were identified. A total of 996 DEGs, including 499 up regulated genes and 497 down regulated genes, were detected in hepatoblastoma are listed in Table 2. The heatmap of the up and down regulated hub genes are shown in Fig. 2 and Fig. 3. The volcano plot of gene expression profile data was shown in Fig. 4.

**Fig. 2.**
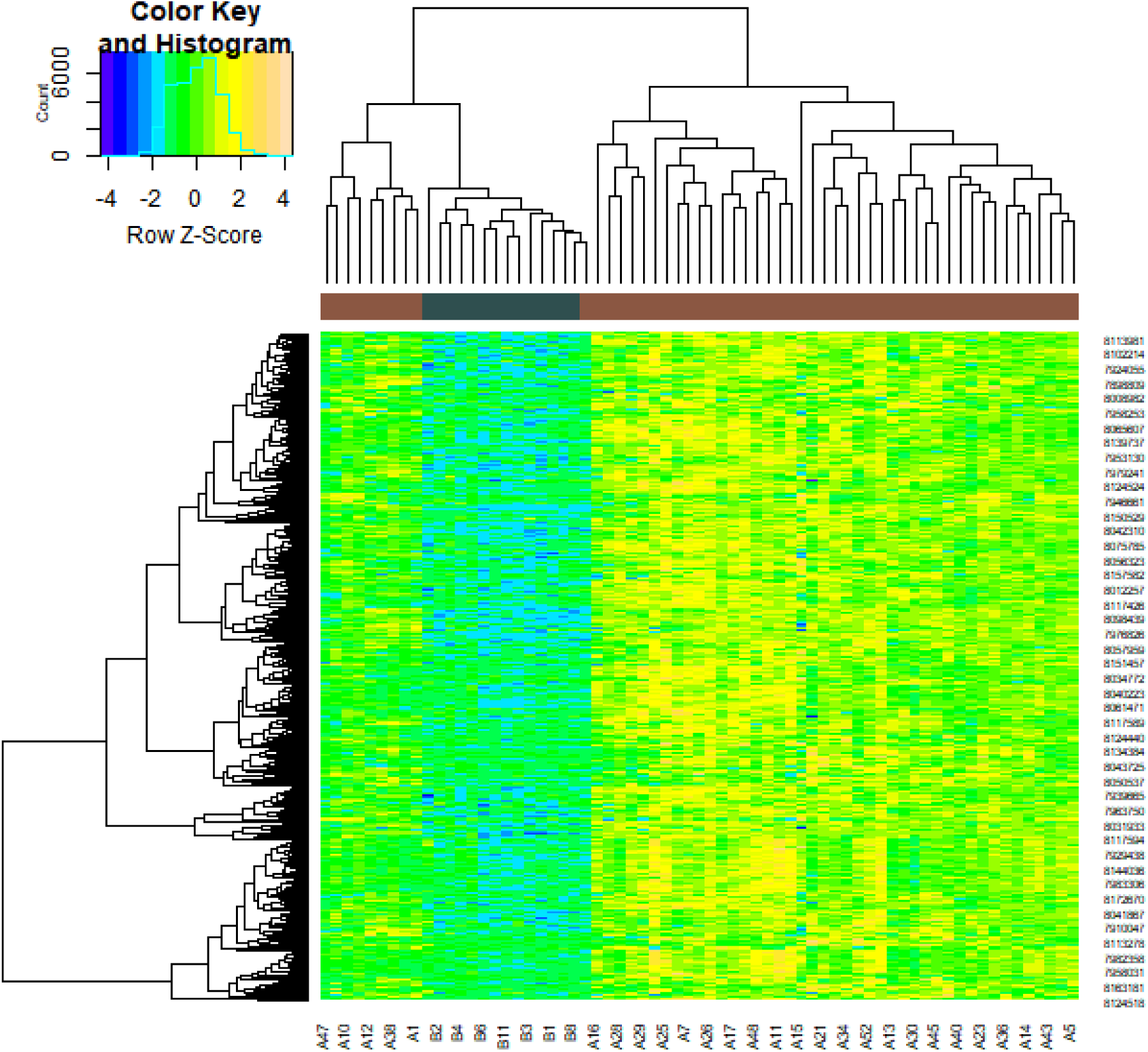
Heat map of up regulated differentially expressed genes. Legend on the top left indicate log fold change of genes. (A1 – A53 = hepatoblastoma tissues samples; B1 – B14 = noncancerous liver tissue samples)

**Fig. 3.**
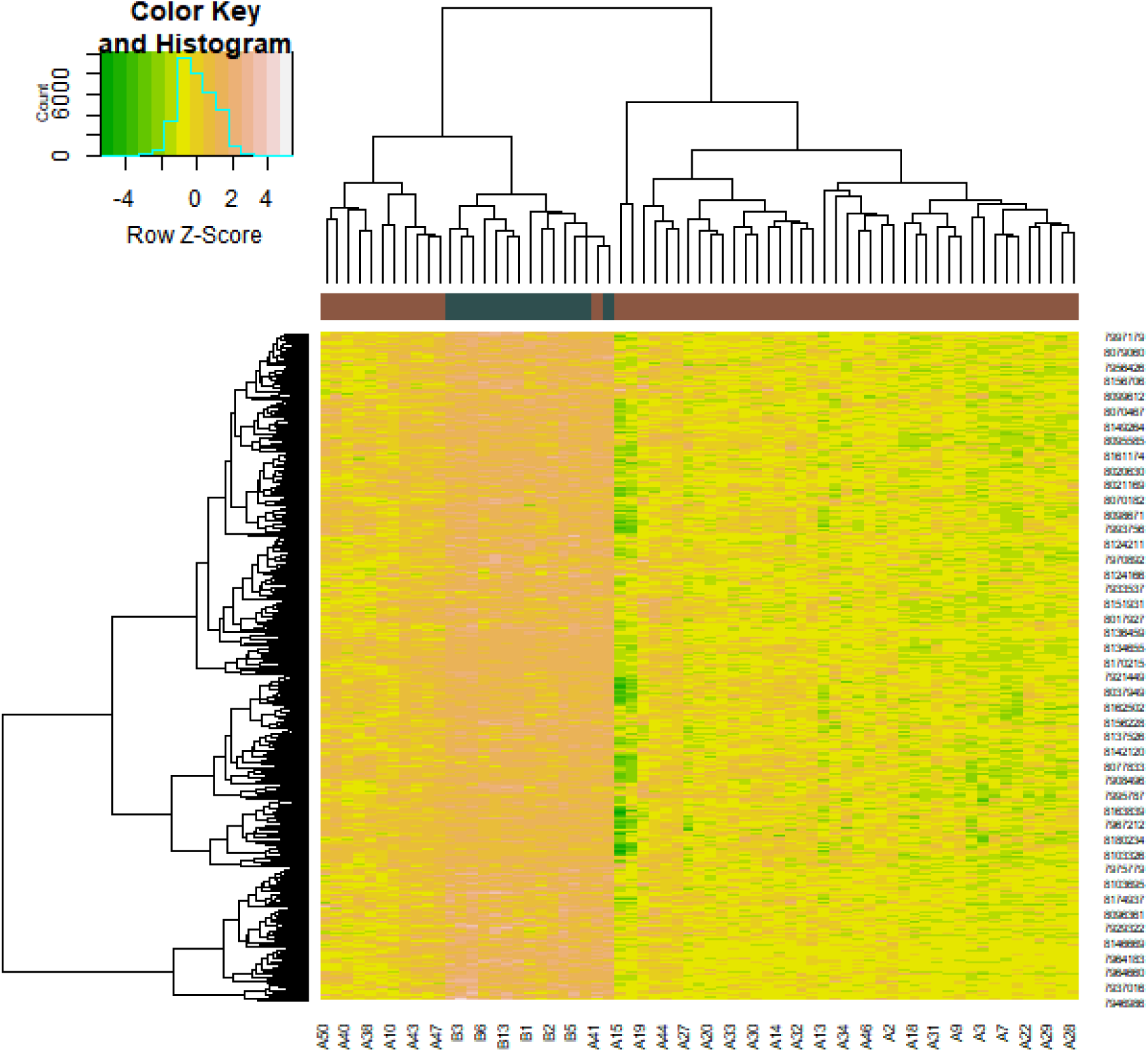
Heat map of down regulated differentially expressed genes. Legend on the top left indicate log fold change of genes. (A1 – A53 = hepatoblastoma tissues samples; B1 – B14 = noncancerous liver tissue samples)

**Fig. 4.**
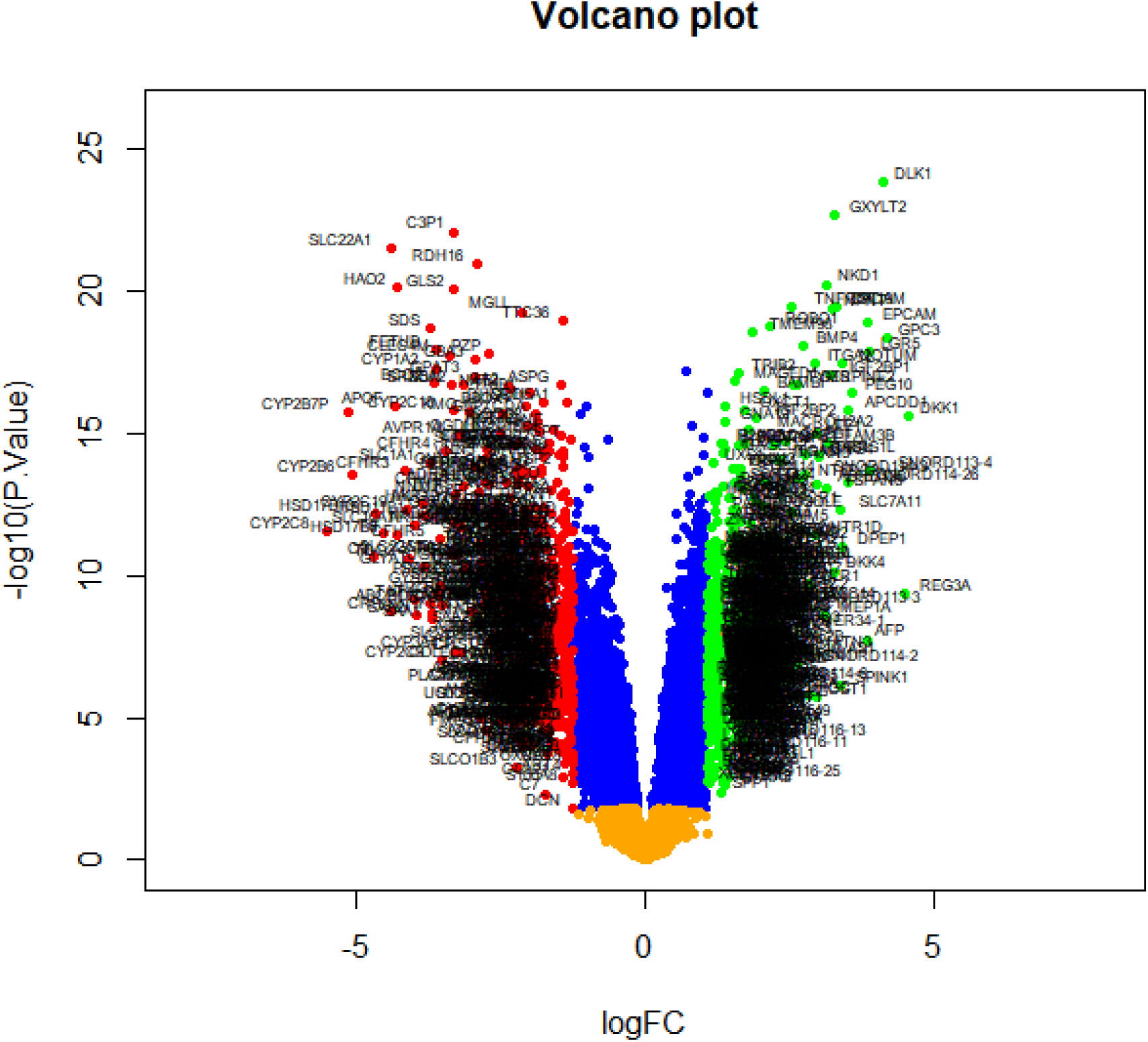
Volcano plot of differentially expressed genes. Genes with a significant change of more than two-fold were selected.

**Table 2.**
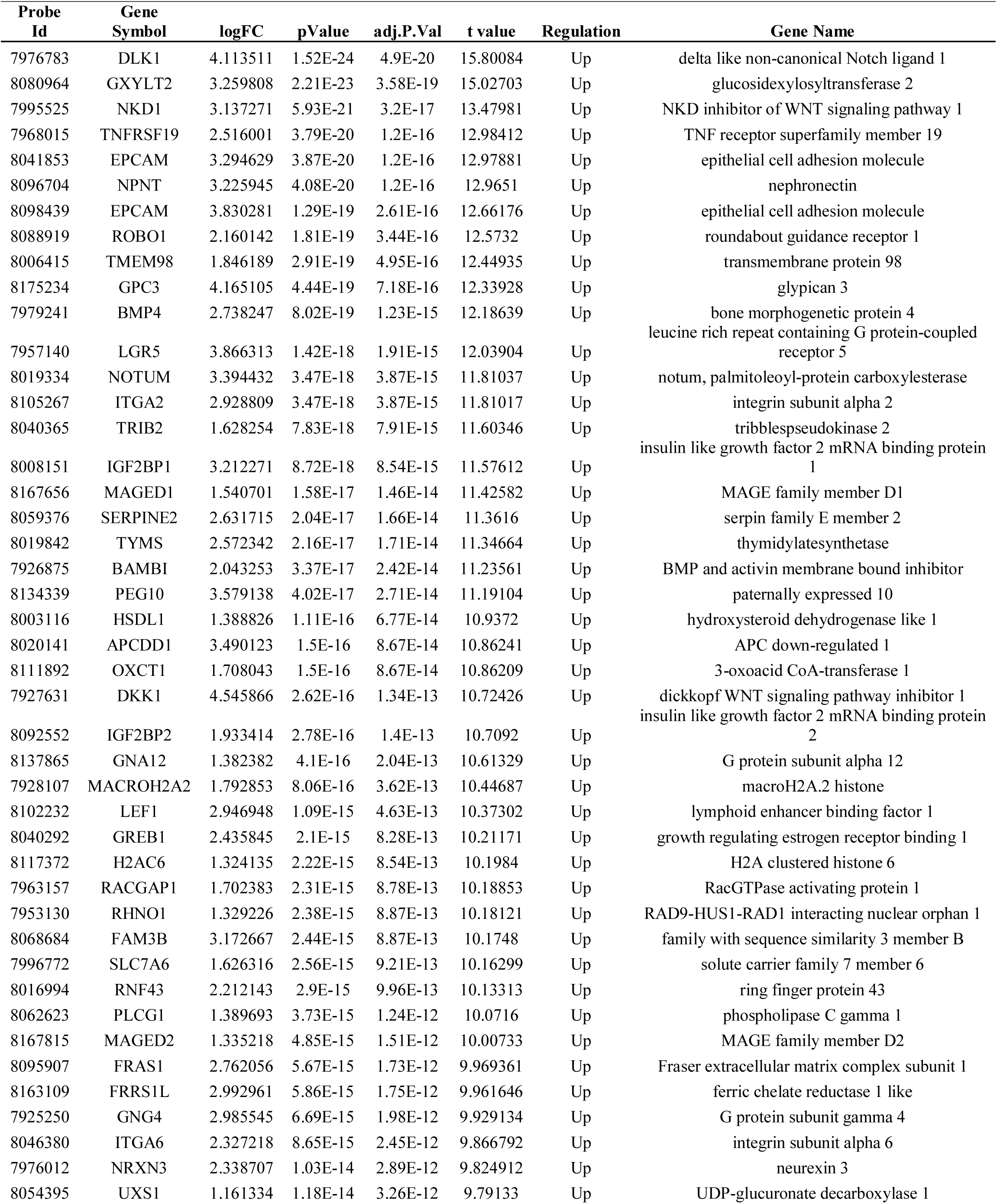
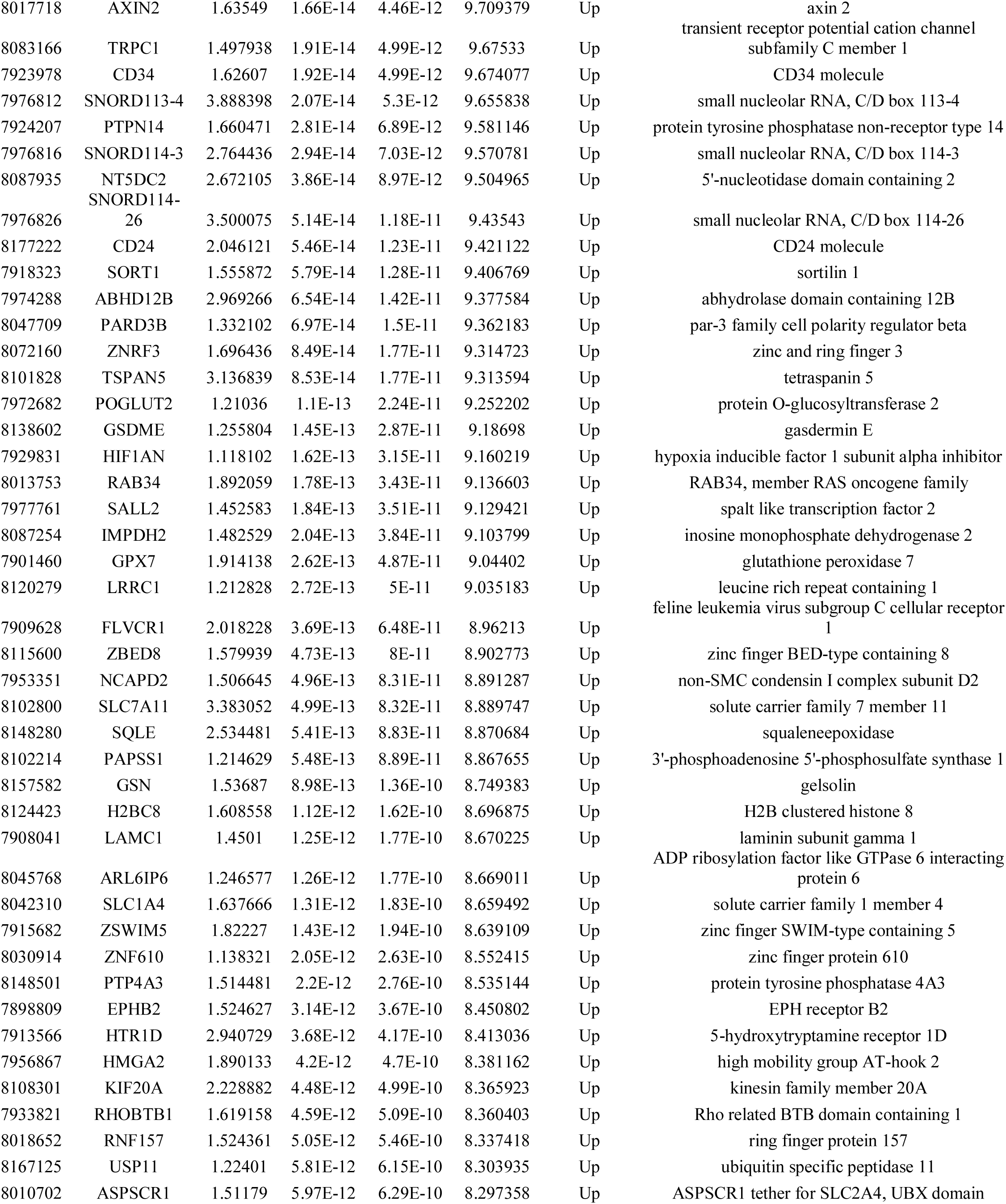
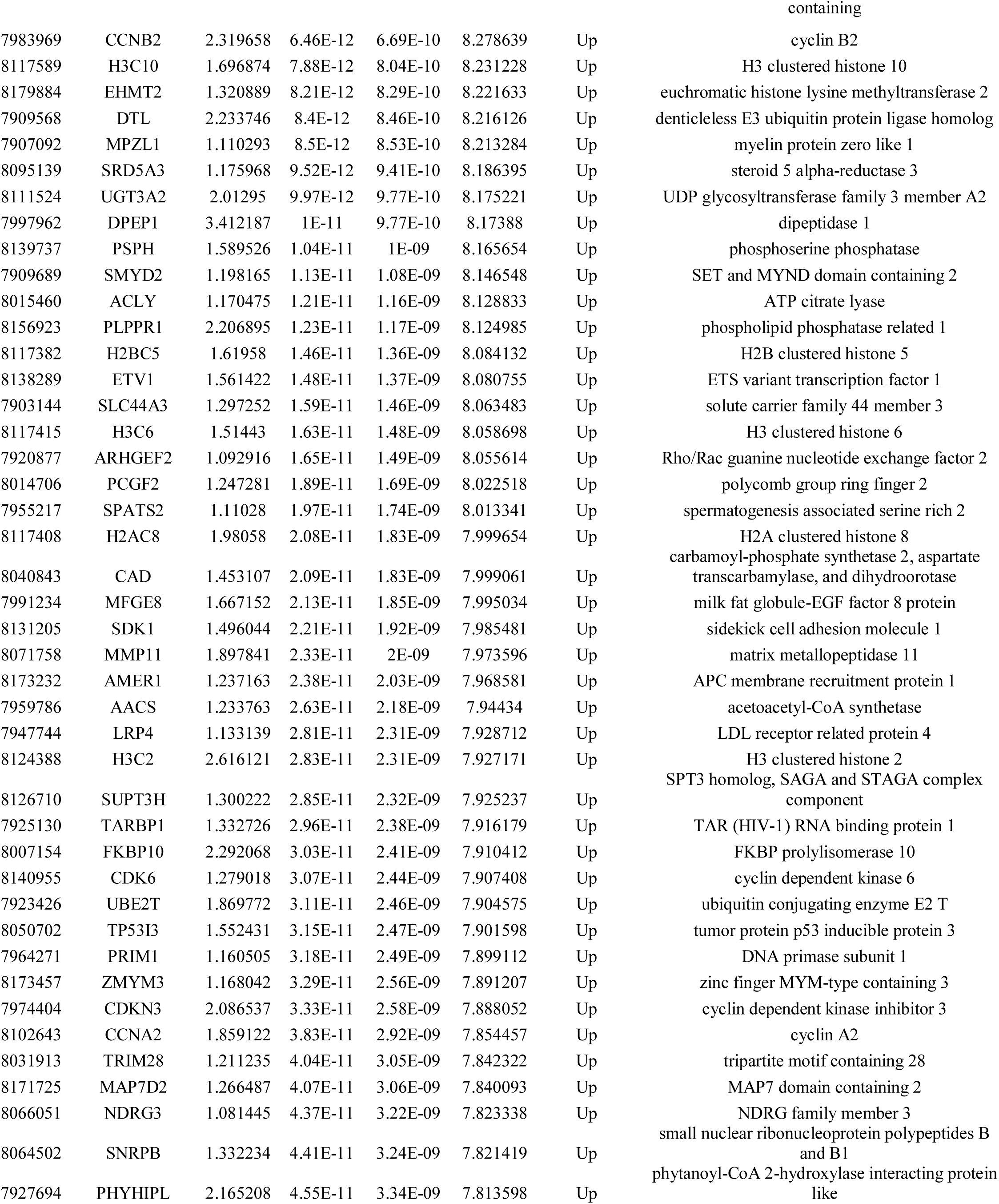
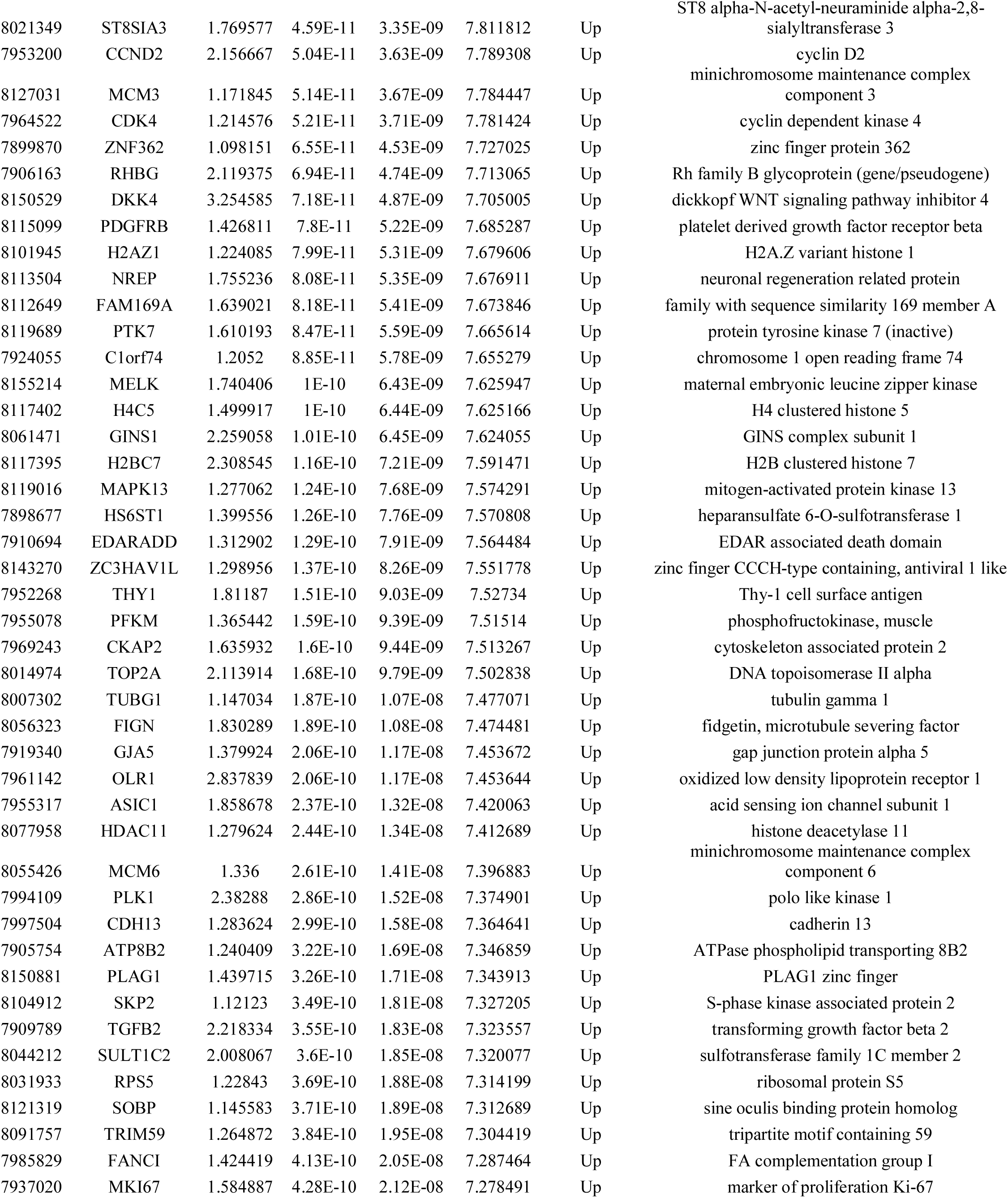
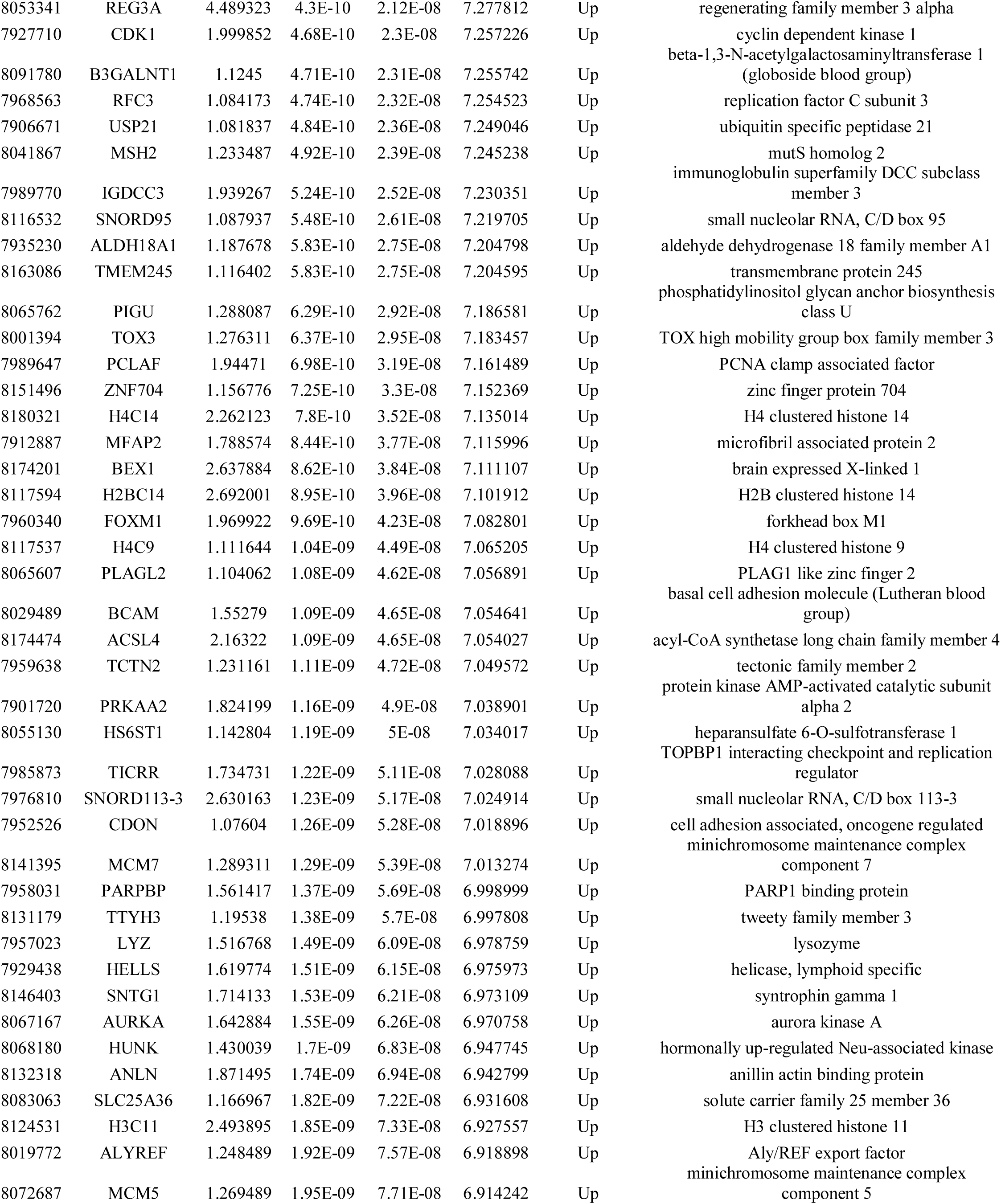
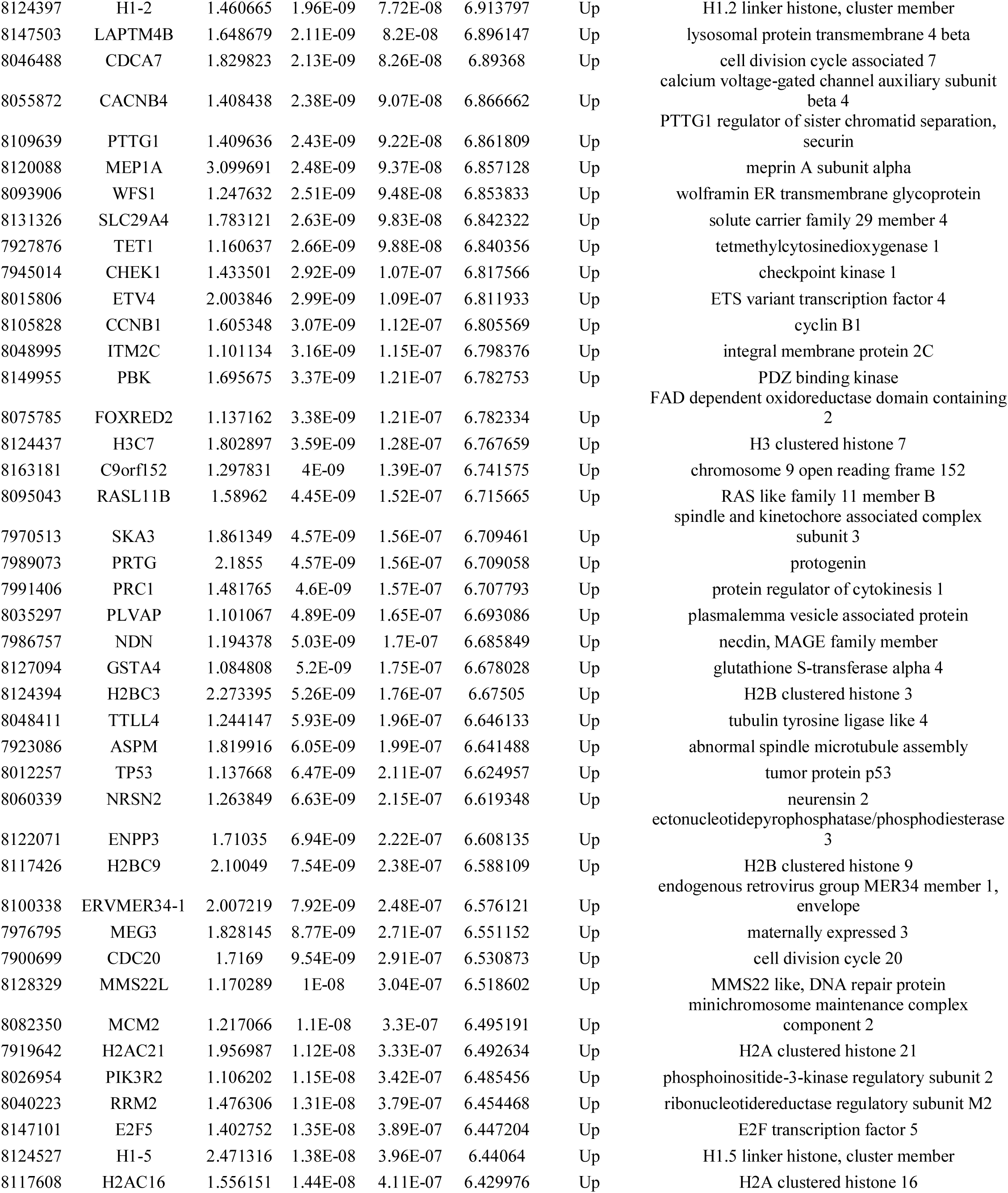
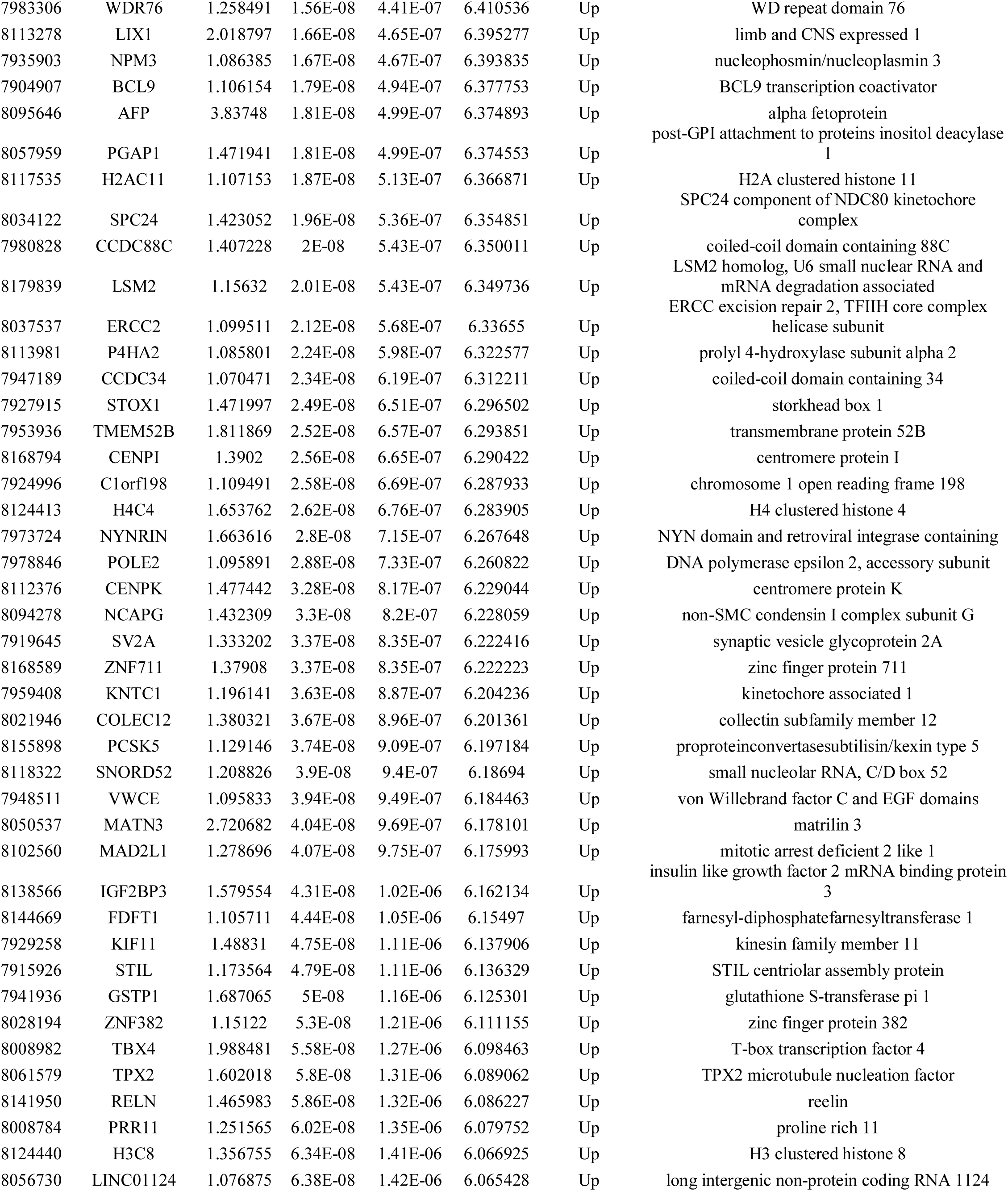
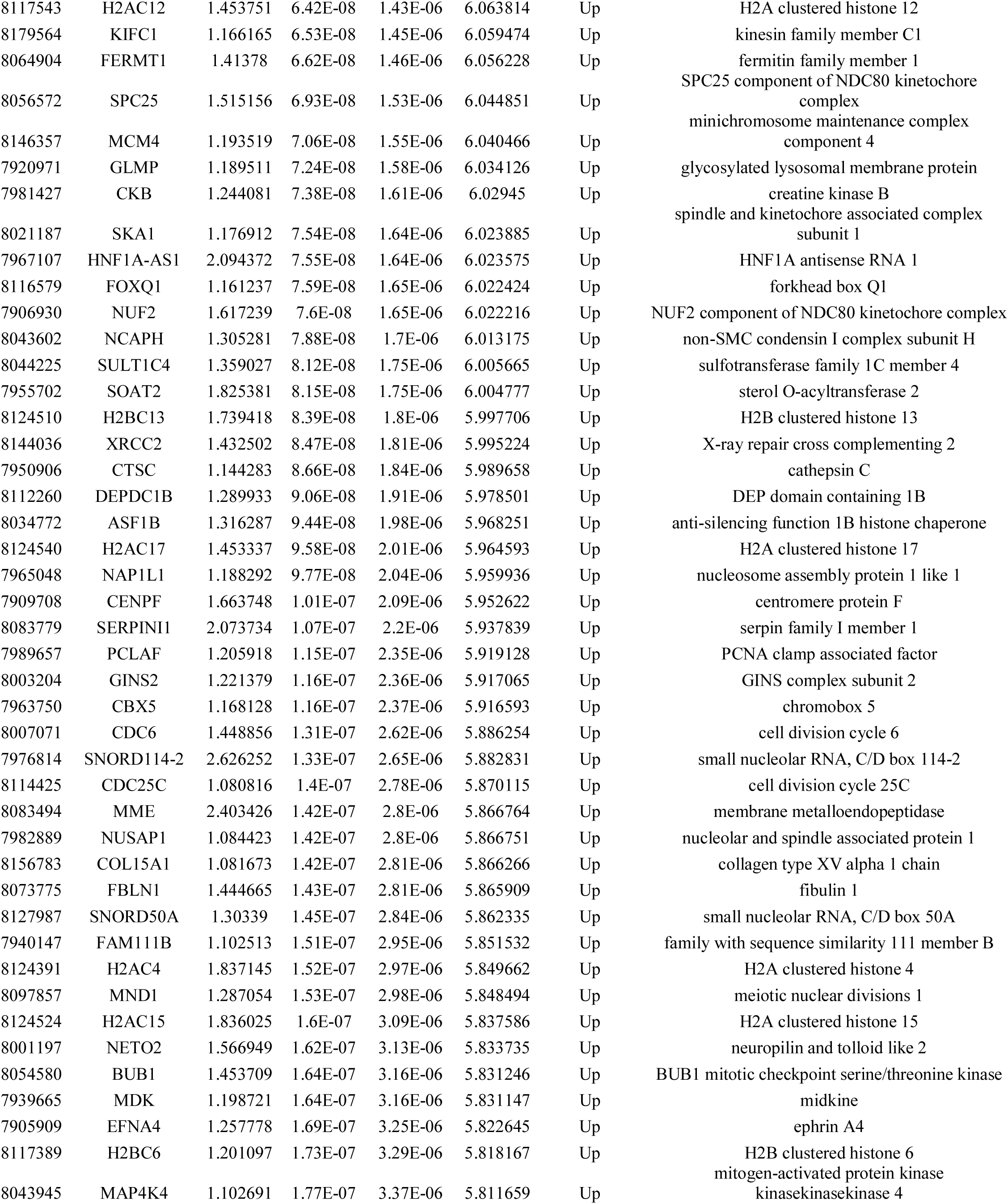
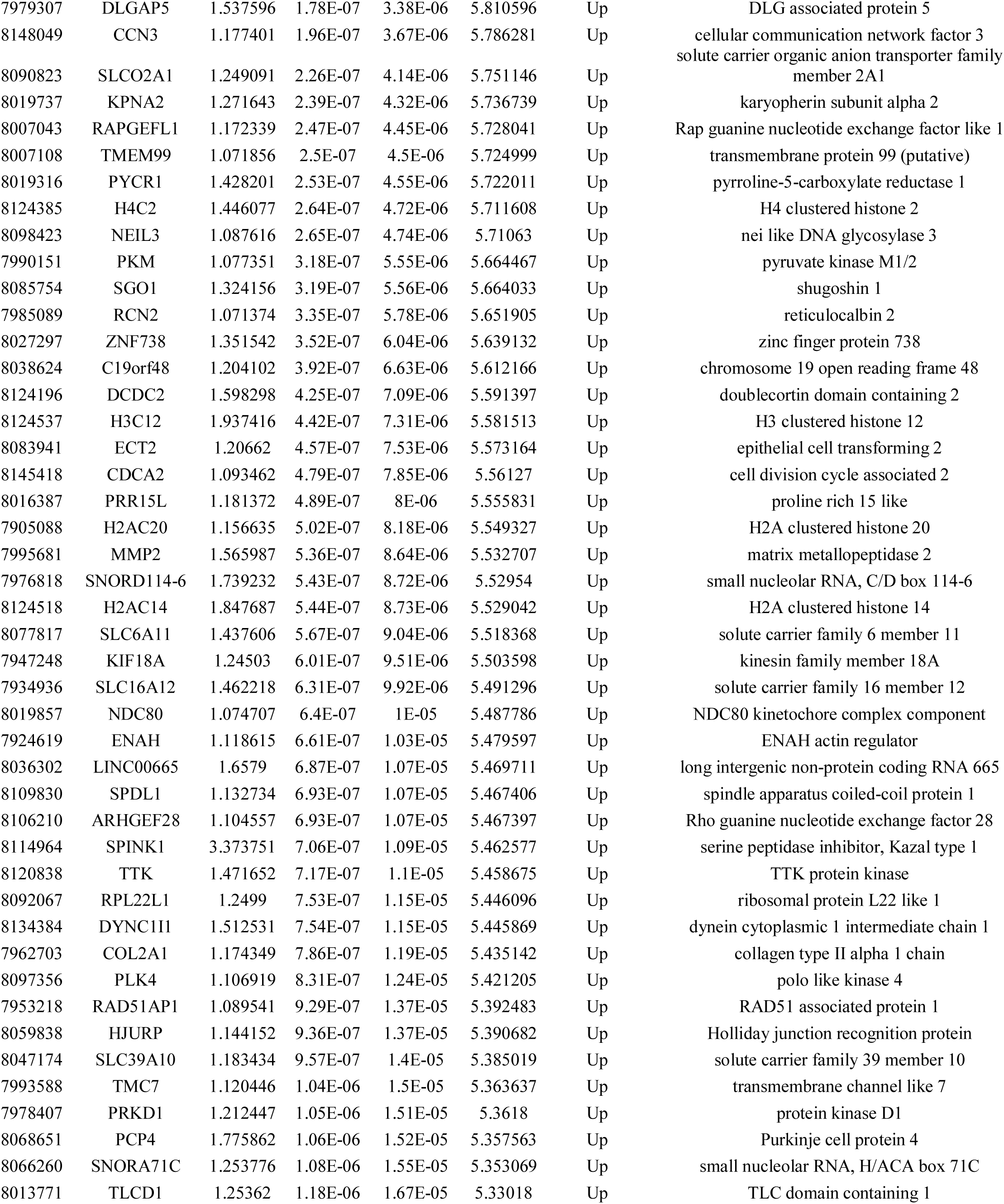
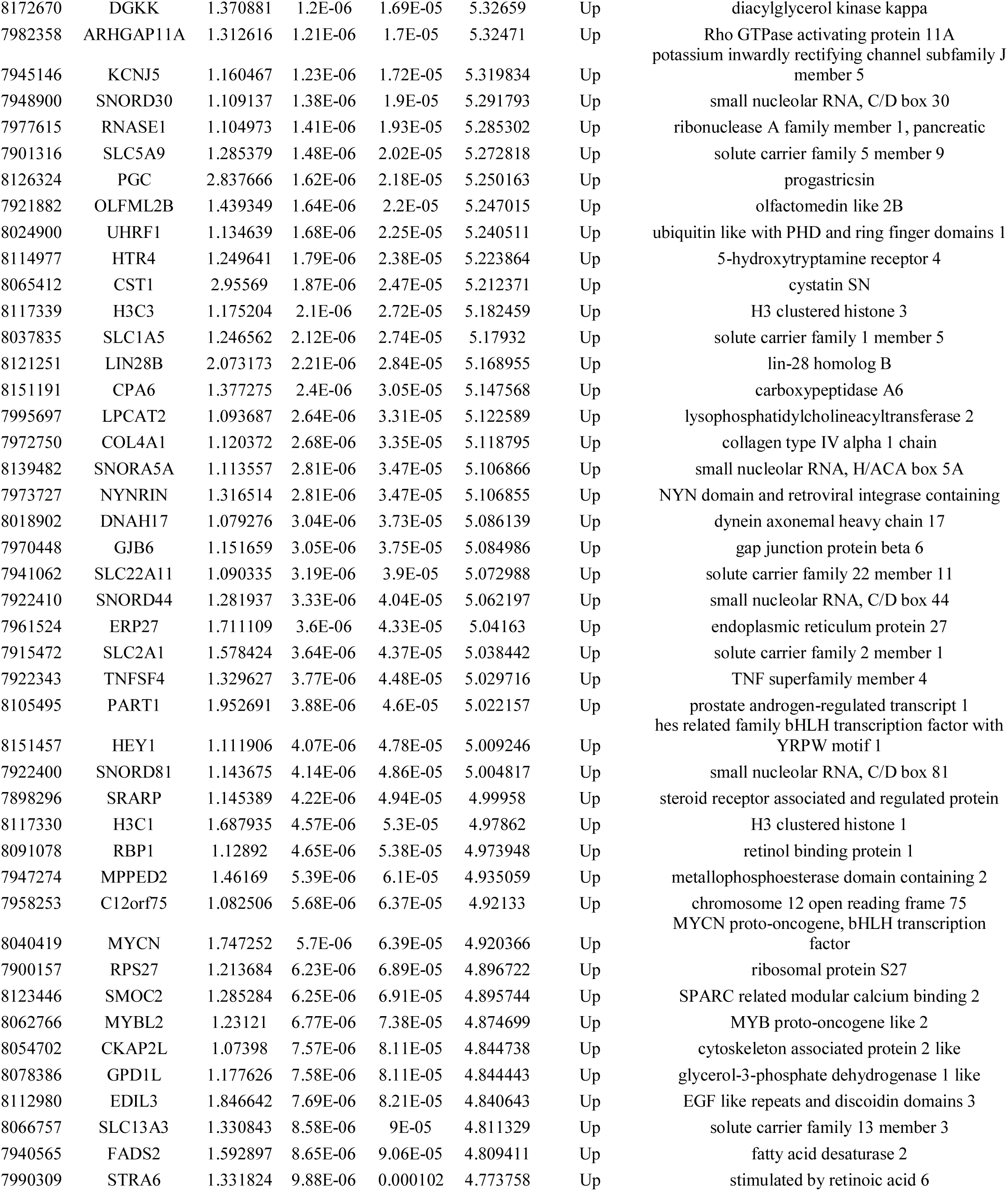
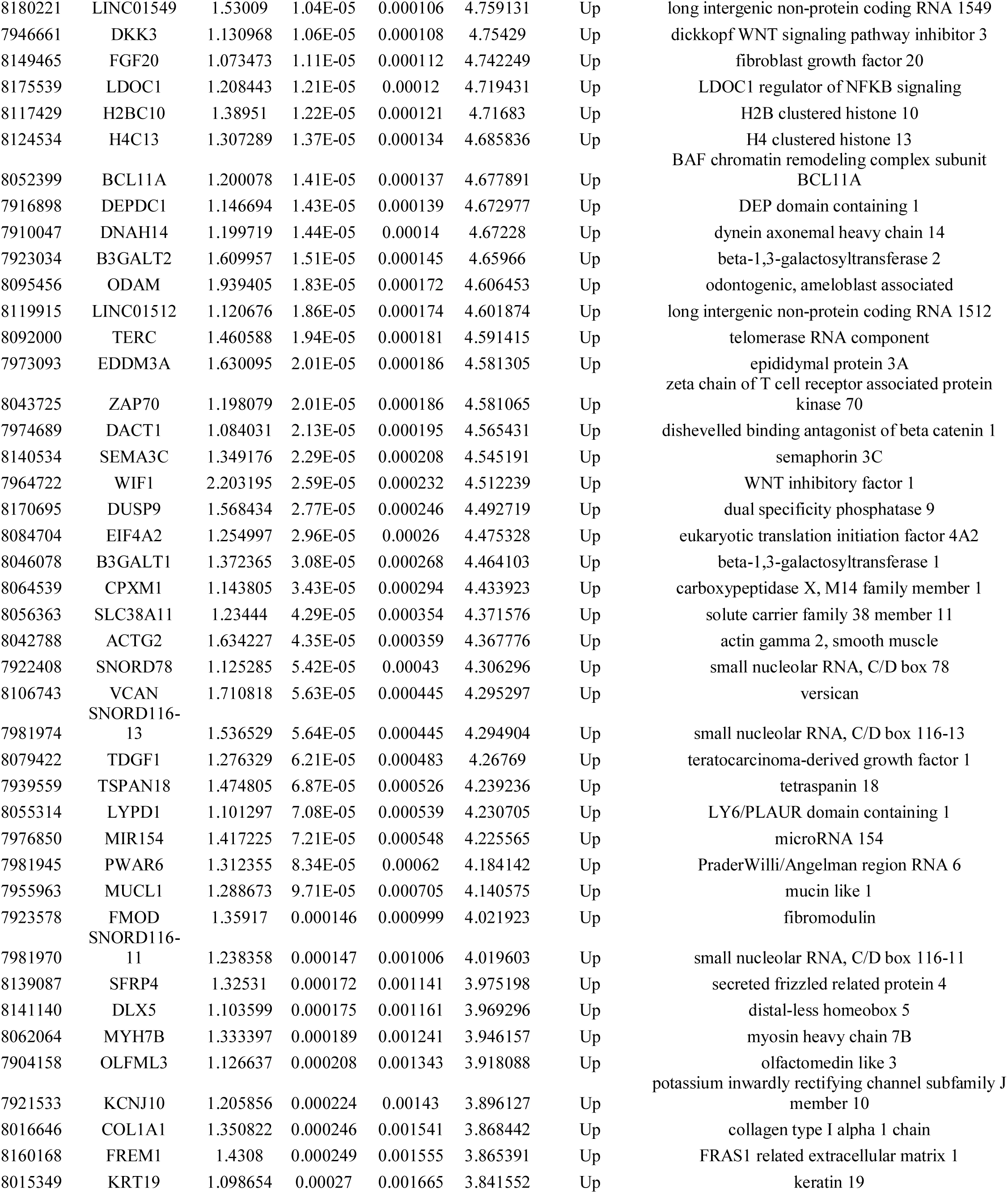
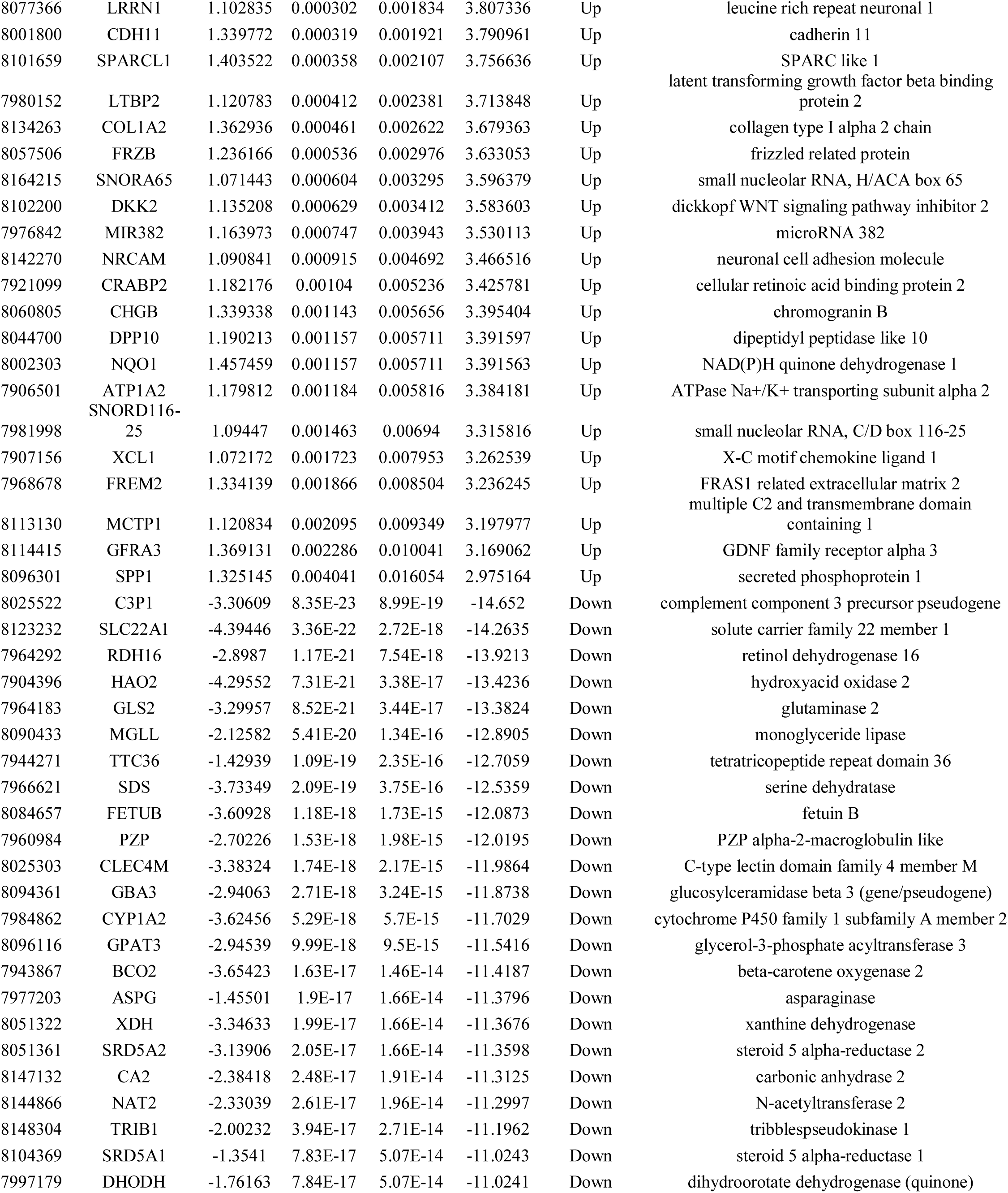
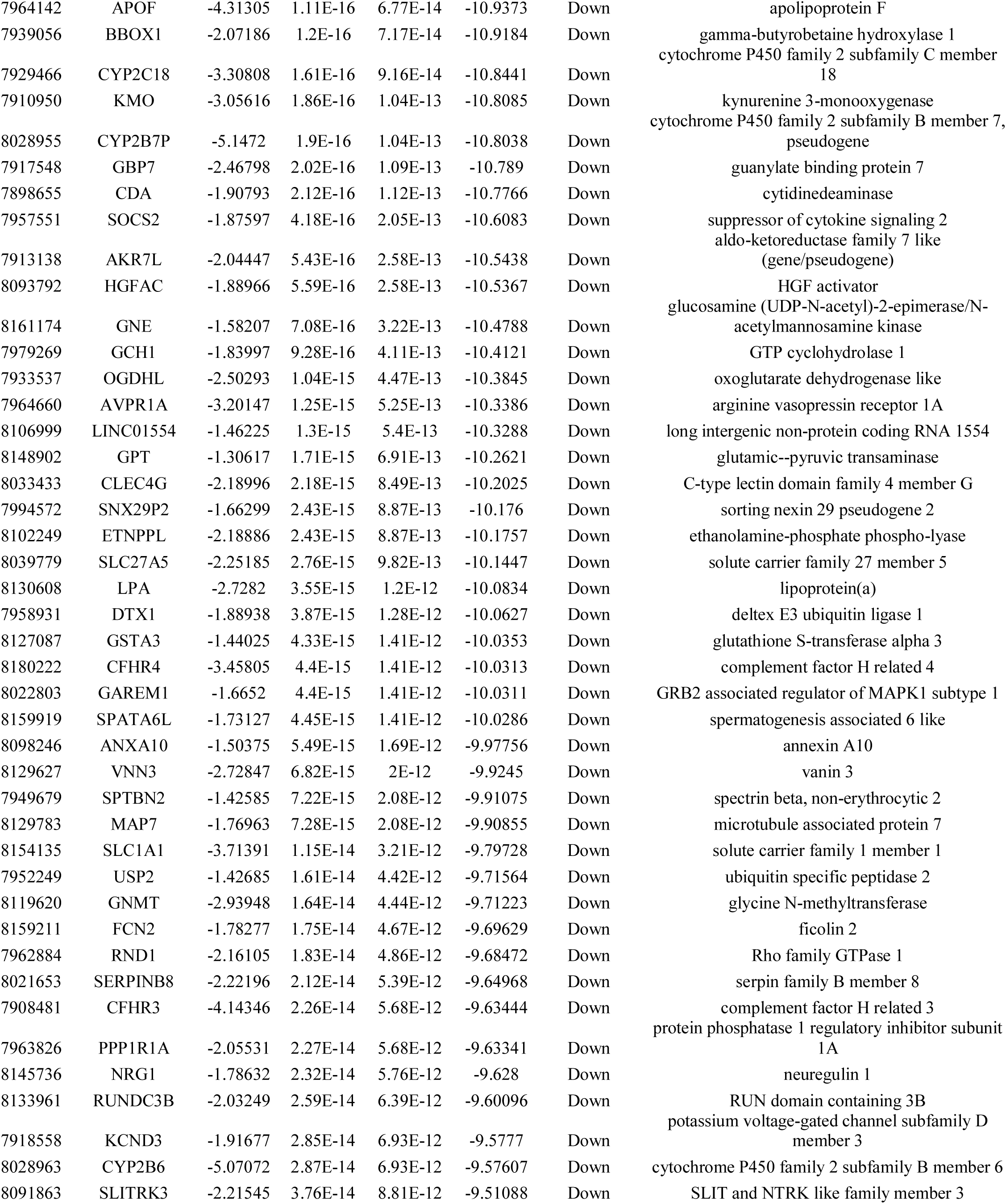
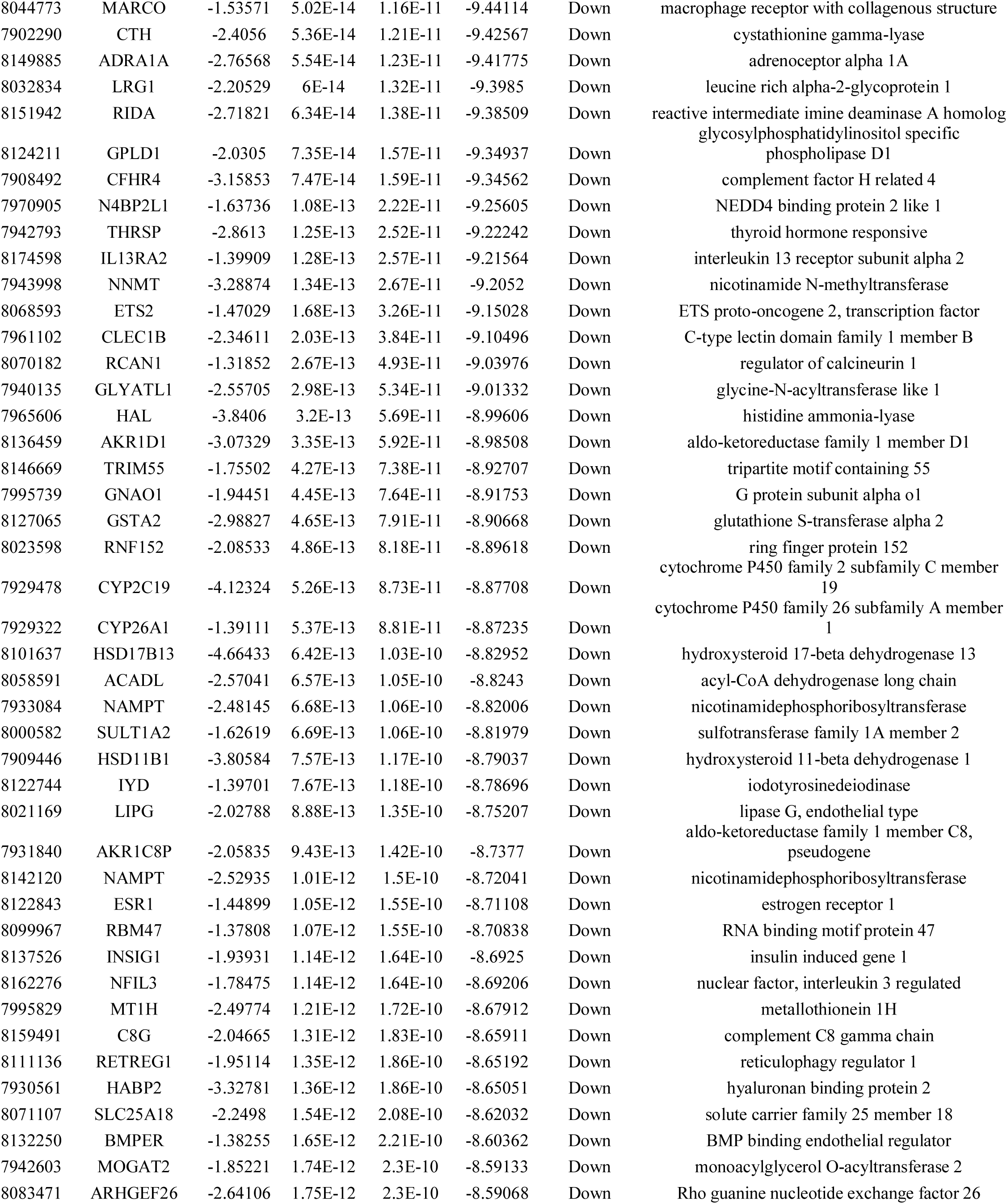
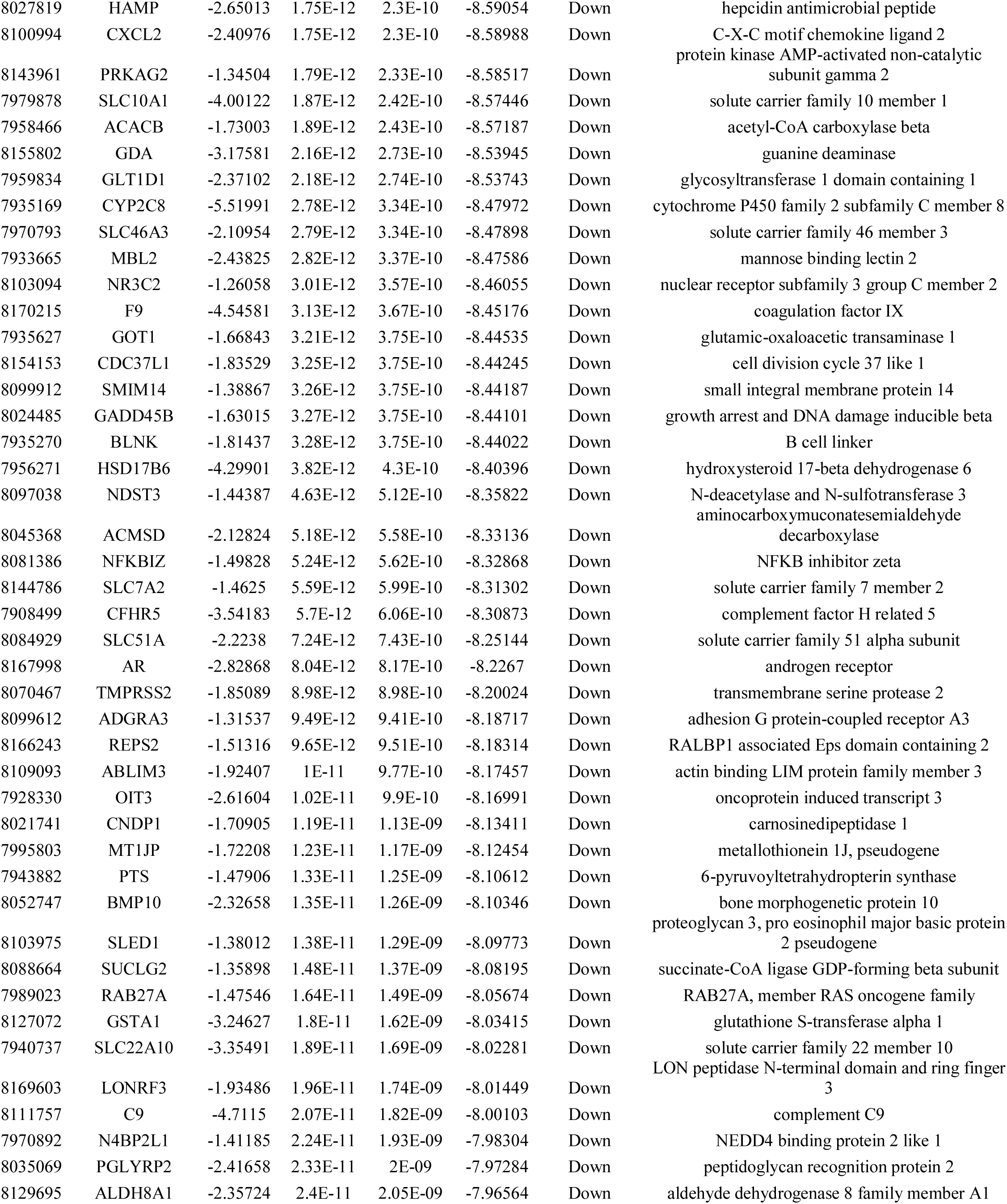
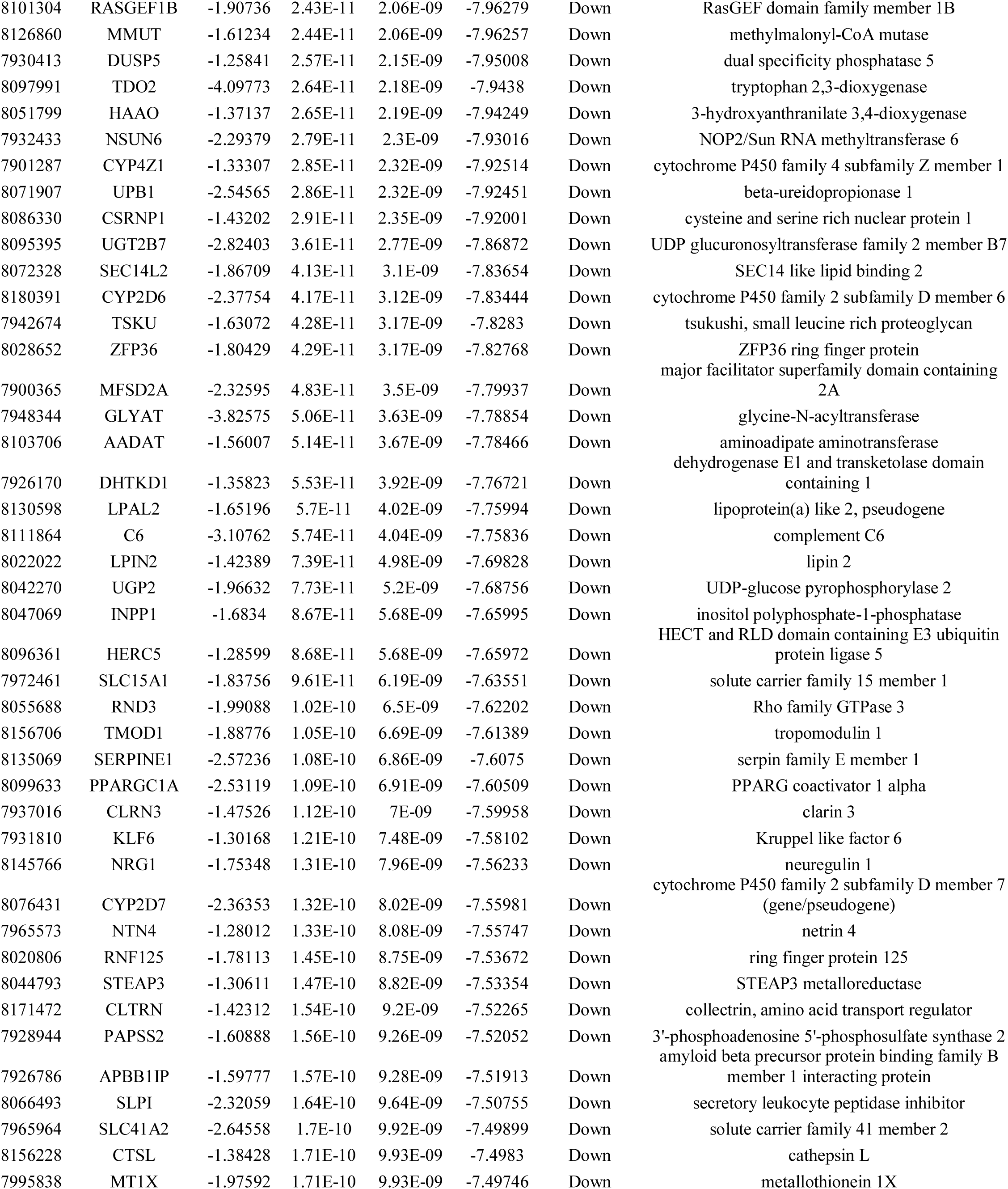
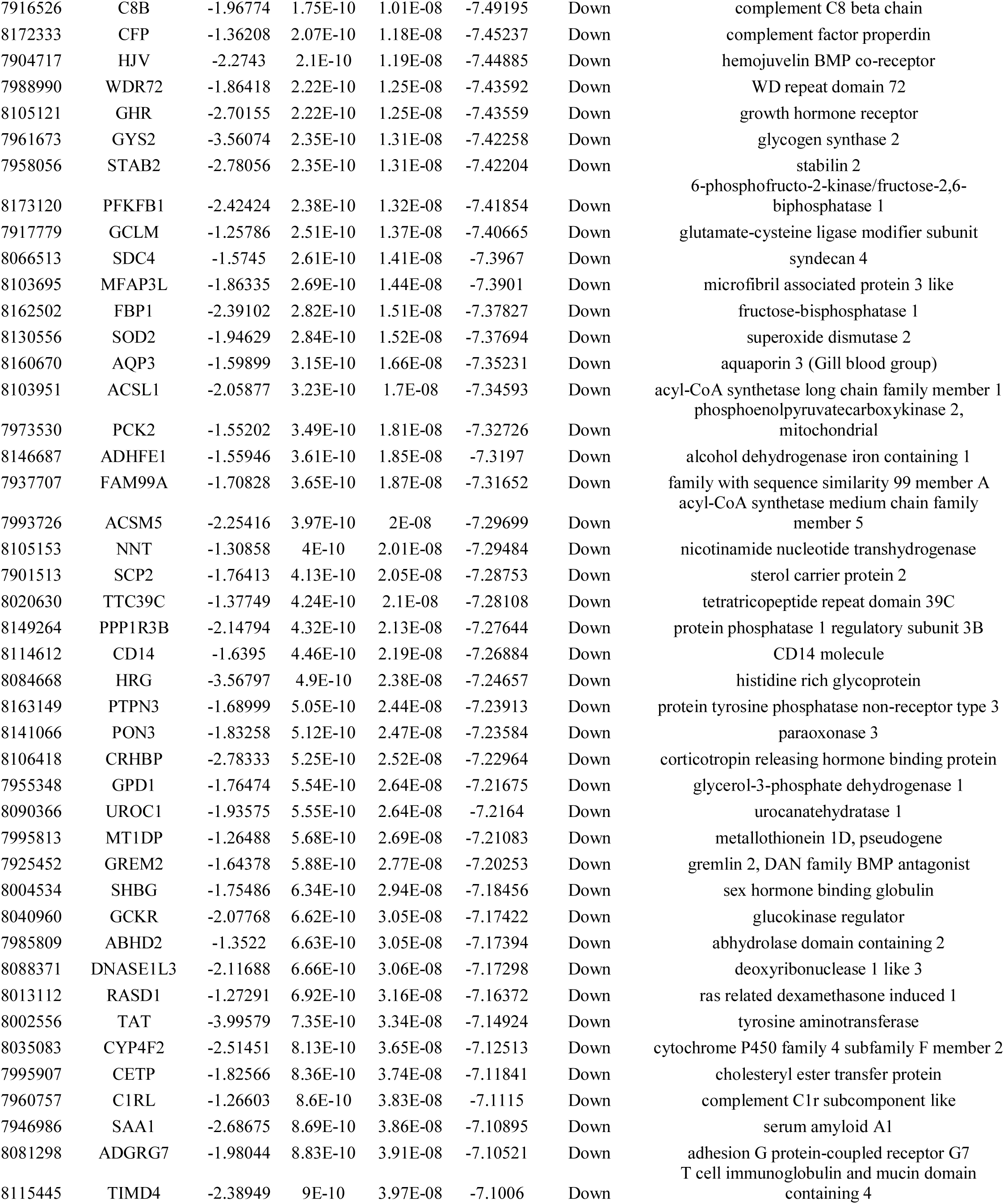
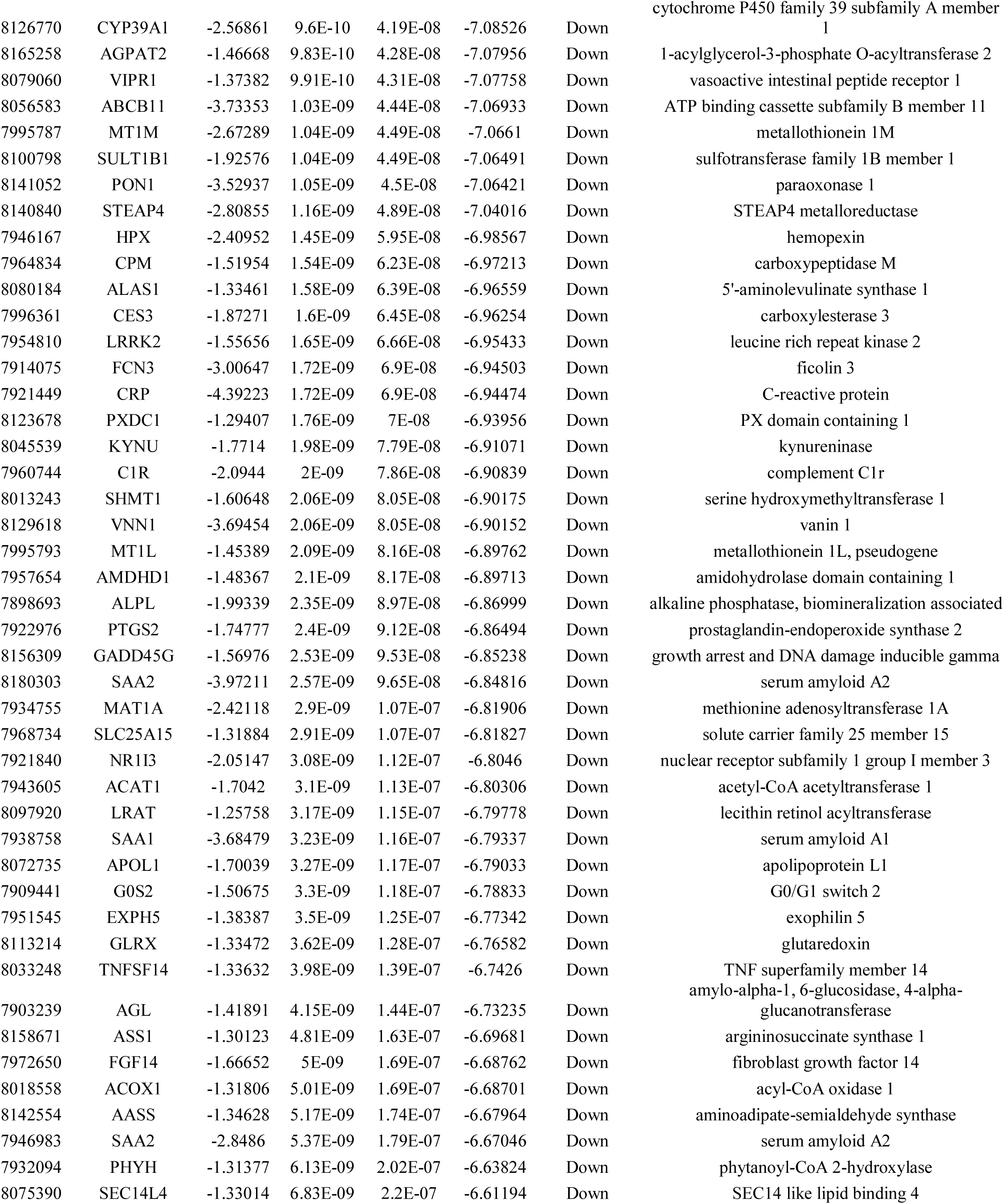
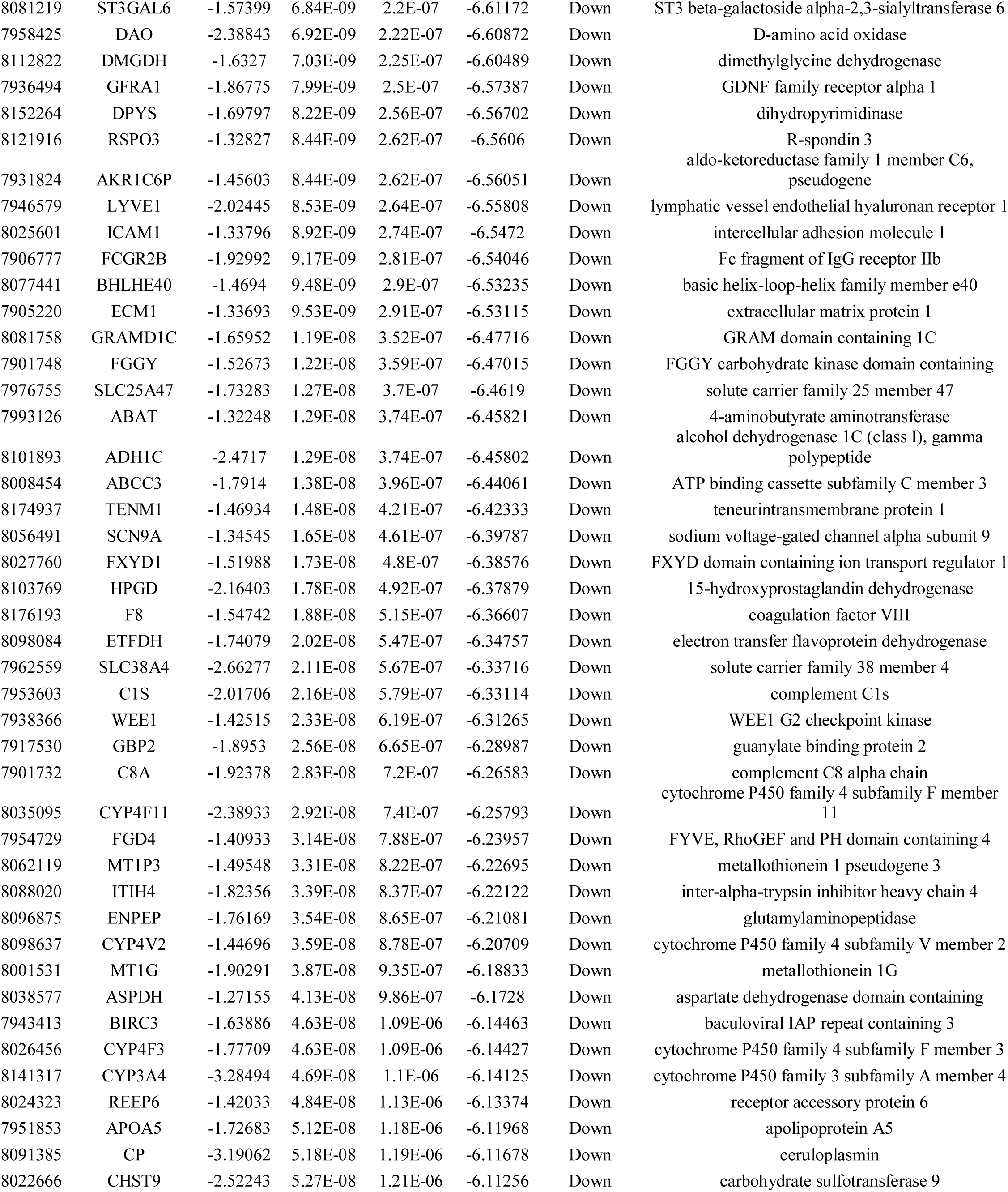
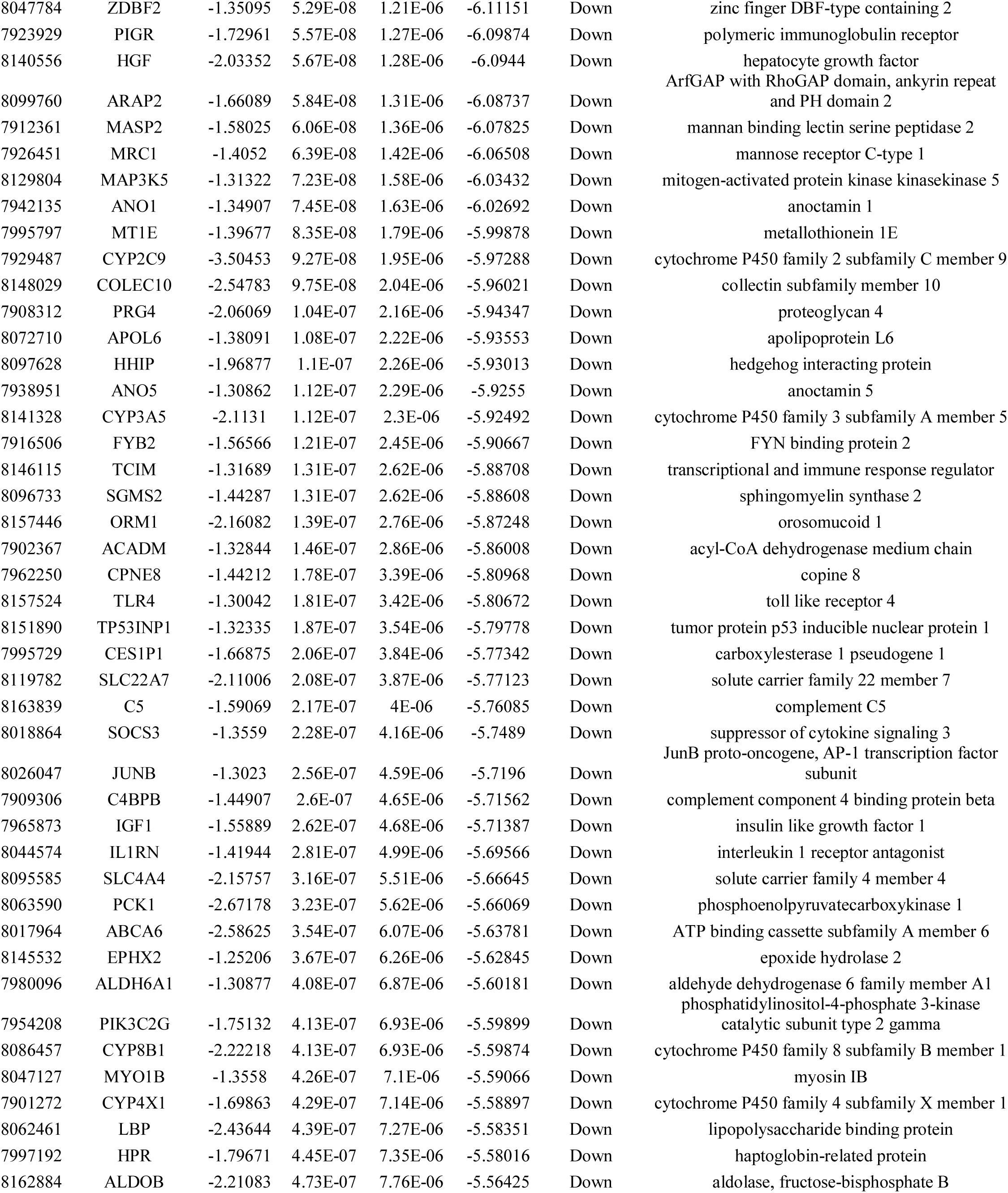
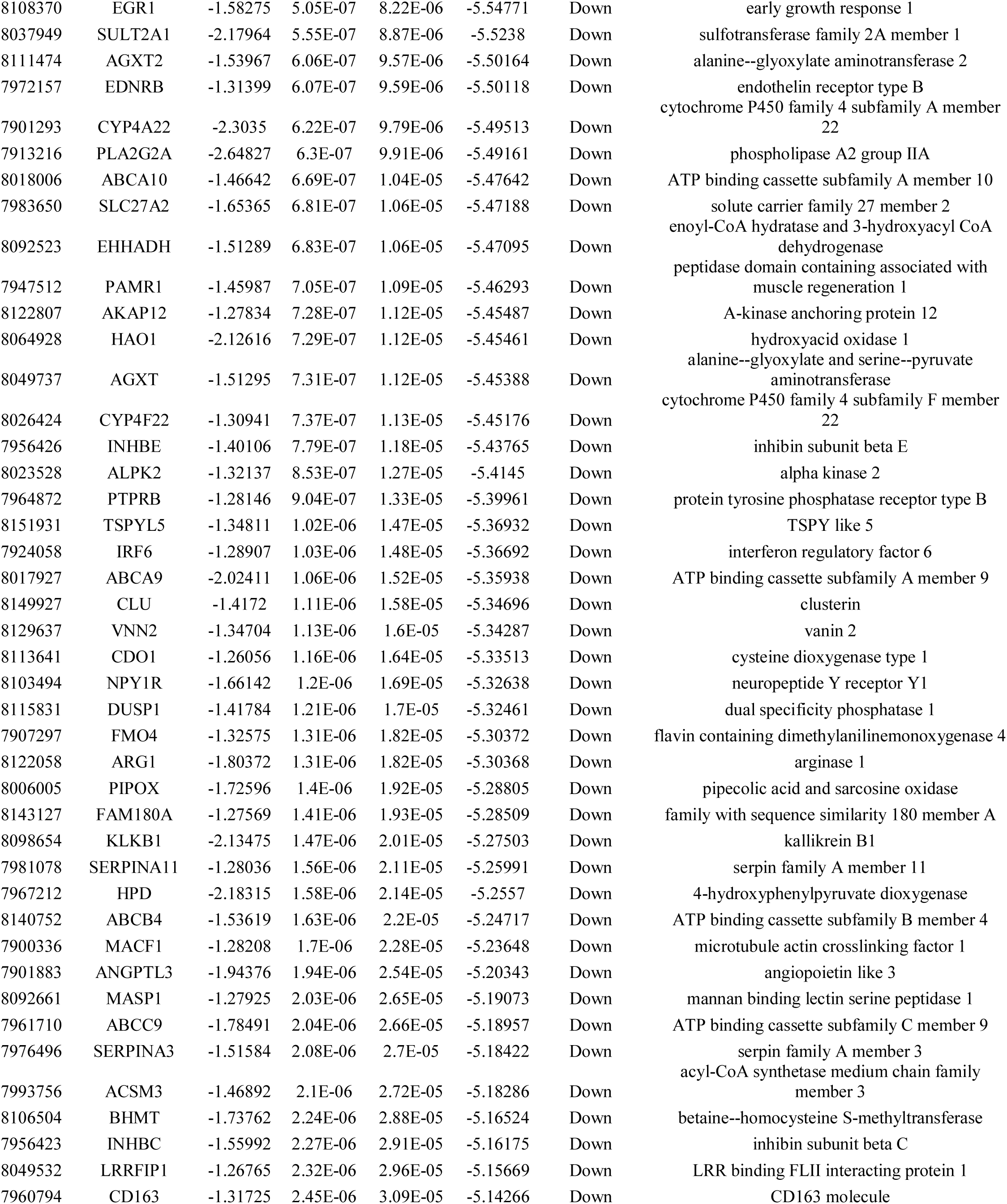
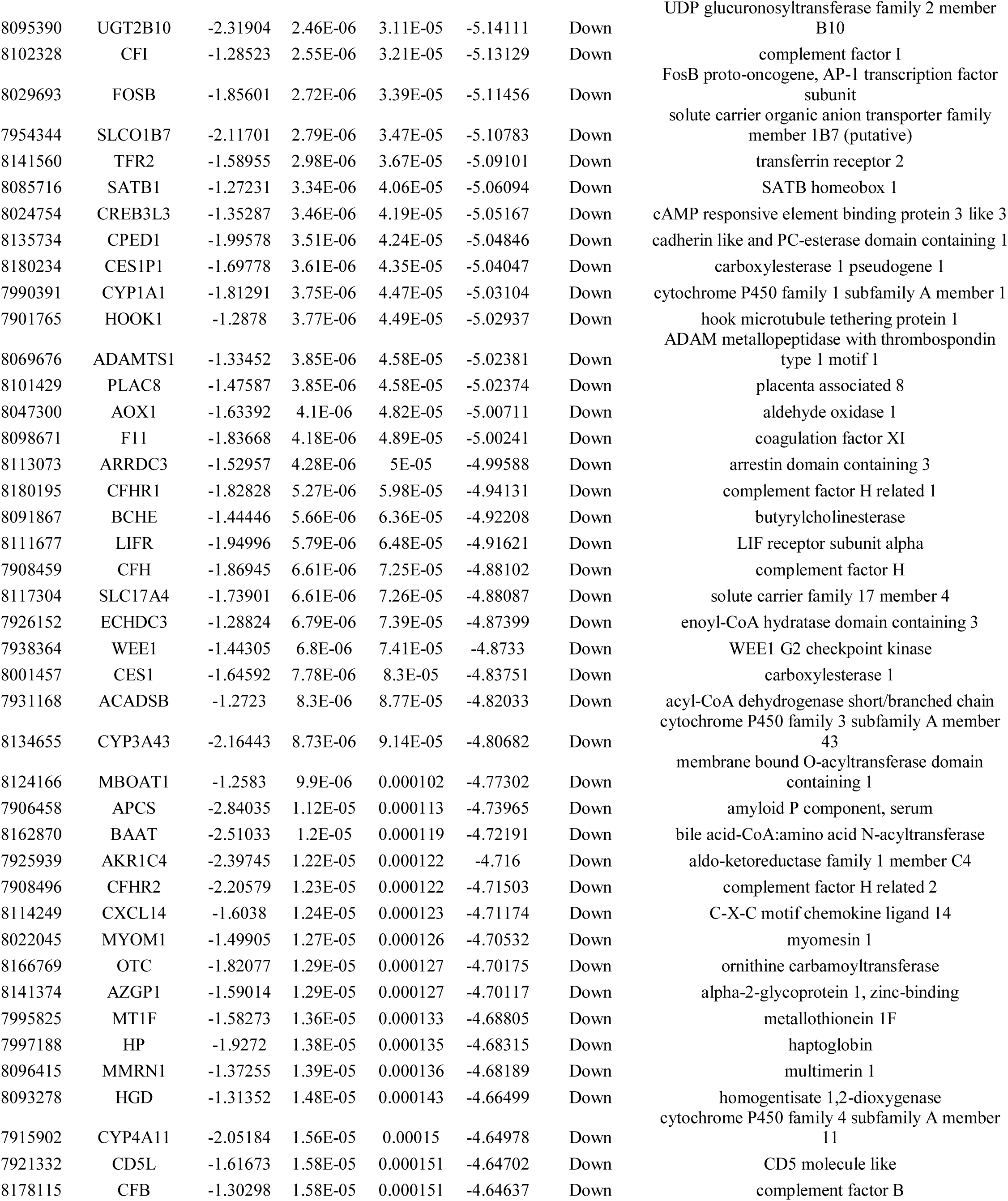
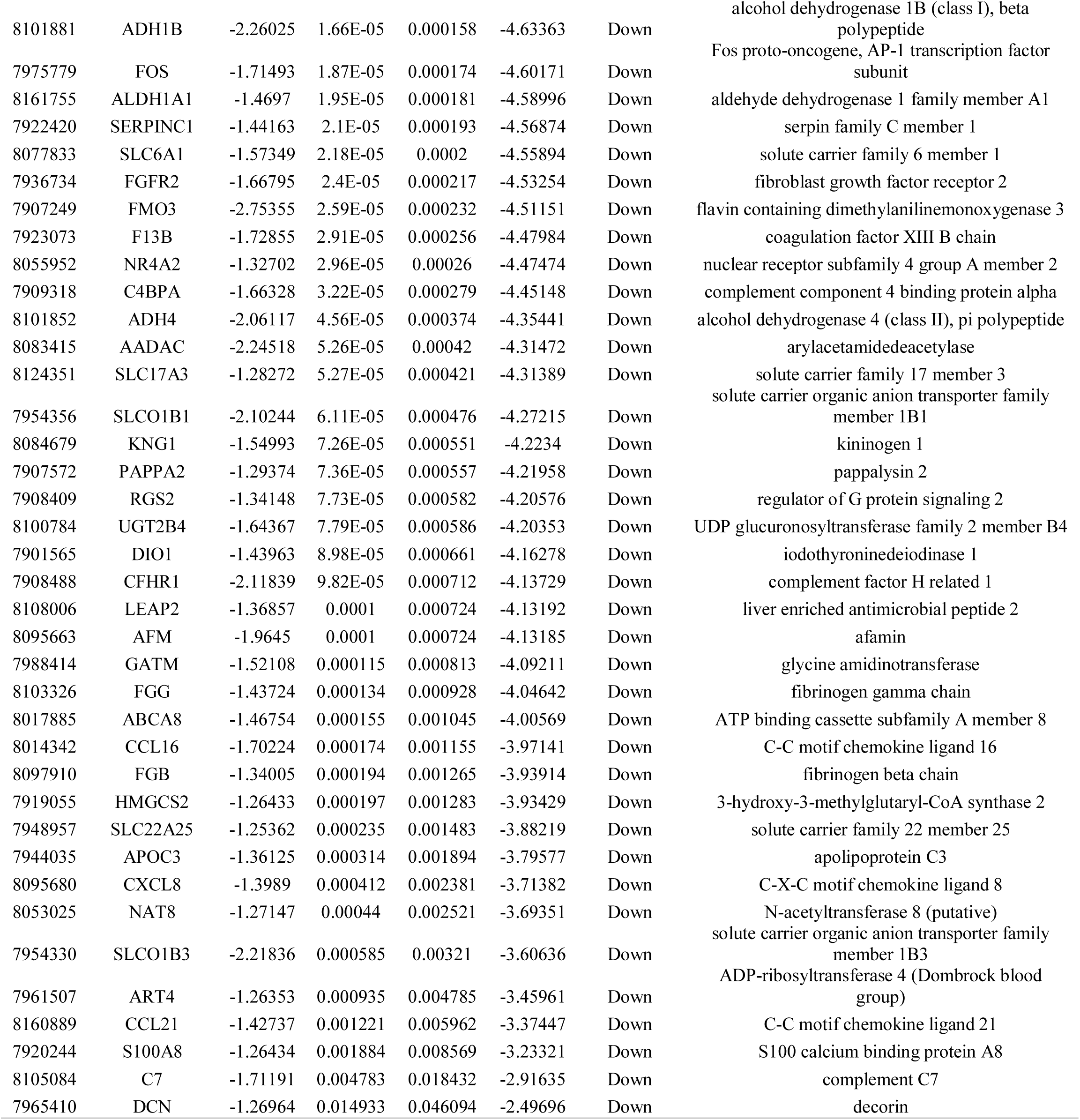
The statistical metrics for key differentially expressed genes (DEGs)

### Pathway enrichment analysis of DEGs

Pathway enrichment analyses for the up regulated and down regulated genes were performed by ToppGene. The up regulated genes were mainly enriched in proline biosynthesis, epoxysqualene biosynthesis, alcoholism, cell cycle, HDACs deacetylate histones, sterol biosynthesis, urea cycle and metabolism of amino groups, PLK1 signaling events, aurora B signaling, CDK regulation of DNA replication, Wnt/beta-catenin pathway, Wnt signaling pathway, DNA replication, G2/M DNA damage checkpoint, steroid biosynthetic, lysinuric protein intolerance and risedronate pathway are listed in Table 3. Similarly, the down regulated genes were mainly enriched in superpathway of tryptophan utilization, bile acid biosynthesis, neutral pathway, metabolic pathways, complement and coagulation cascades, ATF-2 transcription factor network, IL6-mediated signaling events, biological oxidations, complement cascade, fatty acid metabolism, tryptophan metabolism, complement pathway, intrinsic prothrombin activation pathway, blood coagulation, pyrimidine metabolism, tryptophan metabolic, statin pharmacokinetics pathway, enoxaparin pathway and glycine, serine and threonine metabolism are listed Table 4.

**Table 3.**
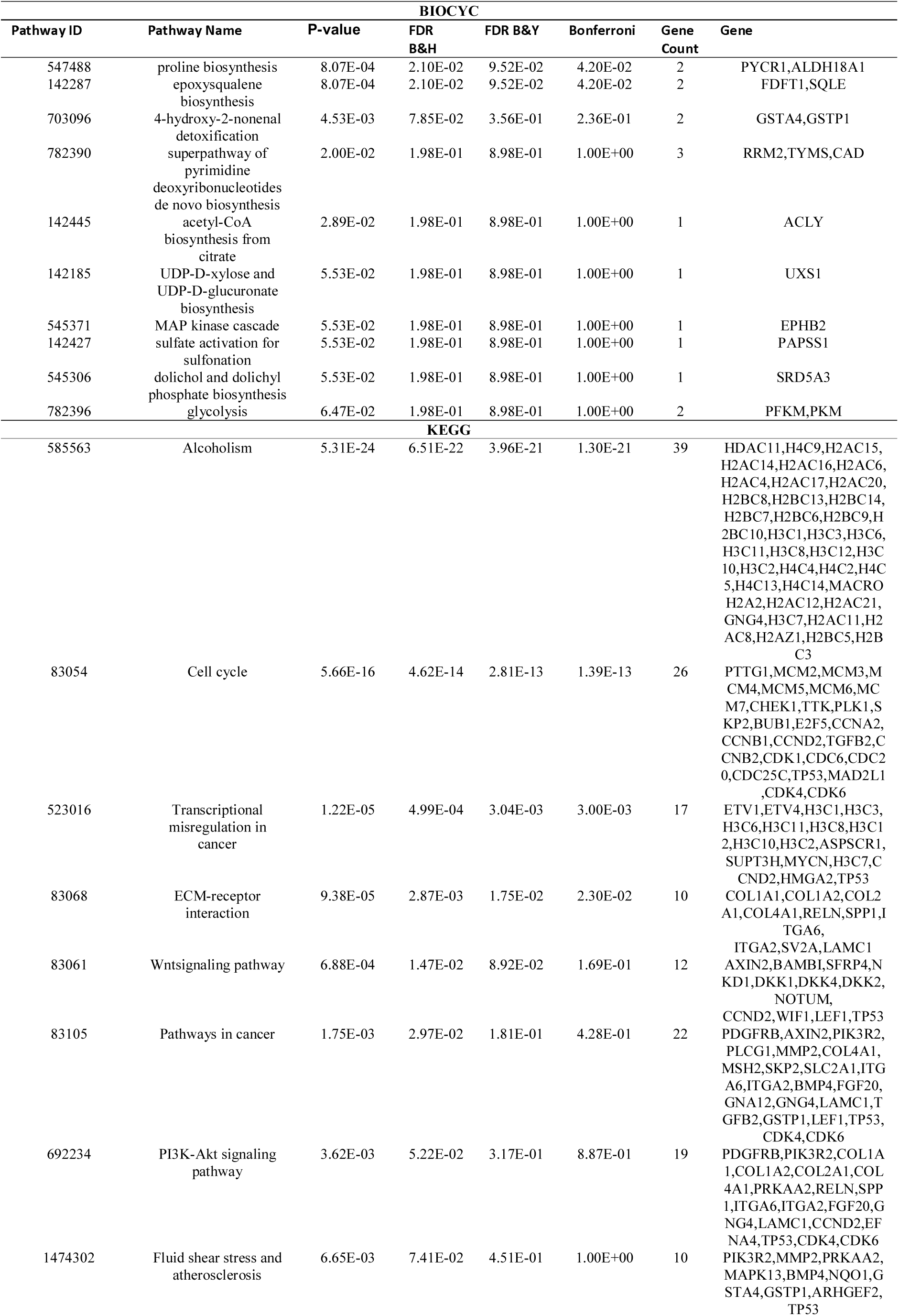
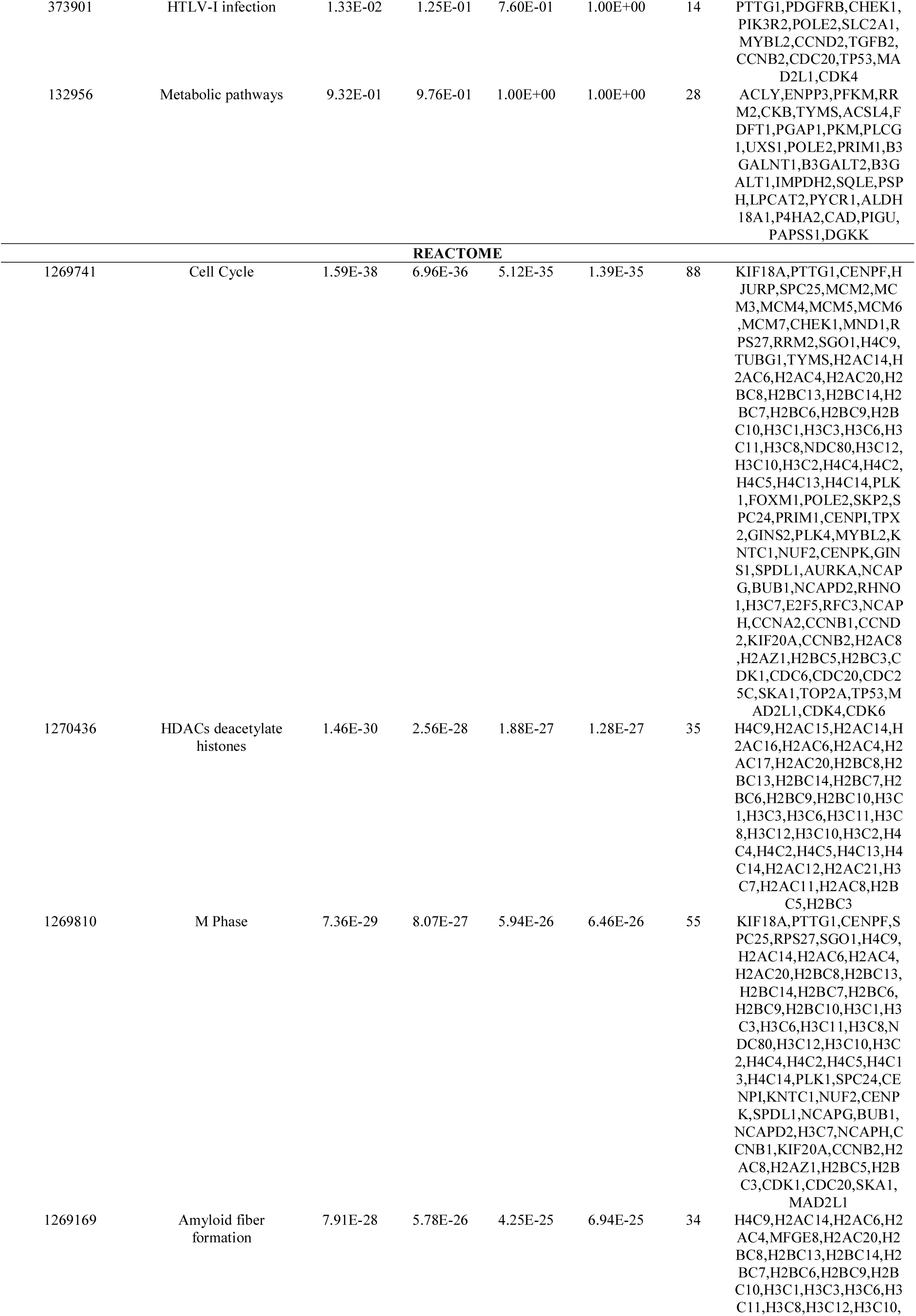
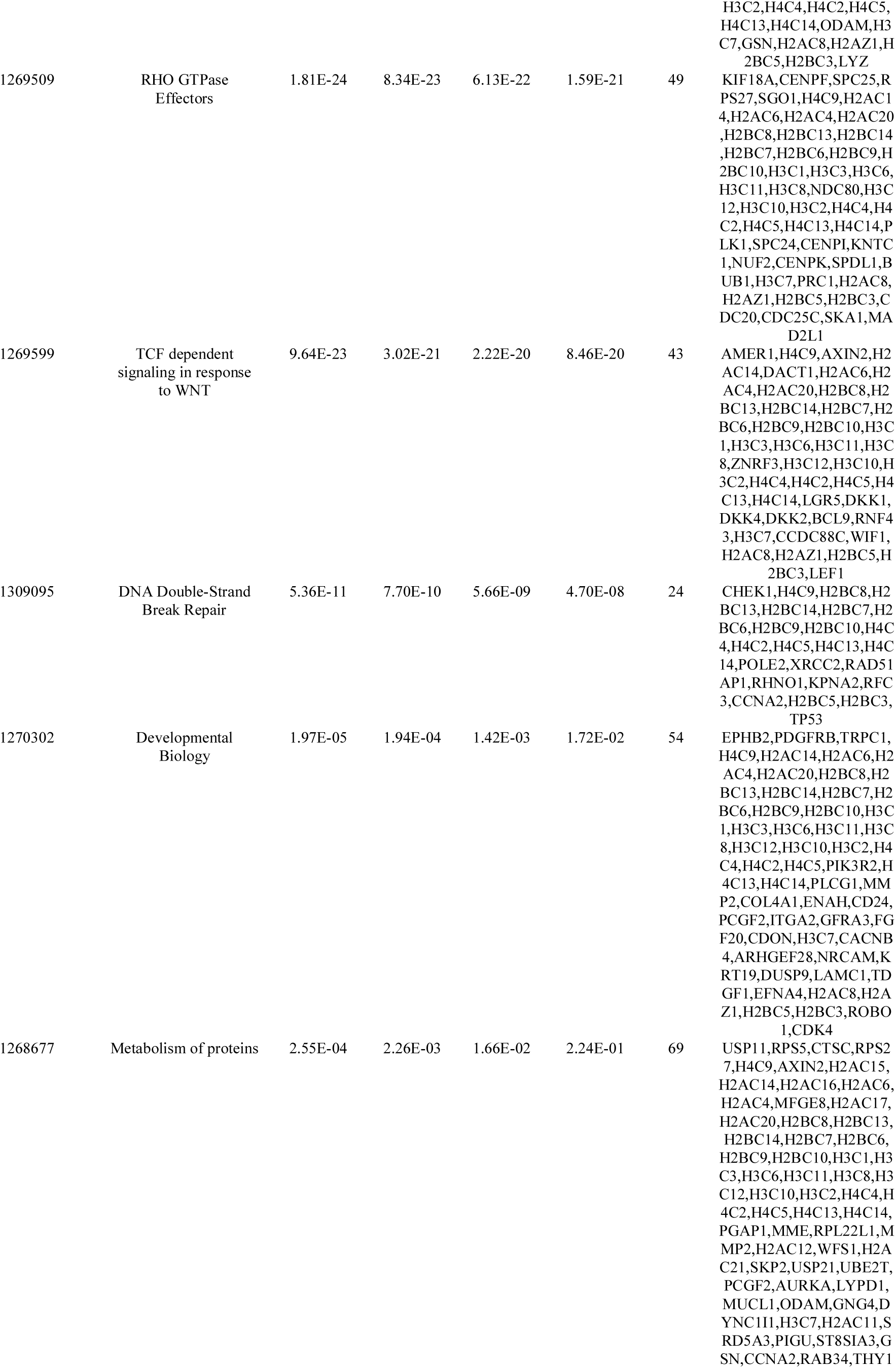
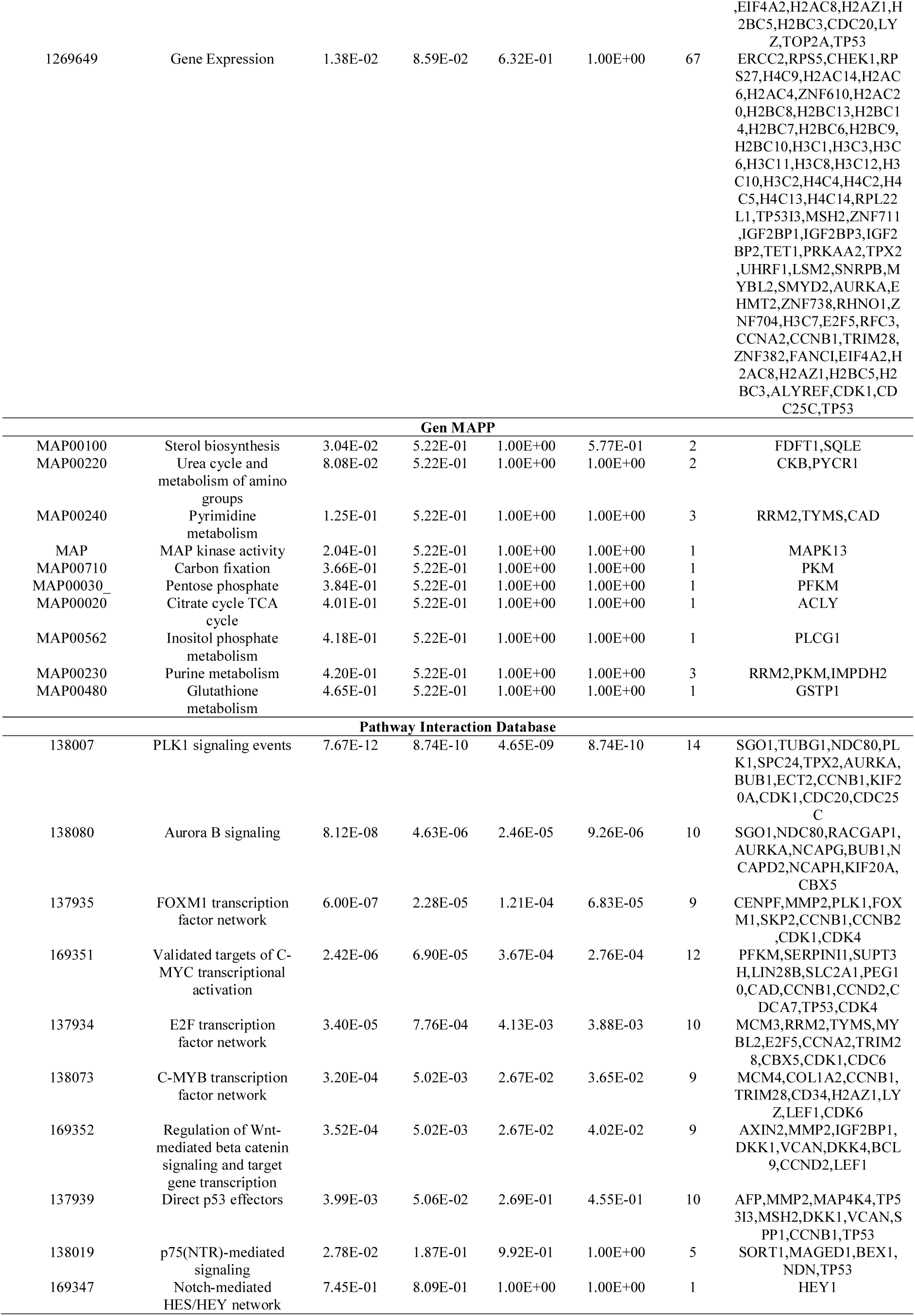
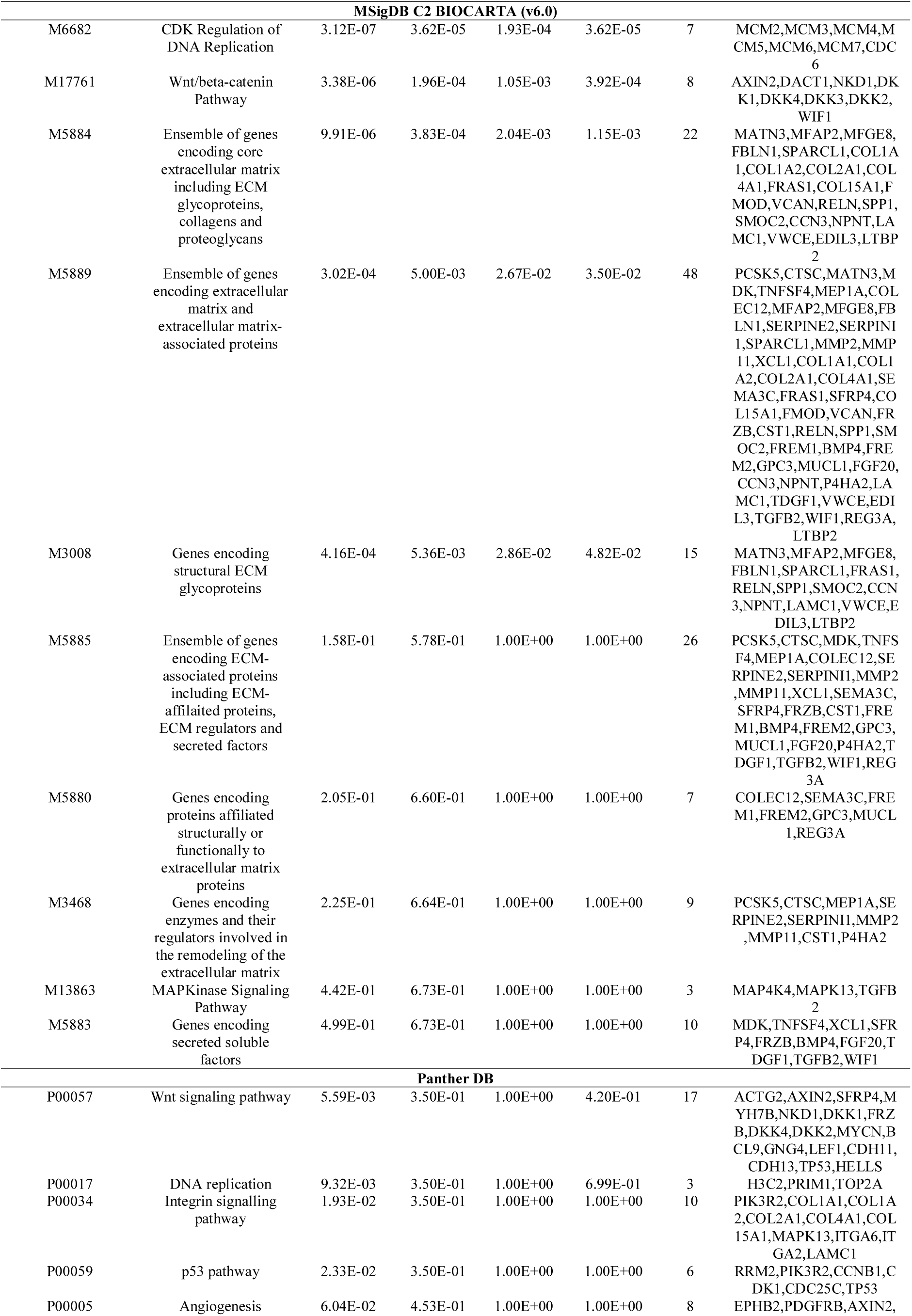
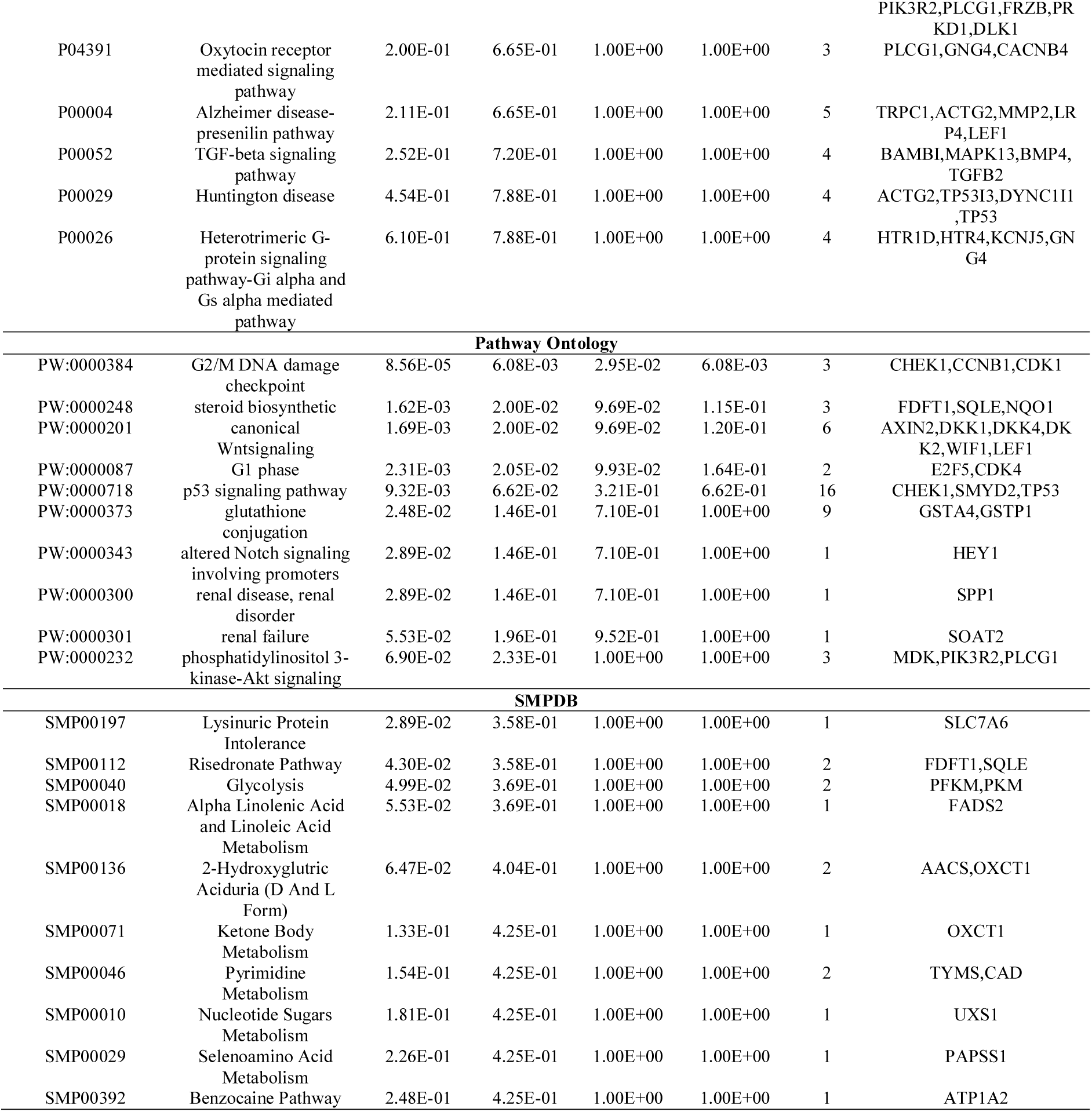
The enriched pathway terms of the up regulated differentially expressed genes

**Table 4.**
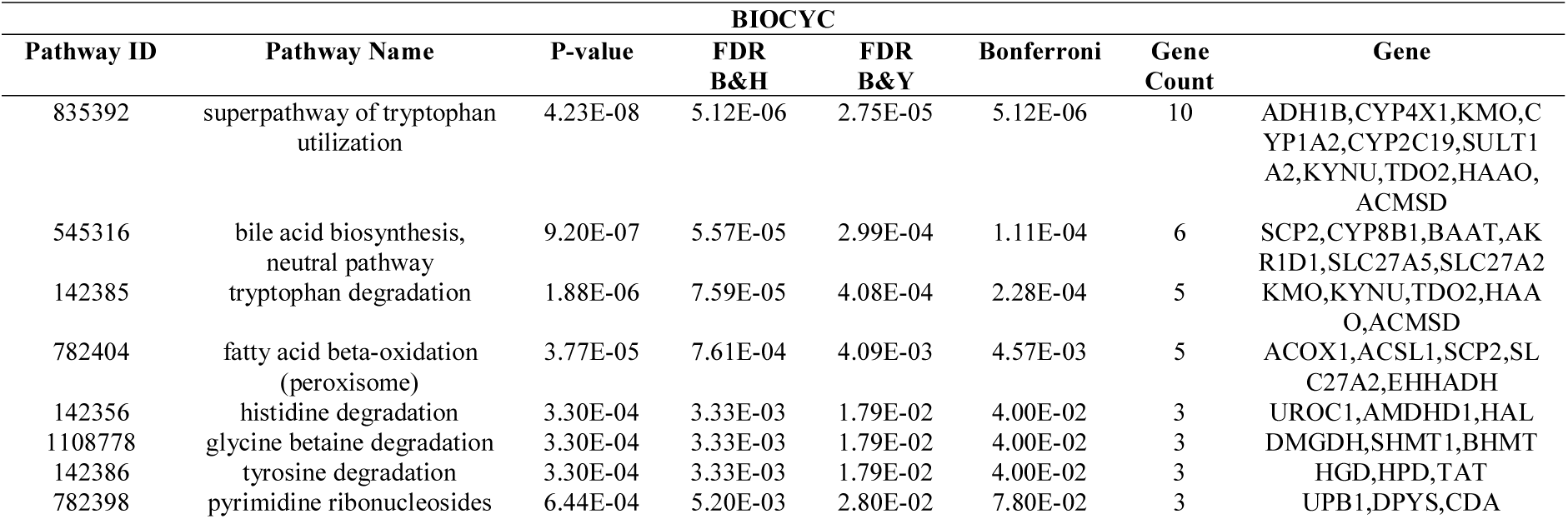
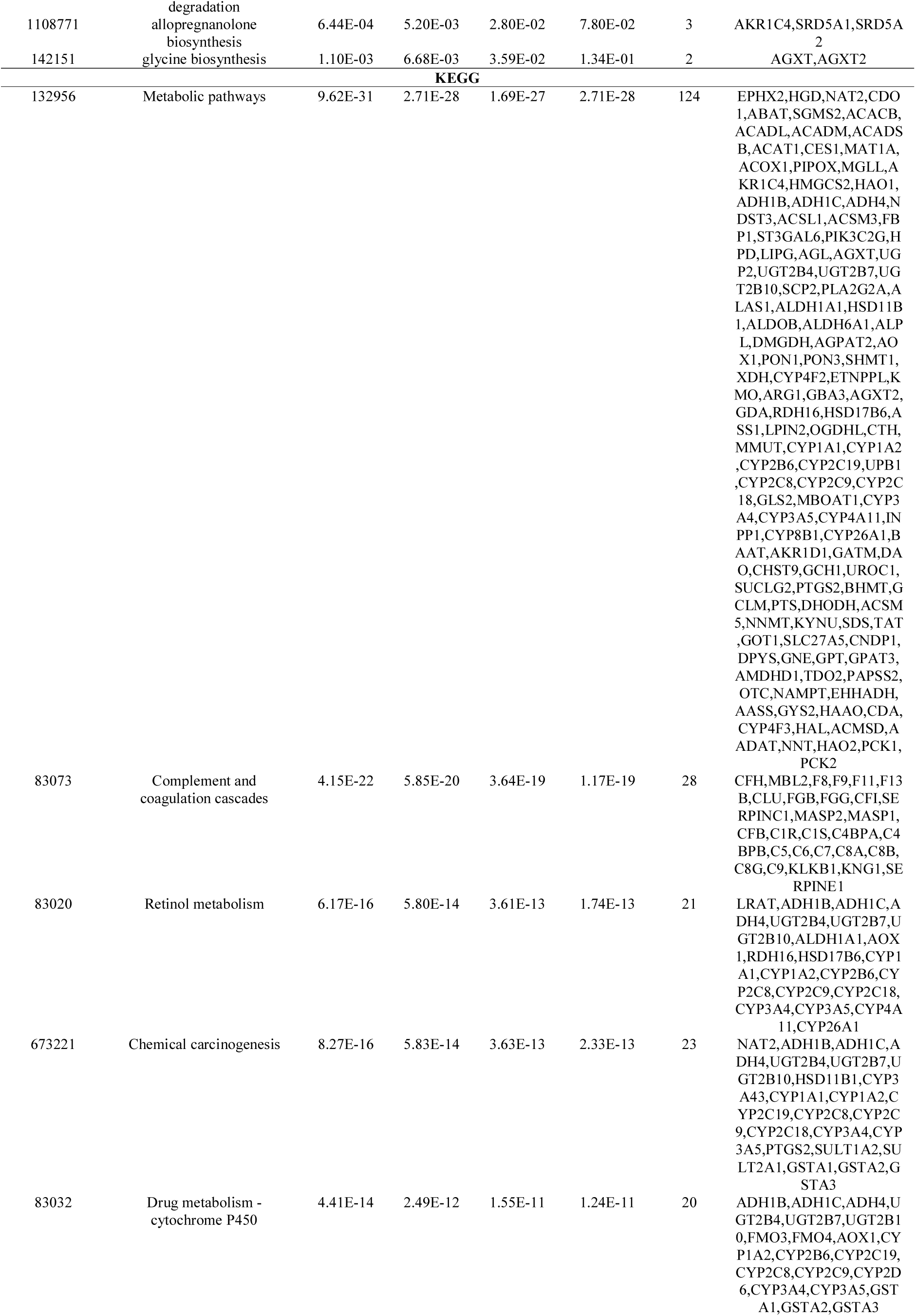
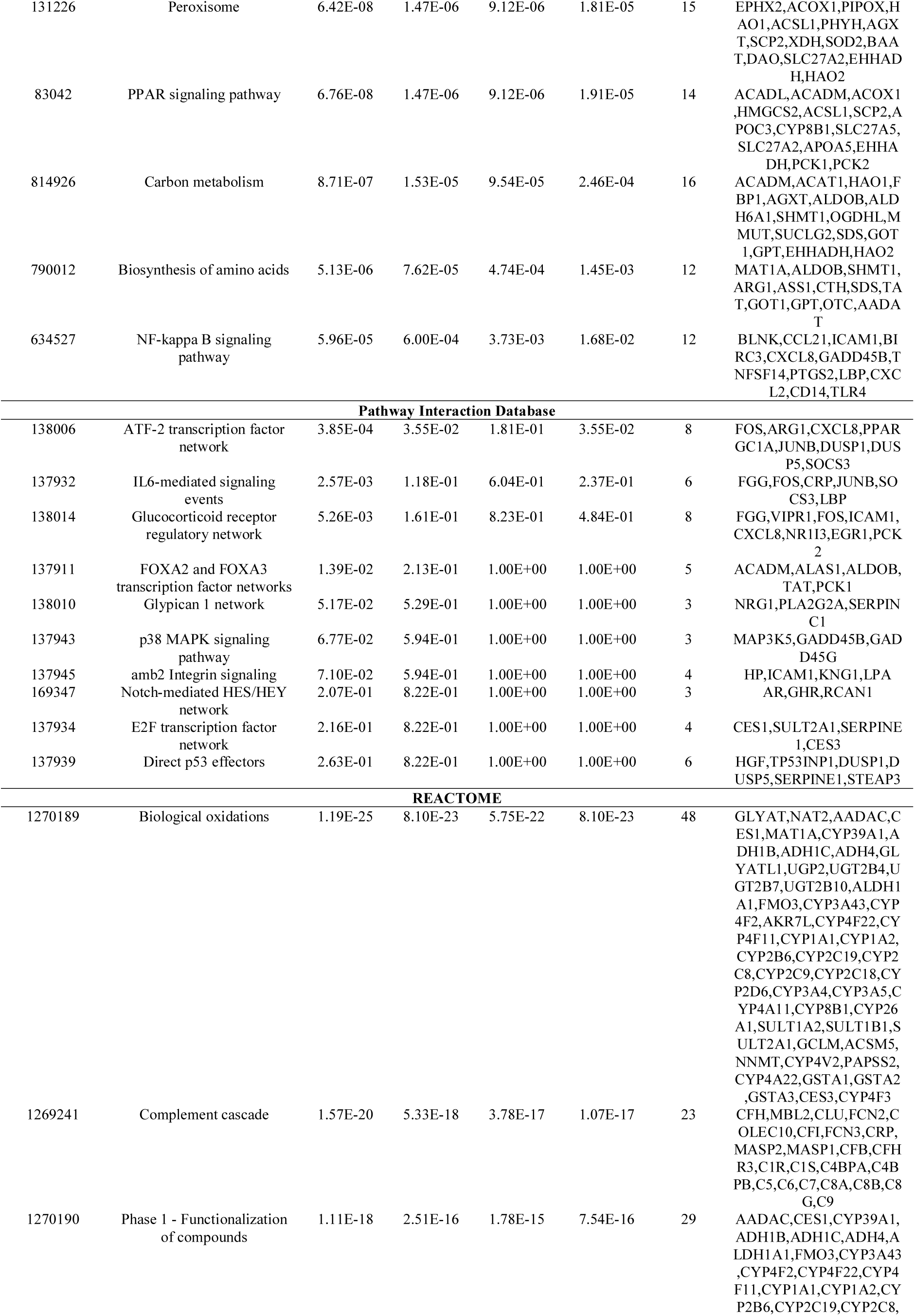
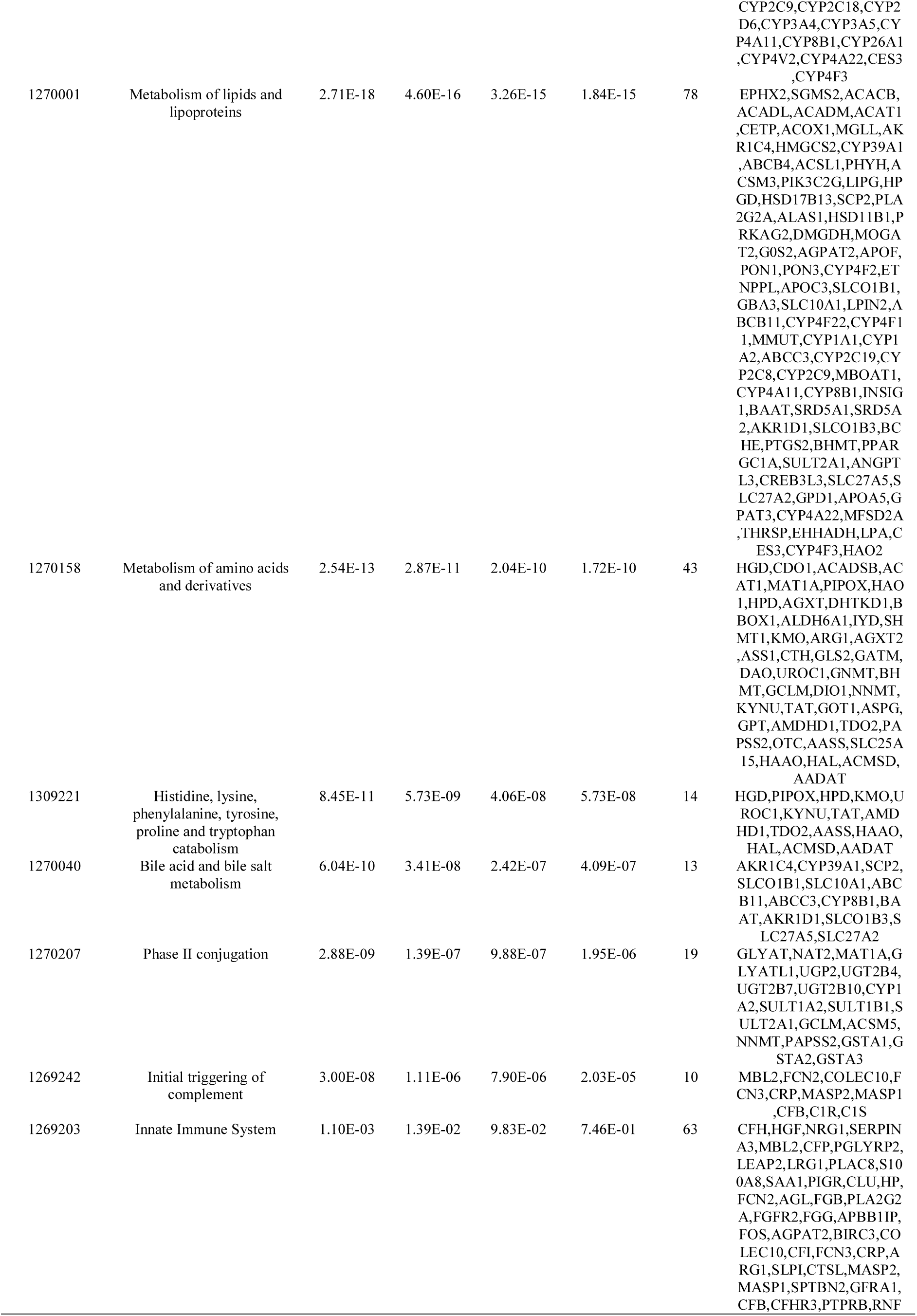
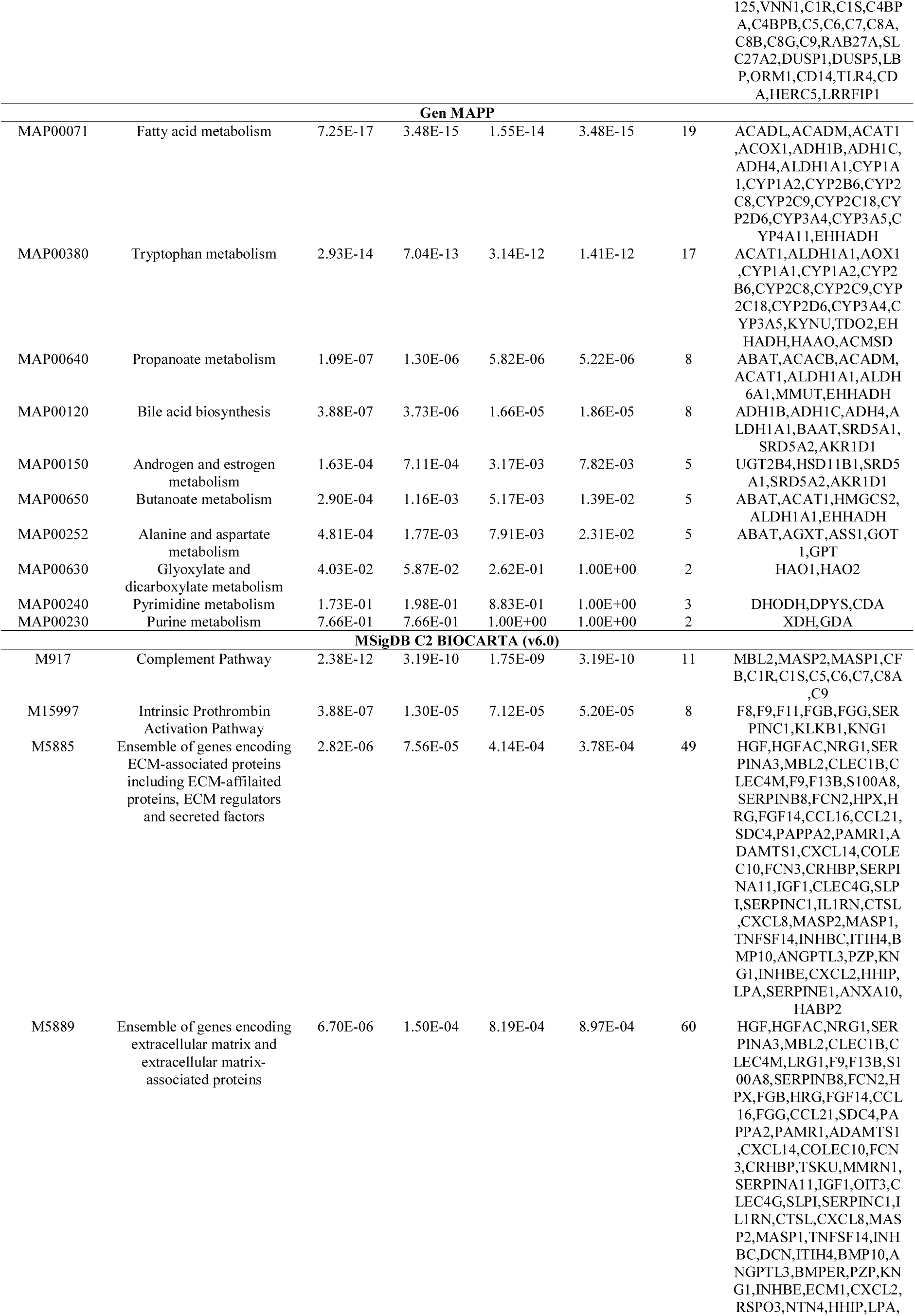
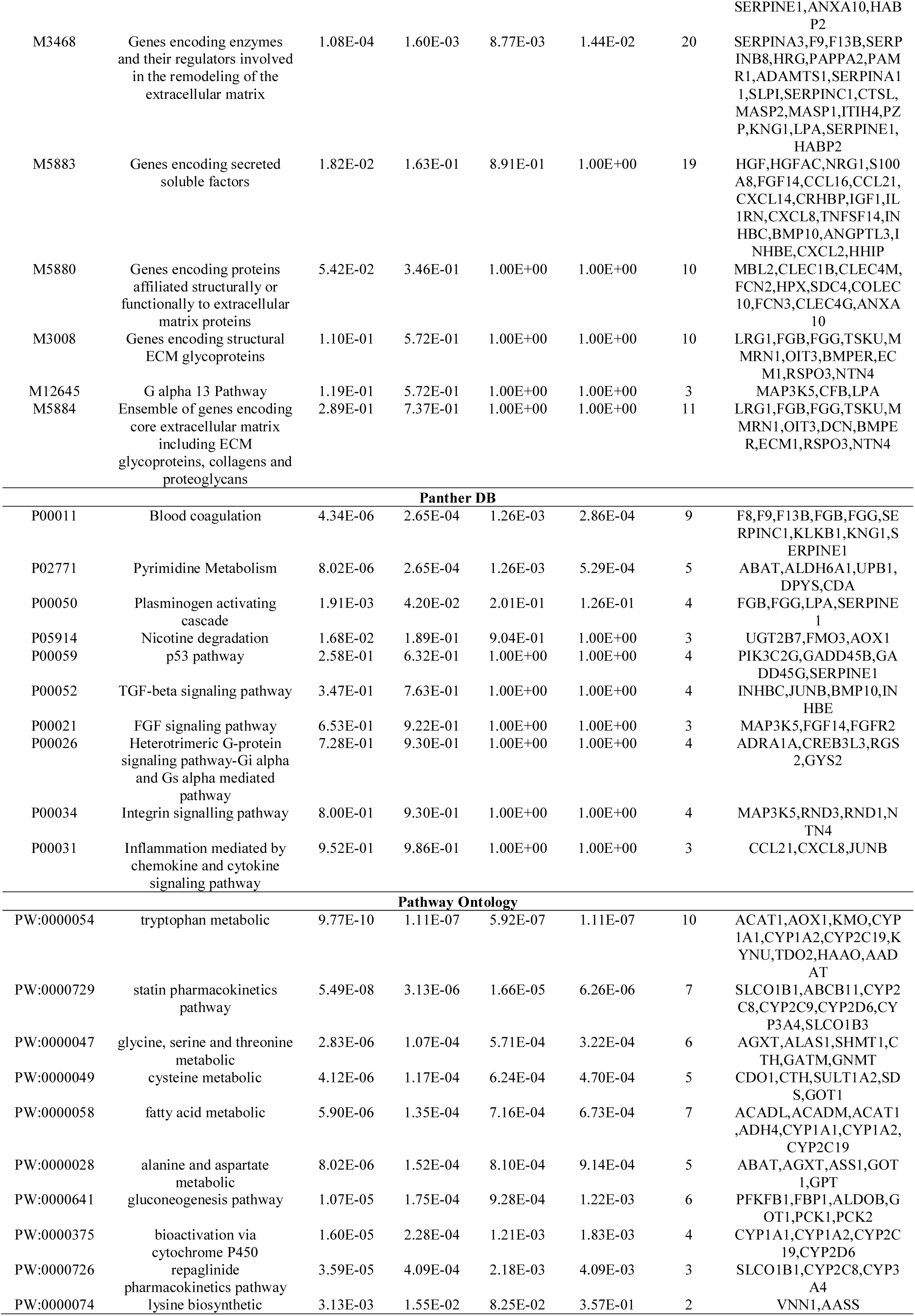
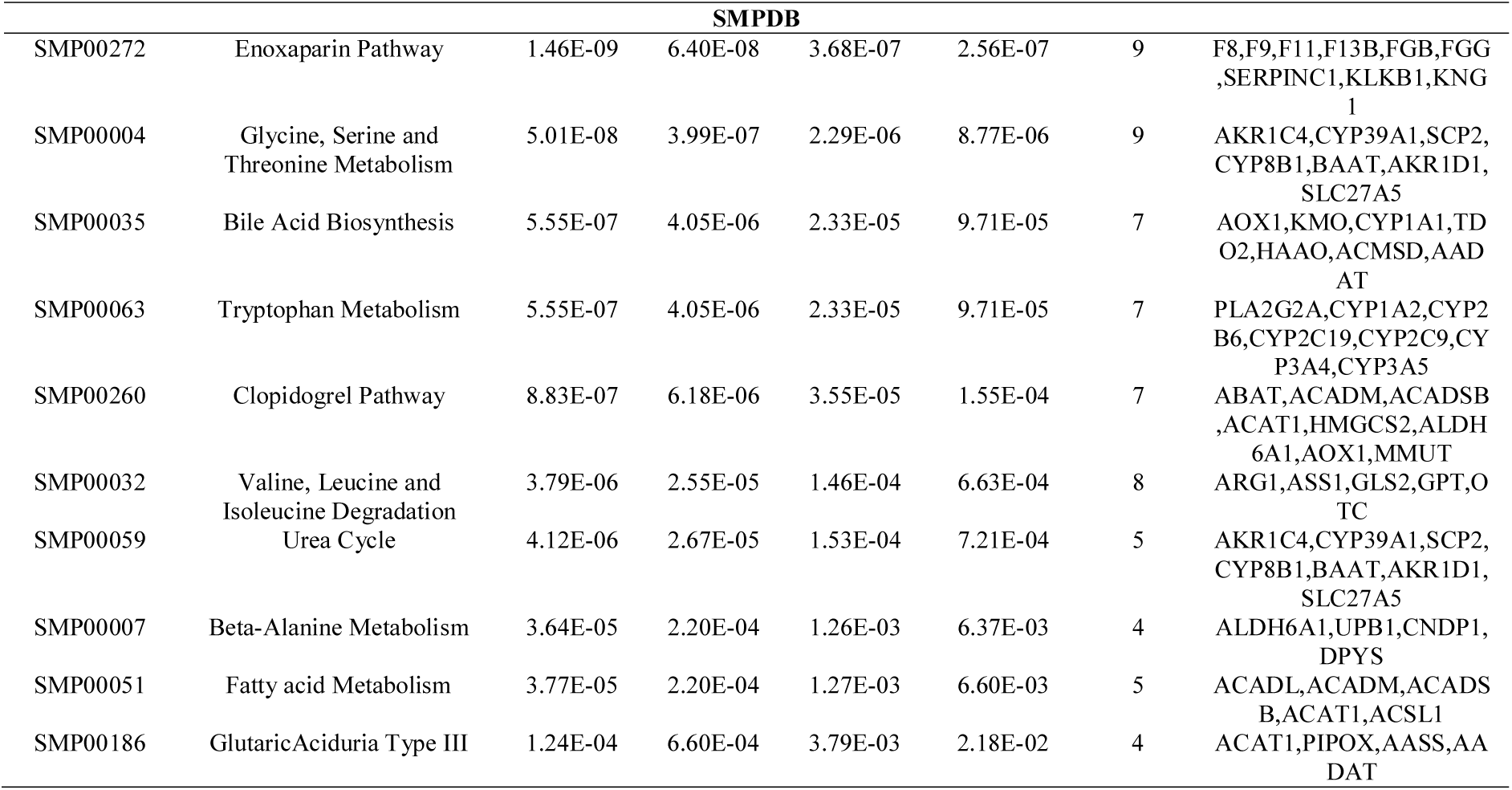
The enriched pathway terms of the down-regulated differentially expressed genes

### GO enrichment analysis of DEGs

The enriched GO terms for up and down regulated genes, which are expressed by biological process (BP), cellular component (CC) and molecular function (MF), are shown in Table 5 and Table 6. The GO functional annotation analysis of these up regulated genes revealed that (1) the BPs were mainly involved in chromosome organization and DNA conformation change, (2) the CCs of the altered genes were mainly involved in DNA packaging complex and chromosomal region, and (3) the MFs were mainly involved in DNA binding, bending and protein dimerization activity. Similarly, GO functional annotation analysis of these down regulated genes revealed that (1) the BPs were mainly involved in organic acid metabolic process and small molecule catabolic process, 2) the CCs of the altered genes were mainly involved in blood microparticle and endoplasmic reticulum membrane, and (3) the MFs were mainly involved in cofactor binding and oxidoreductase activity.

**Table 5.**
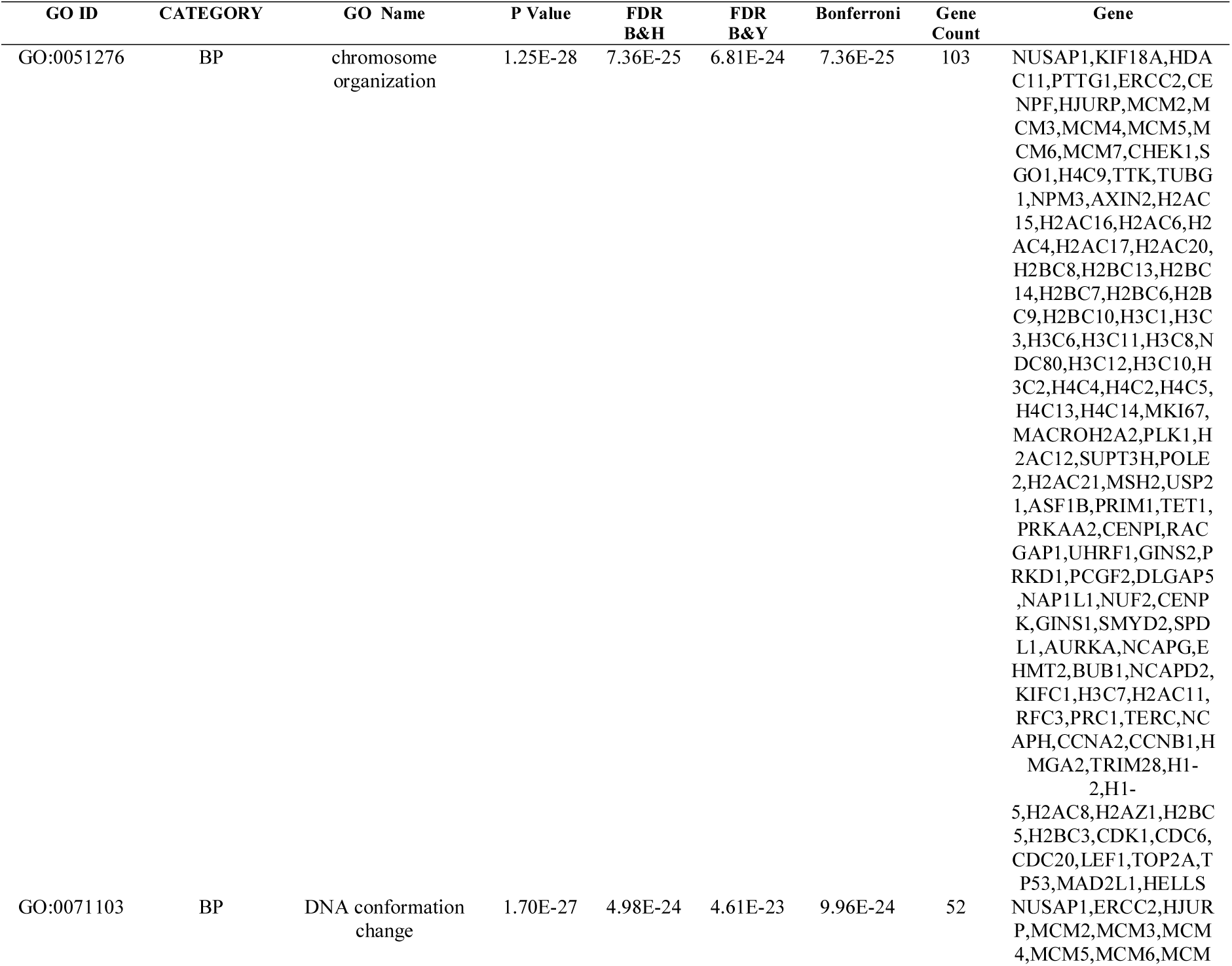
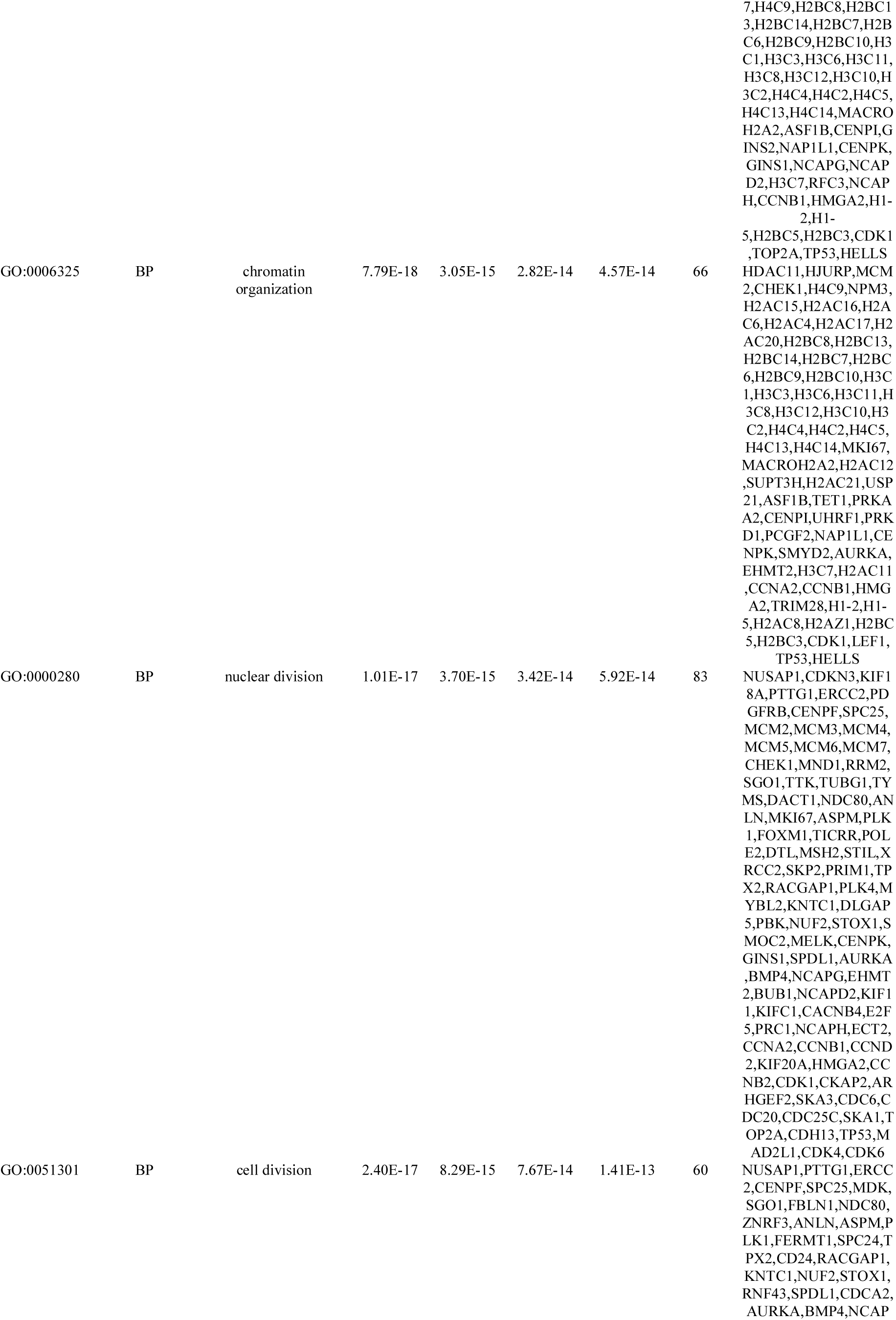
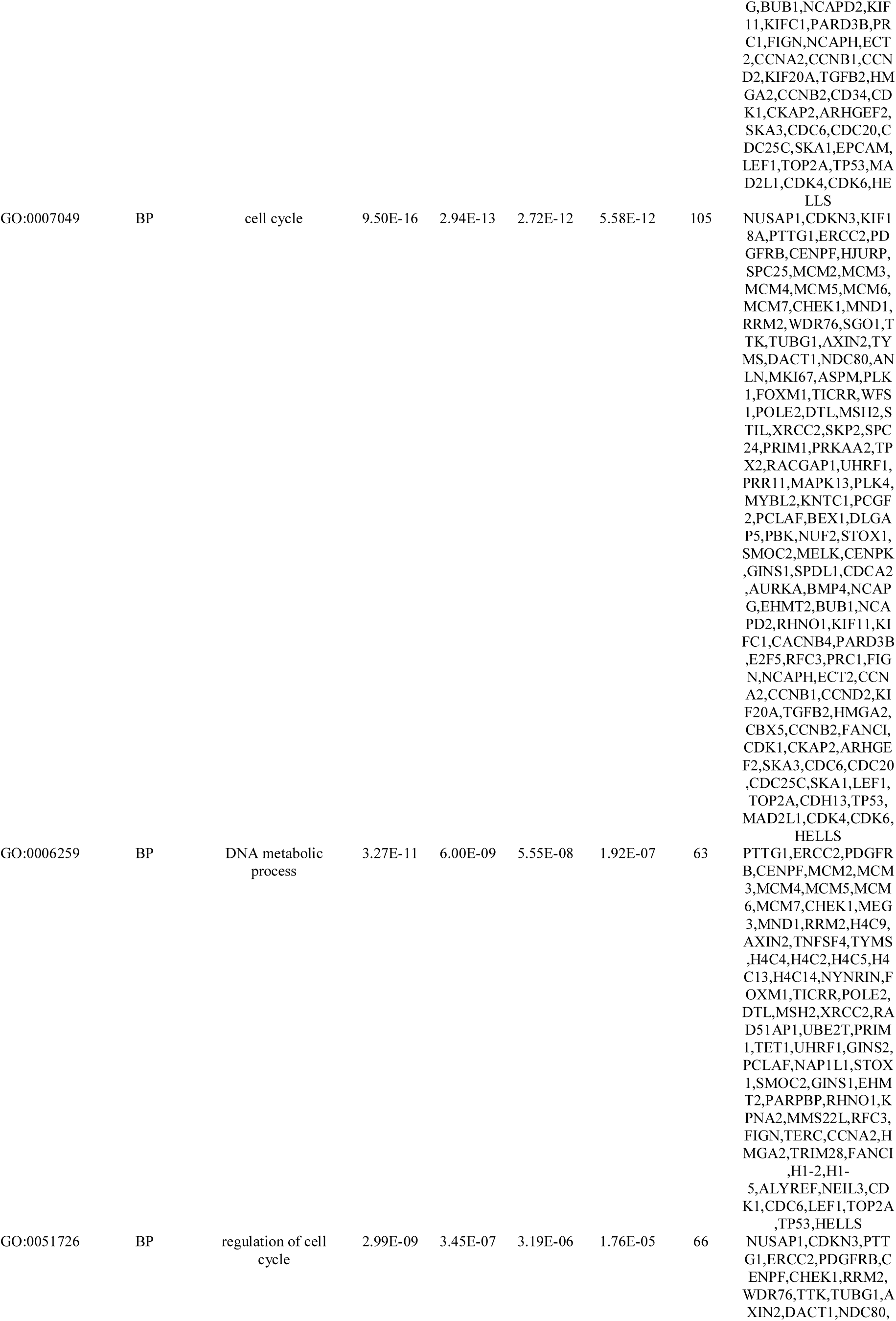
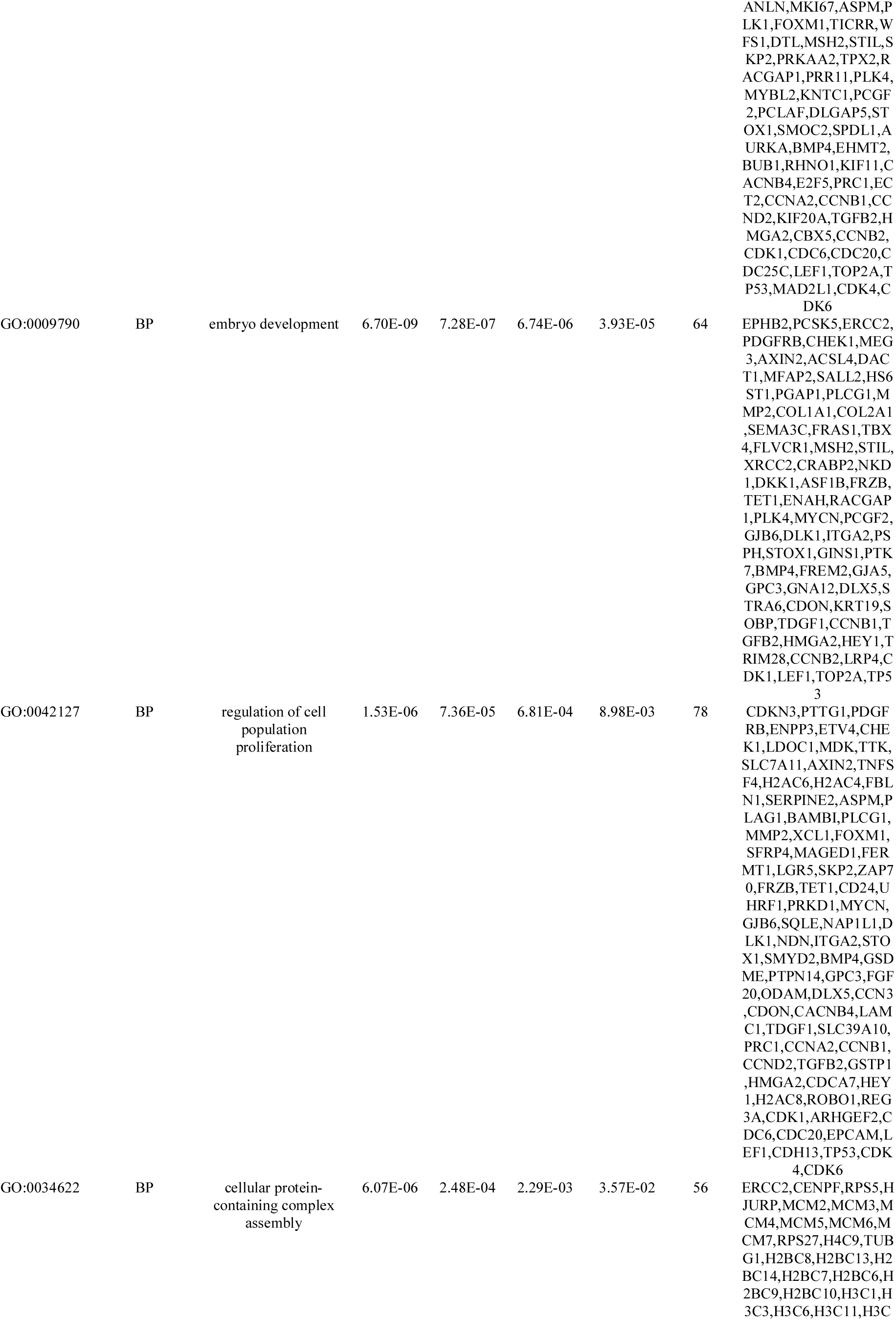
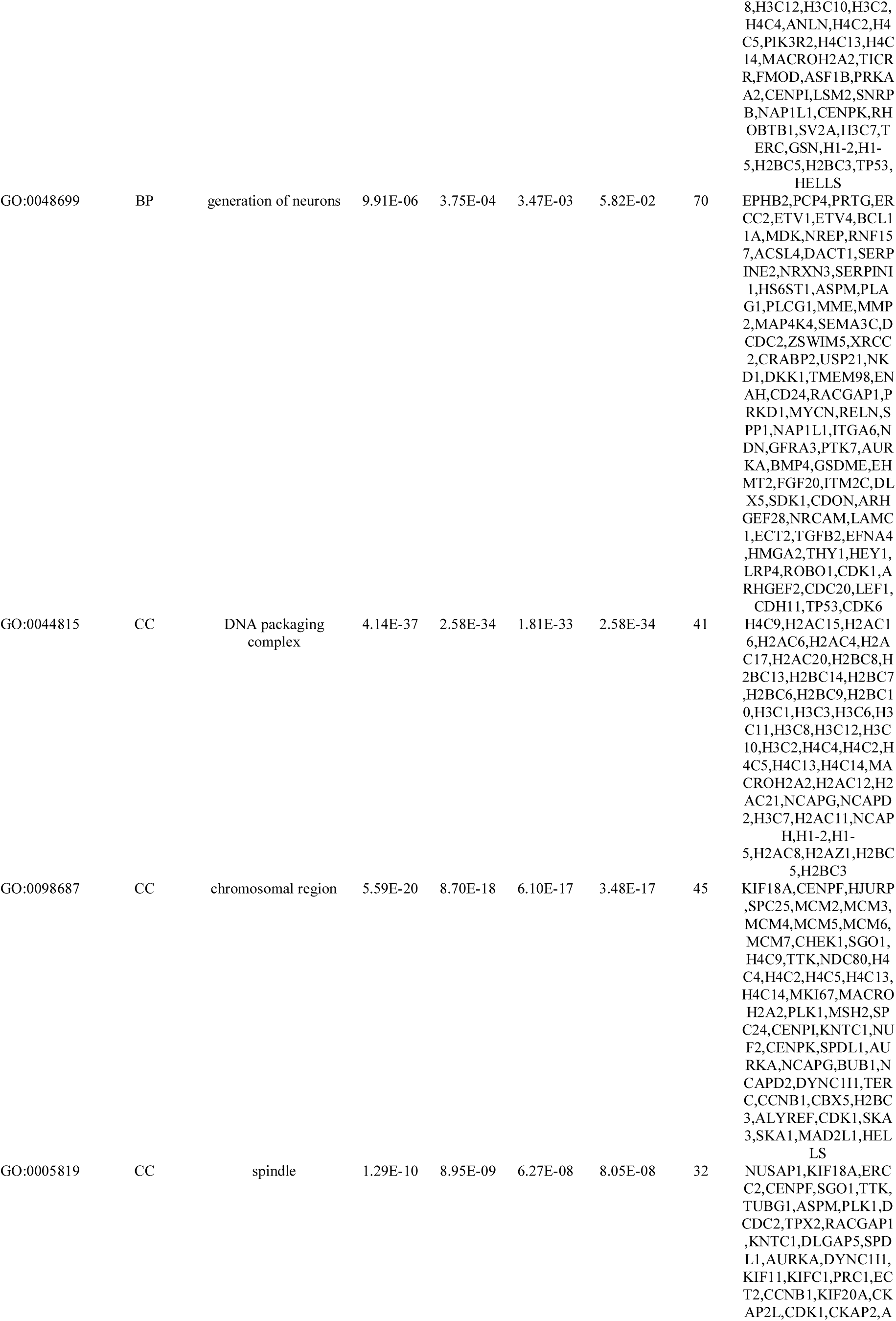
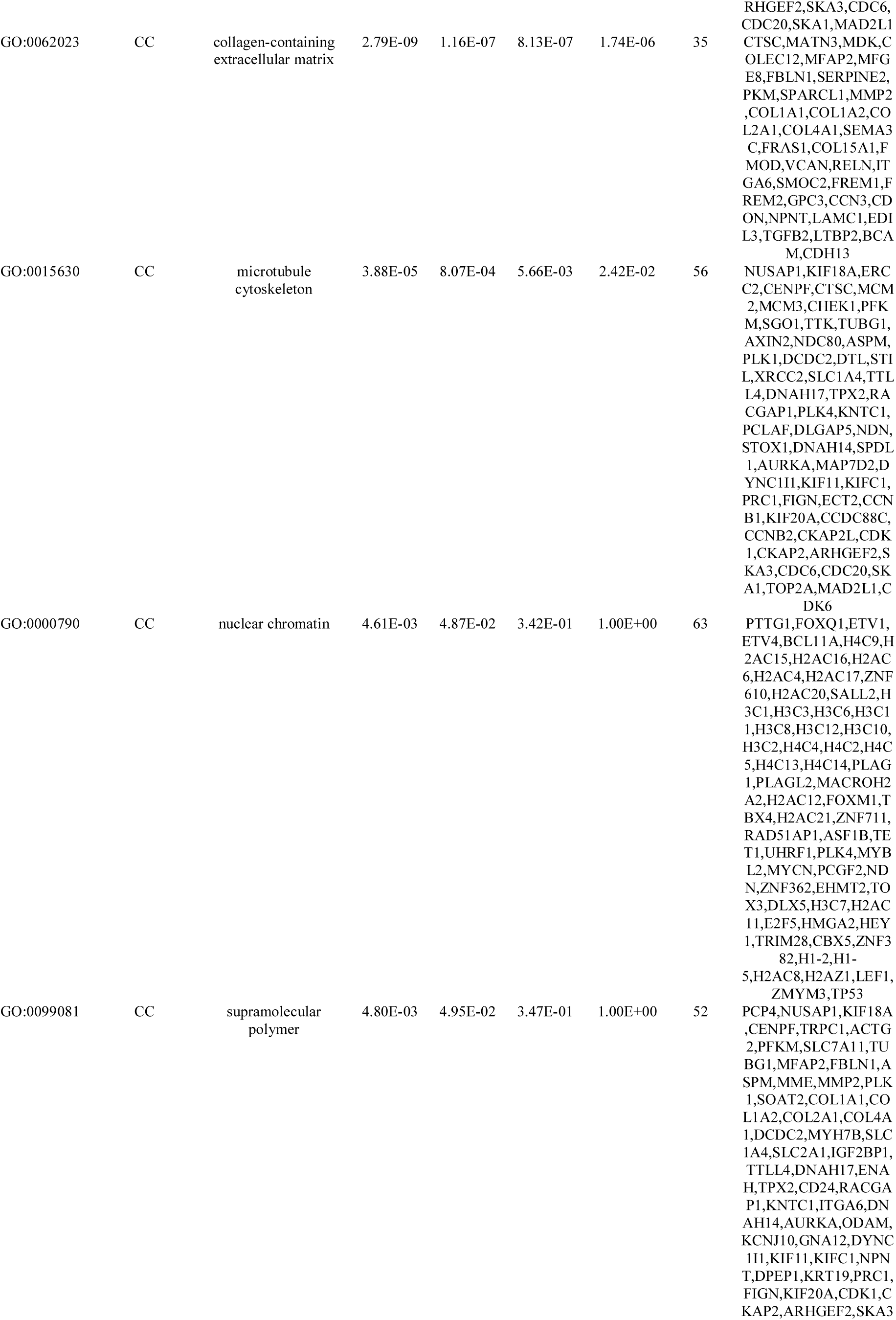
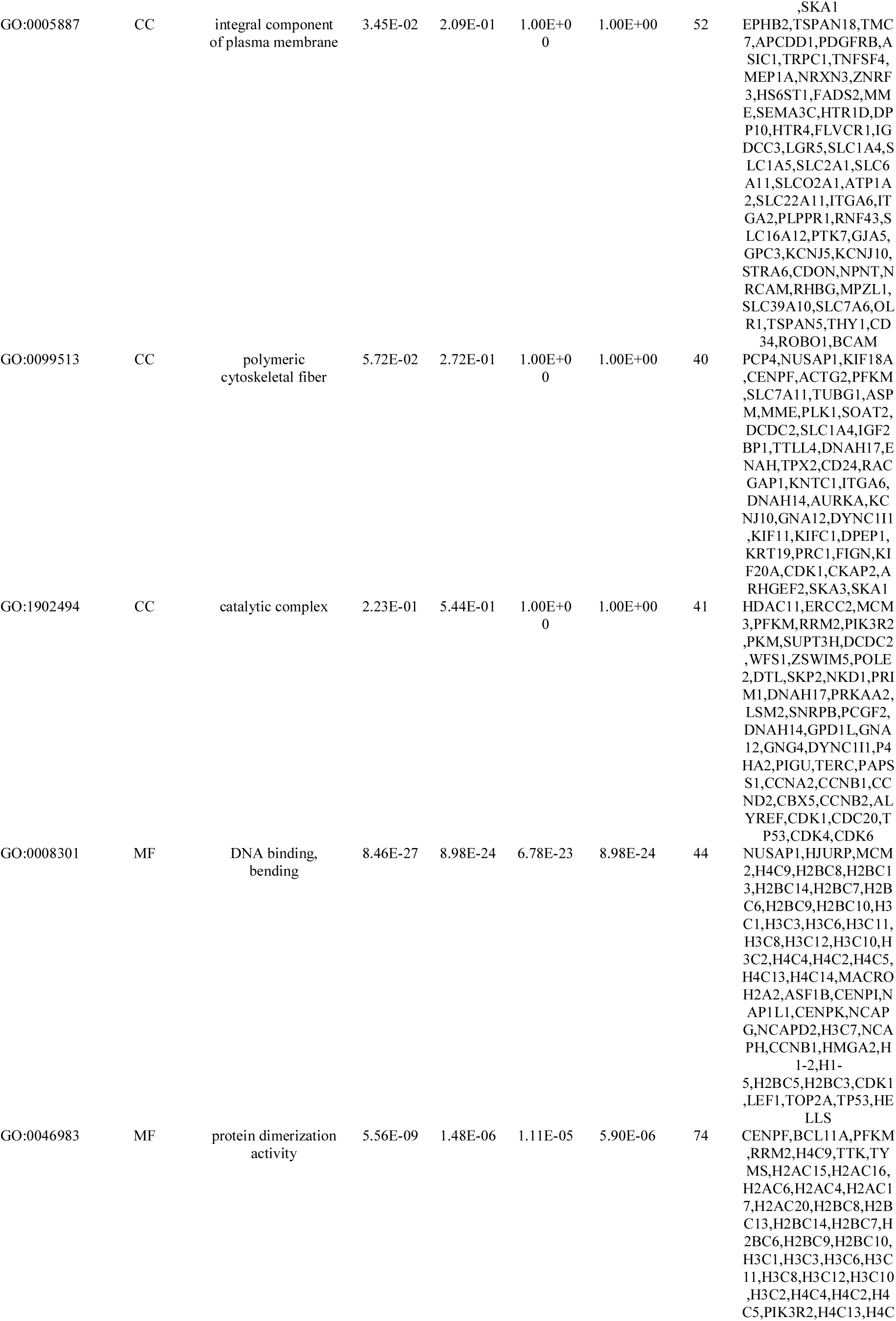
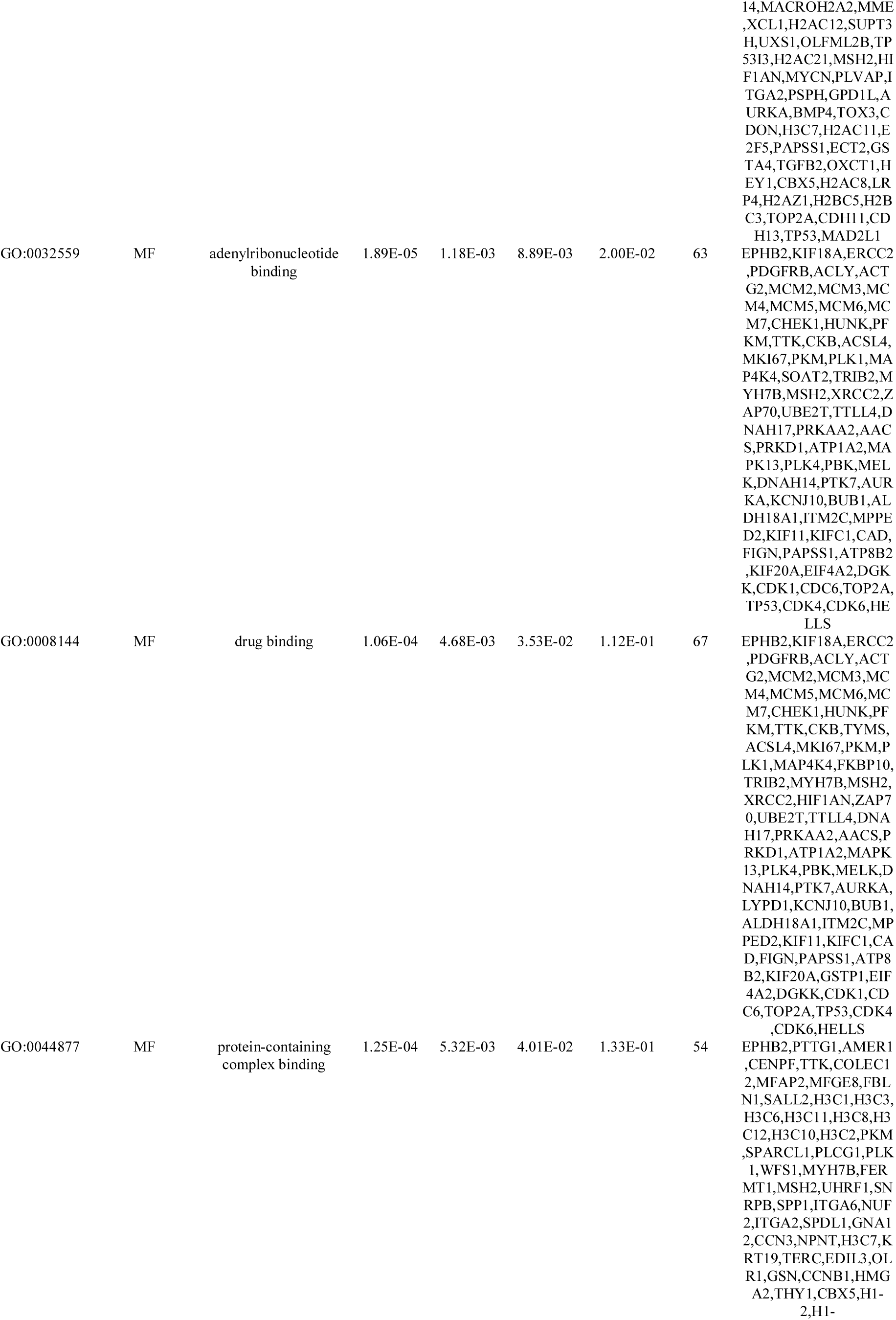
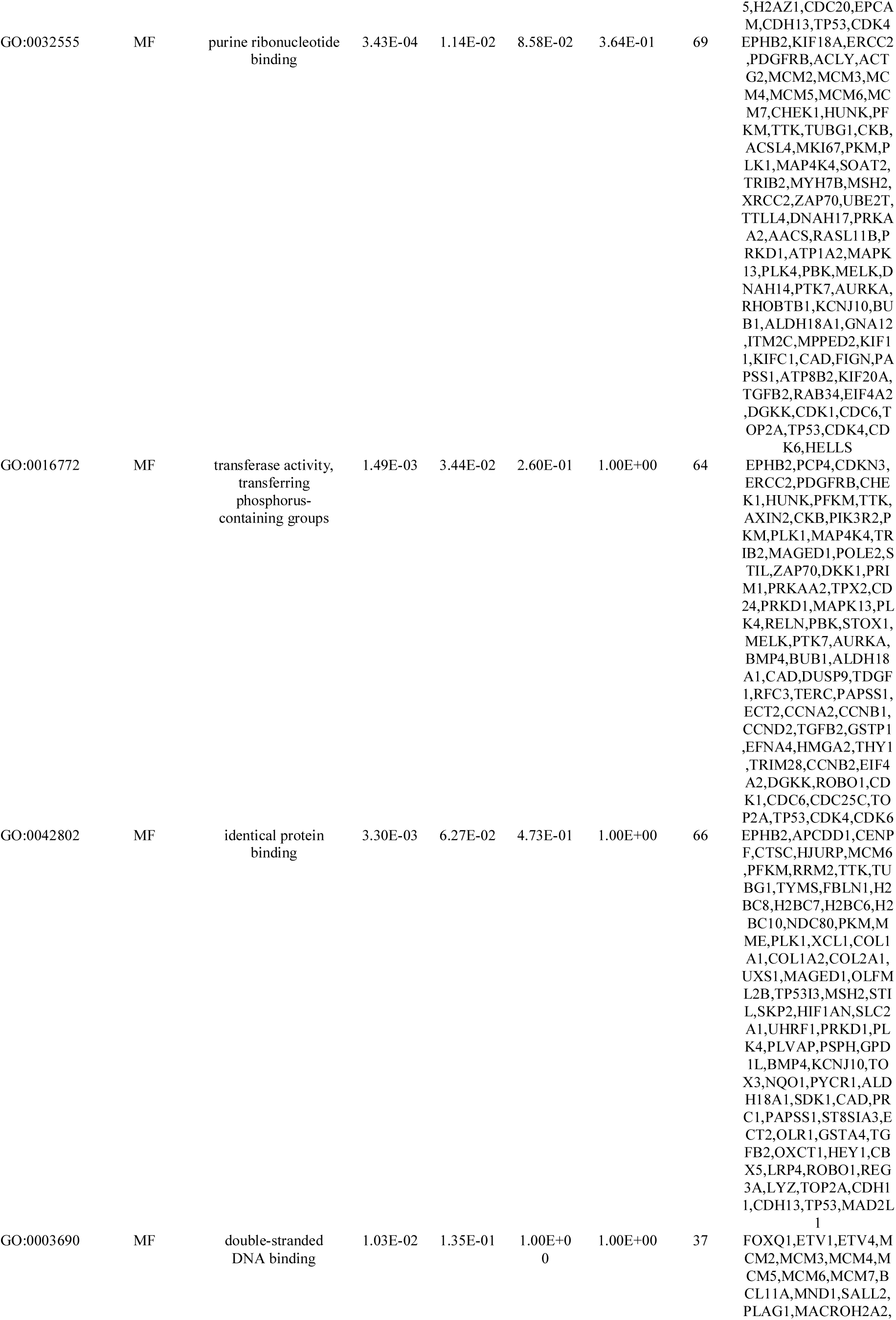
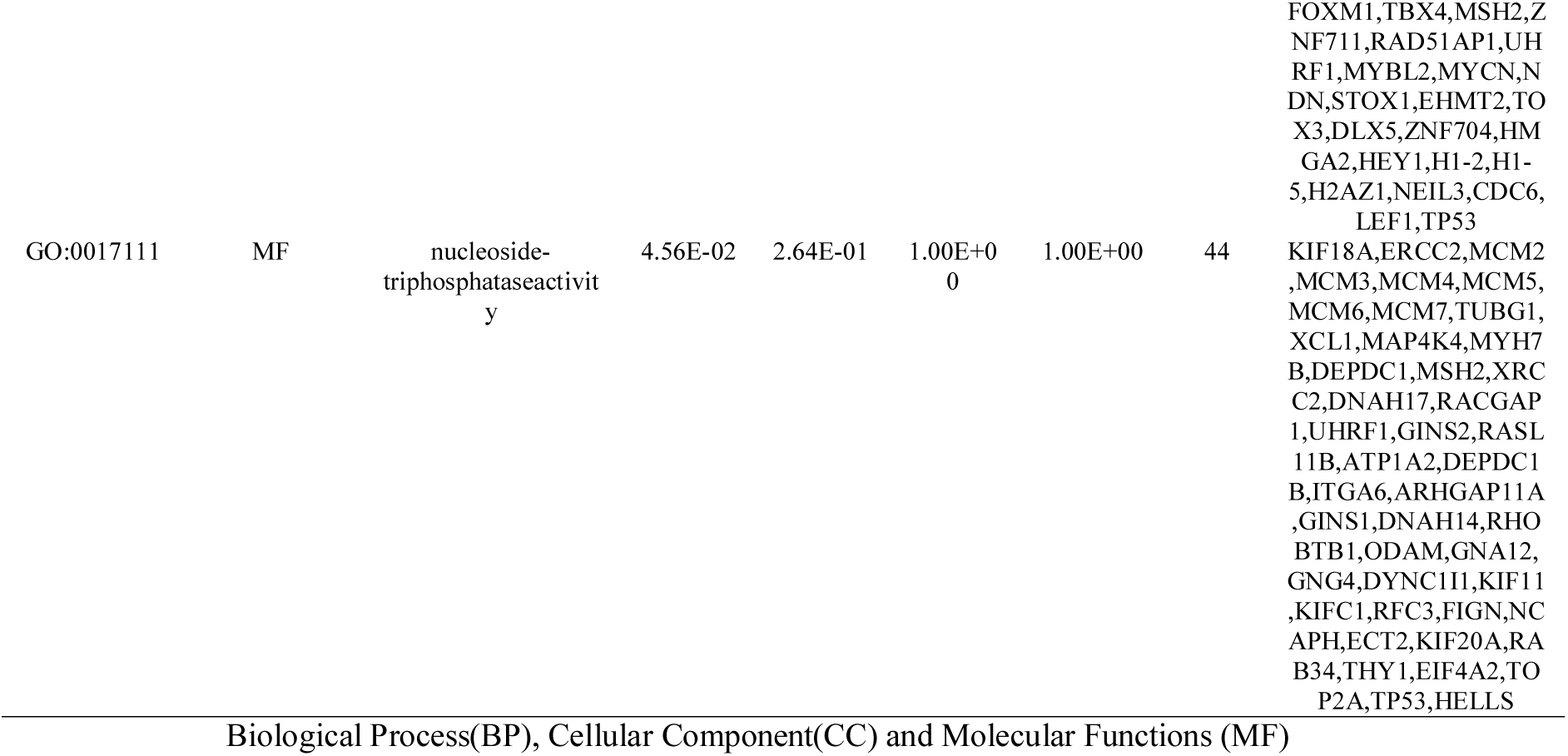
The enriched GO terms of the up regulated differentially expressed genes

**Table 6.**
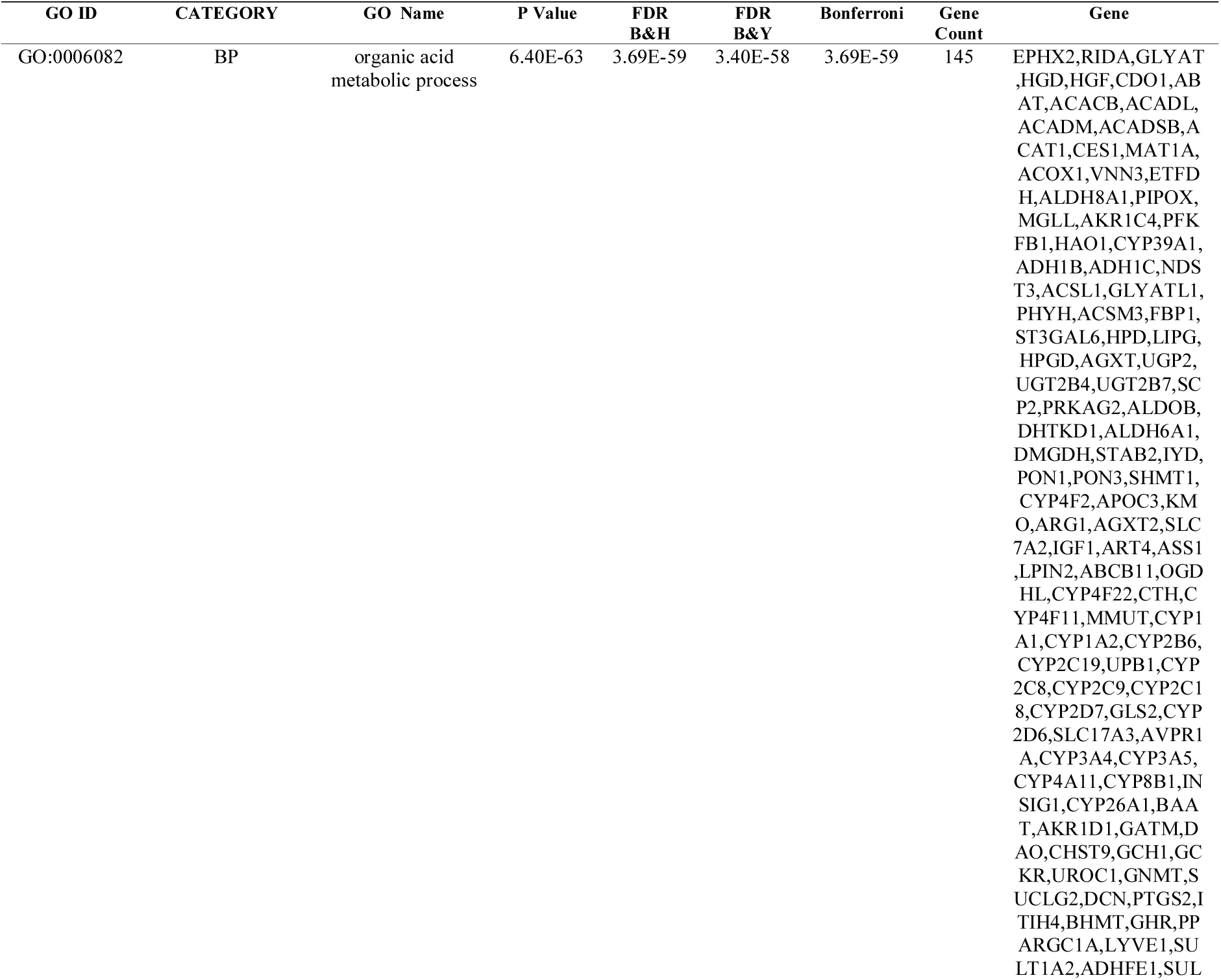
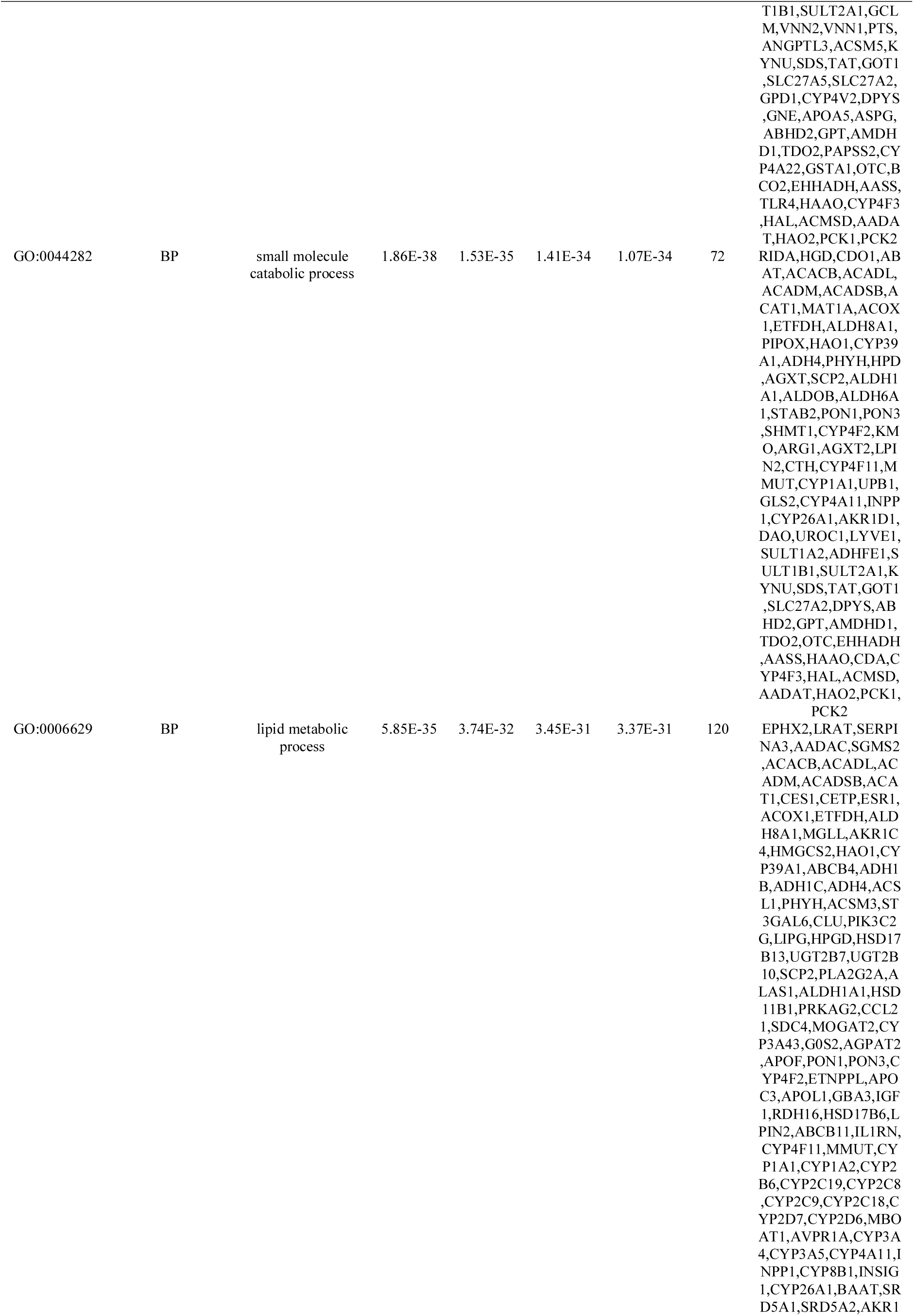
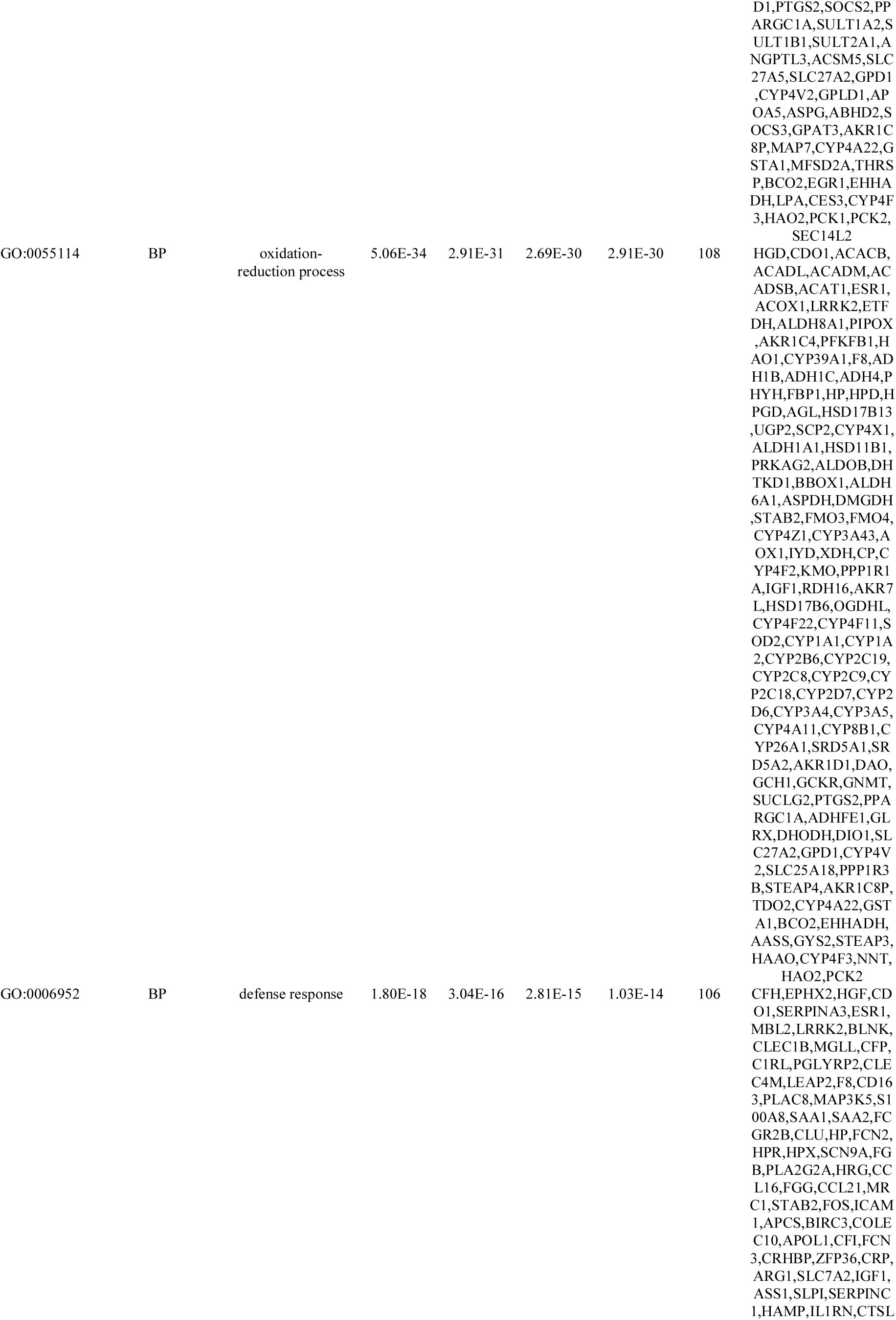
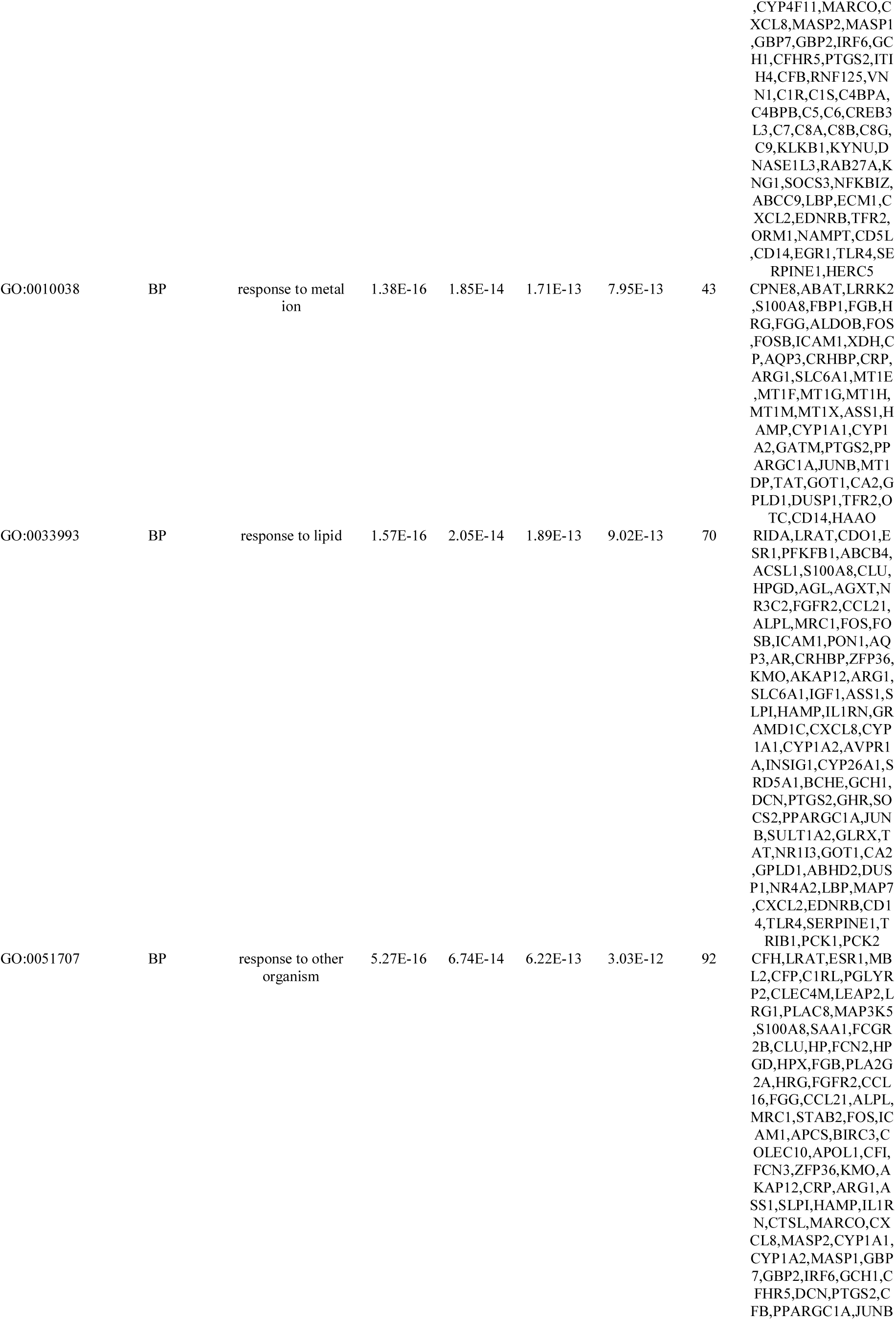
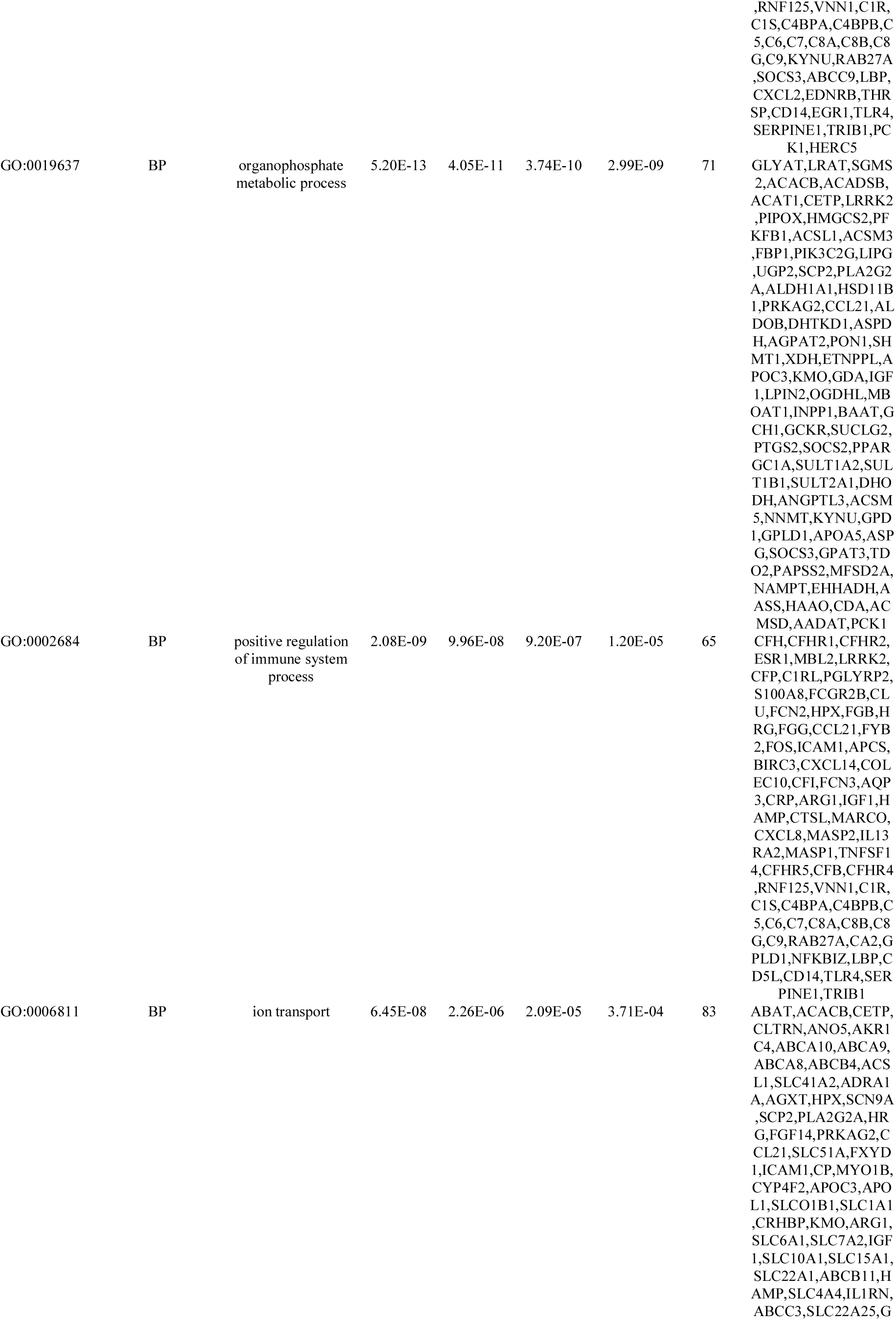
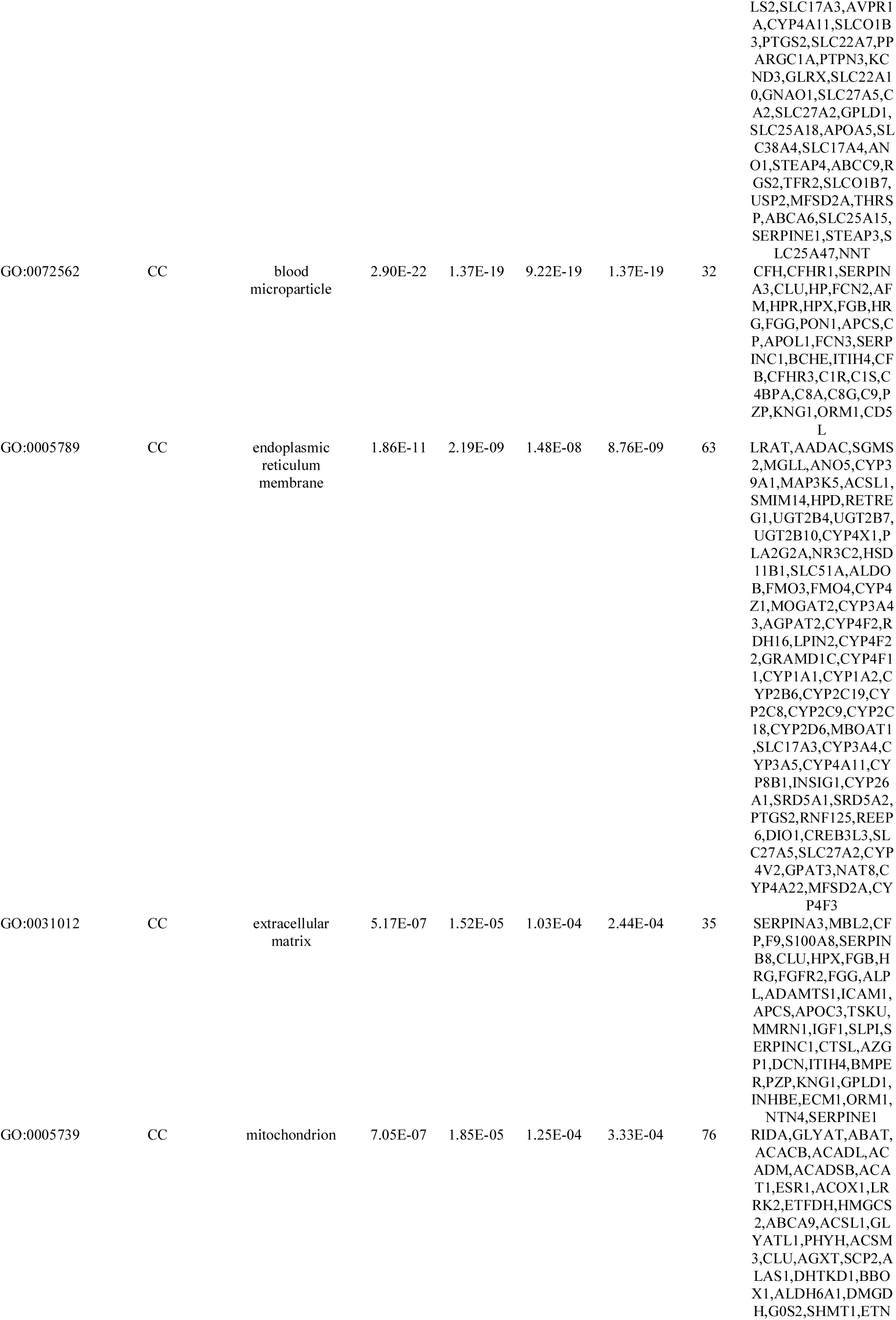
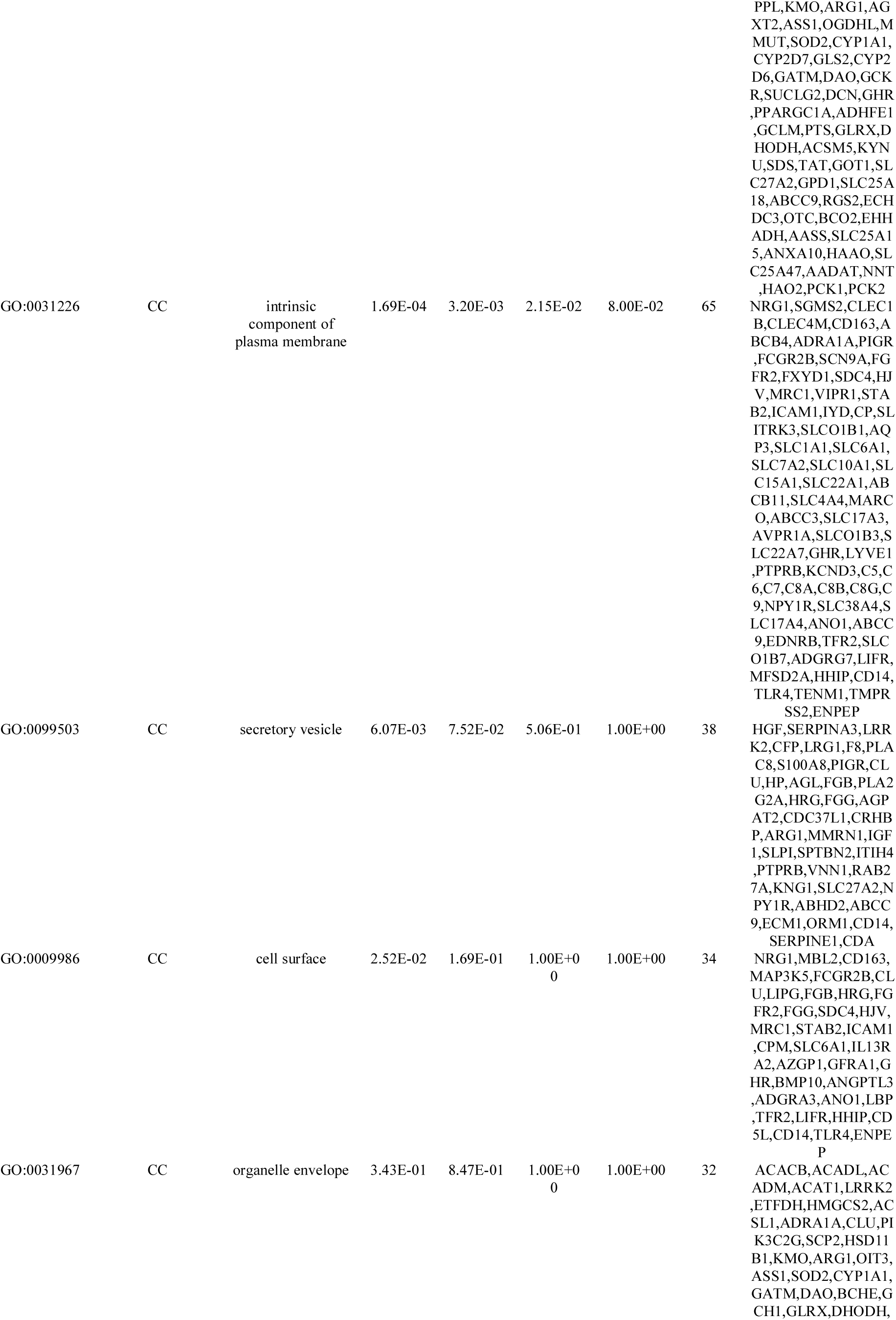
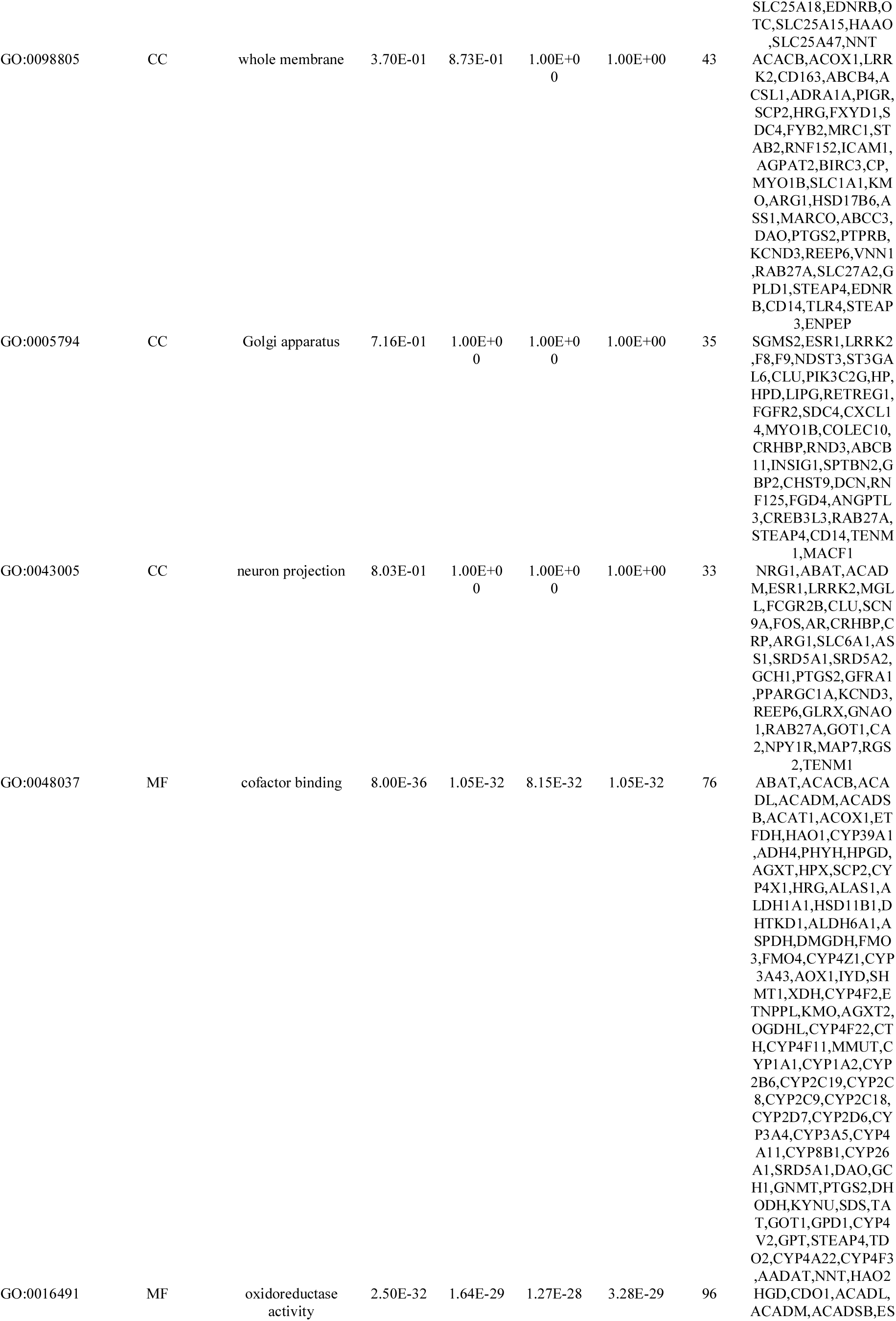
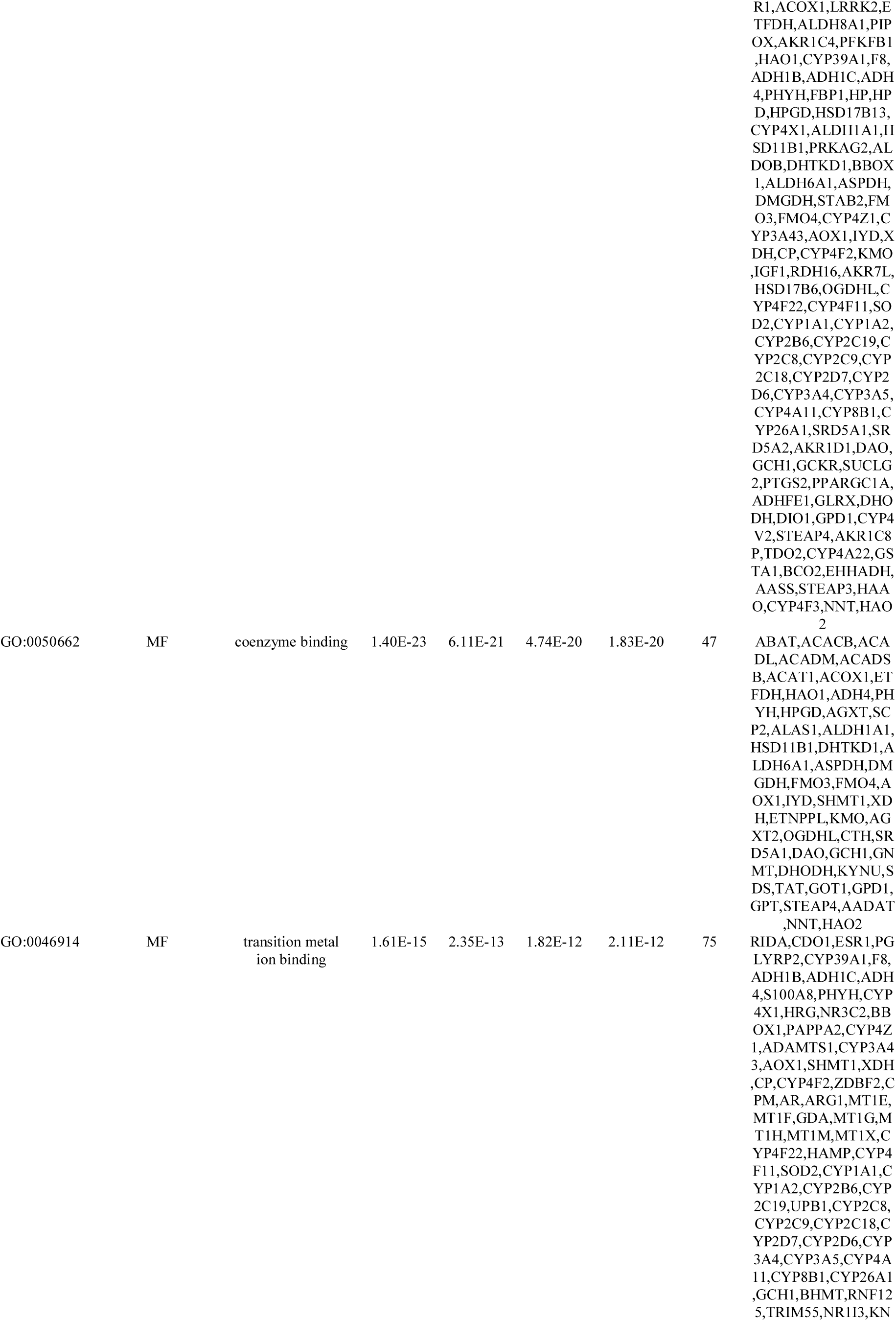
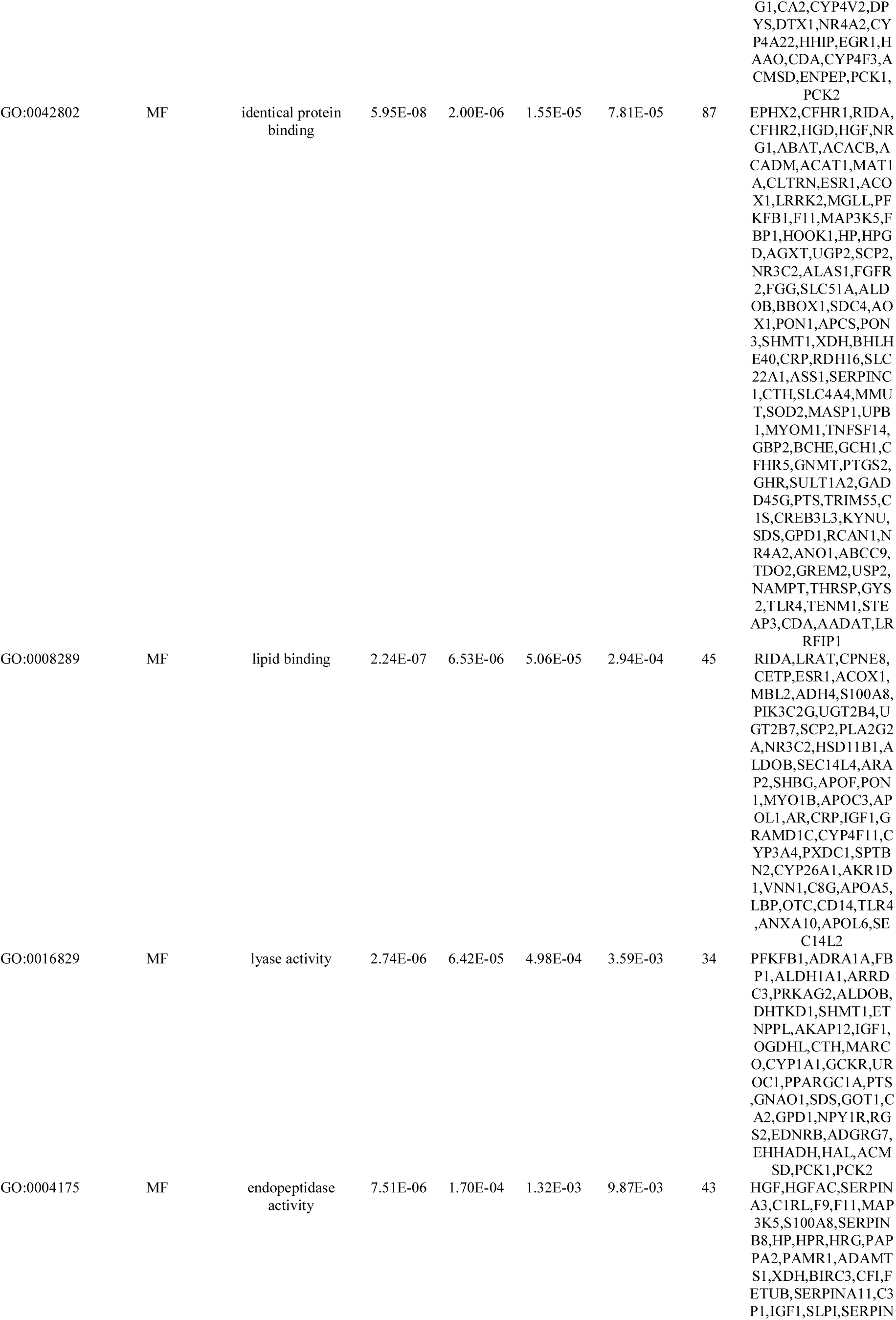
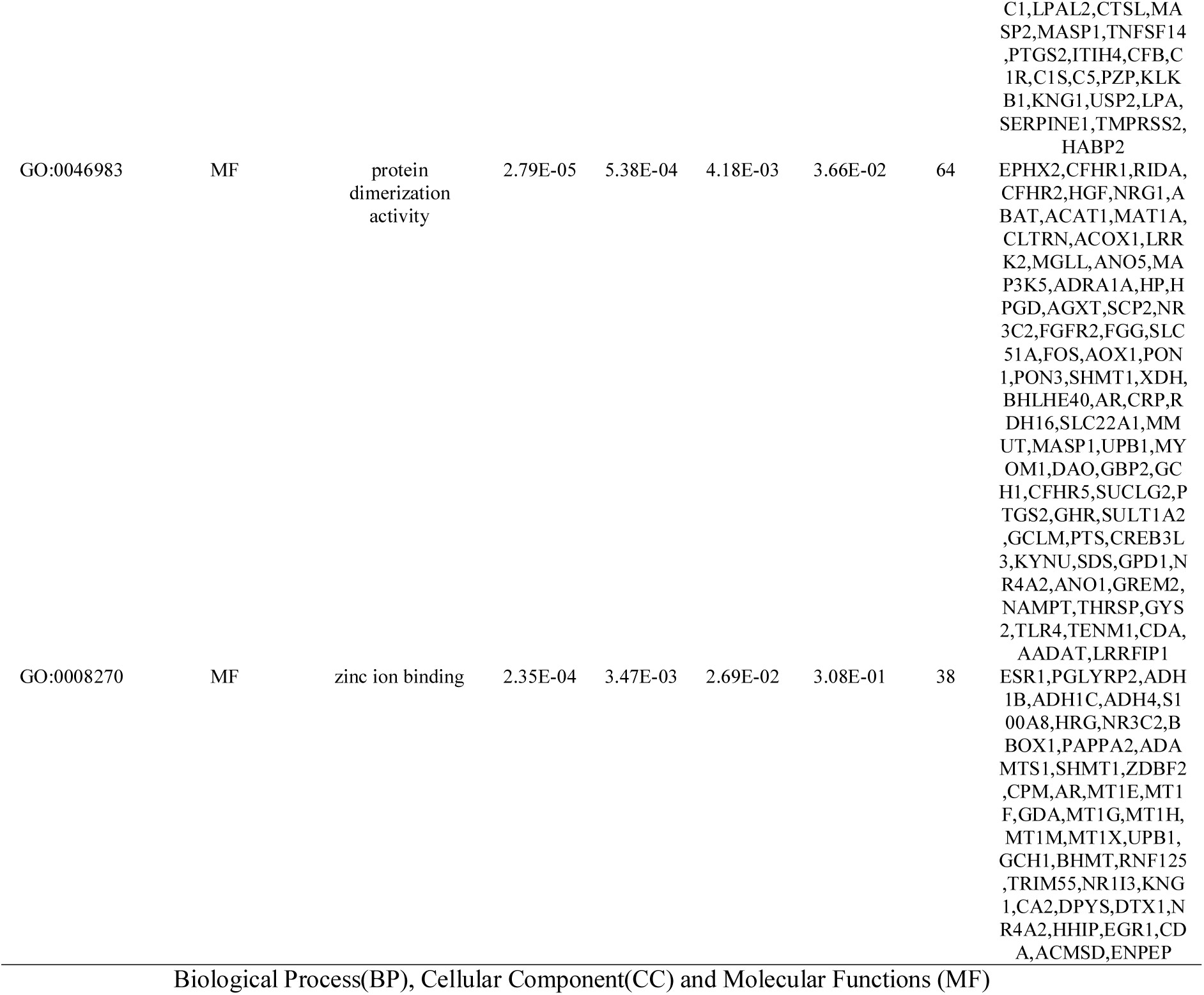
The enriched GO terms of the down regulated differentially expressed genes

### Construction of the PPI network and module analysis

To further explore the biological role of overlapping up and down regulated genes, a PPI network was created using the IID database. The PPI network for up regulated genes consisted of 4396 nodes and 7665 interactions (Fig. 5). The 12 up regulated hub genes TP53, TCTN2, PLK1, AURKA, CDK1, ANLN, TRIM28, BMP4, PLPPR1, FOXRED2, SERPINE2 and EFNA4 were screened with the highest node degree distribution, betweenness centrality, stress centrality, closeness centrality and lowest clustring coefficient using Cytoscape software (Table 6), and scatter plot along with statistical results for node degree distribution, betweenness centrality, stress centrality, closeness centrality and clustring coefficient are presented in Fig. 6A - 6E. These up regulated hub genes were enriched in cell cycle, chromosome organization, metabolism of proteins, gene expression, nuclear division, DNA metabolic process, pathways in cancer, integral component of plasma membrane, ensemble of genes encoding extracellular matrix and extracellular matrix-associated proteins, and generation of neurons. Similarly, PPI network for down regulated genes consisted of 5582 nodes and 10377 interactions (Fig. 7). The 12 down regulated hub genes ESR1, LRRK2, AR, FOS, ICAM1, CLU, MYO1B, NPY1R, SLC22A1, FGGY, SLC10A1 and CYP4X1 were screened with the highest node degree distribution, betweenness centrality, stress centrality, closeness centrality and lowest clustring coefficient using Cytoscape software (Table 6), and scatter plot along with statistical results for node degree distribution, betweenness centrality, stress centrality, closeness centrality and clustring coefficient are presented in Fig. 8A - 8E. These down regulated hub genes were enriched in lipid metabolic process, oxidation-reduction process, response to lipid, ATF-2 transcription factor network, NF-kappa B signaling pathway, complement and coagulation cascades, ion transport, intrinsic component of plasma membrane, identical protein binding, metabolism of lipids and lipoproteins, and superpathway of tryptophan utilization.

**Fig. 5.**
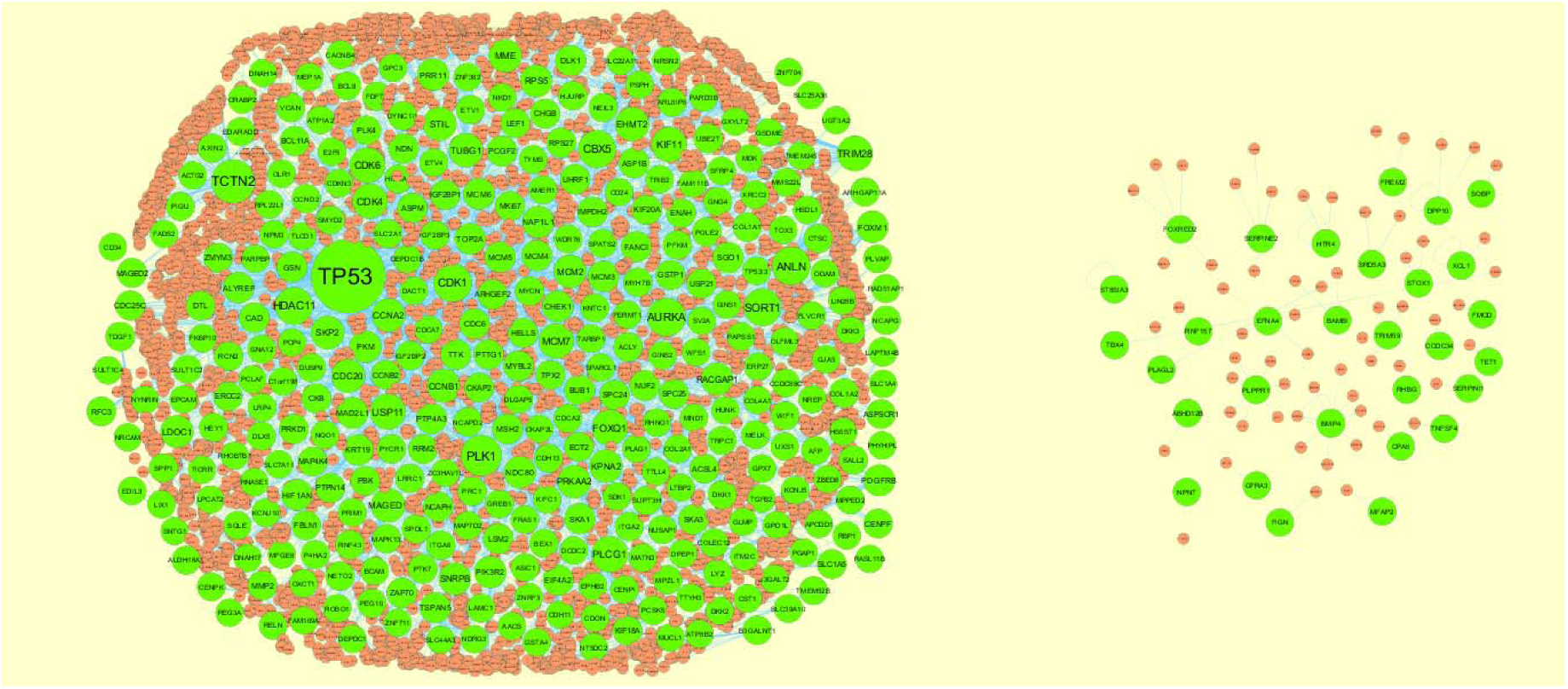
Protein–protein interaction network of up regulated genes. Green nodes denotes up regulated genes.

**Fig. 6.**
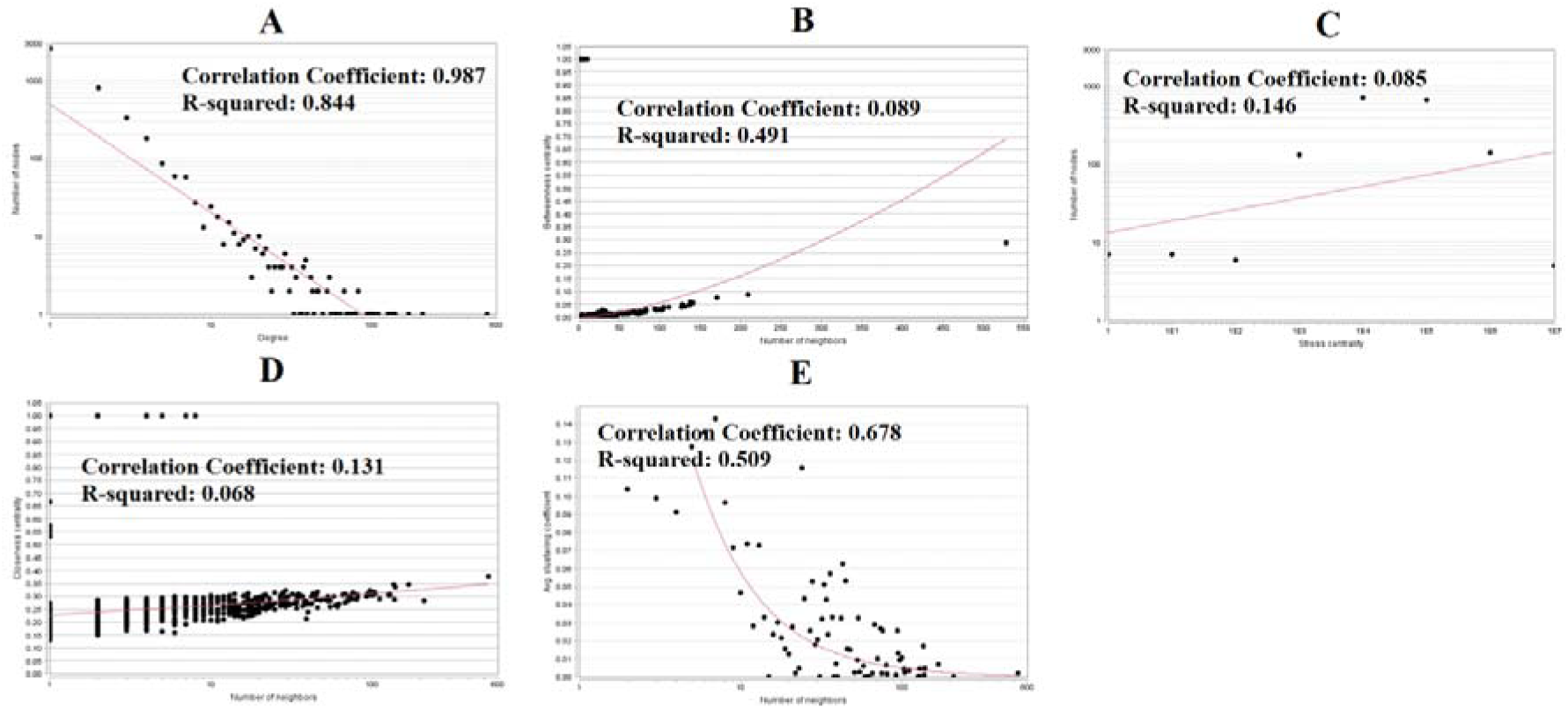
Scatter plot for up regulated genes. (A- Node degree; B- Betweenness centrality; C- Stress centrality; D-Closeness centrality; E- Clustering coefficient)

**Fig. 7.**
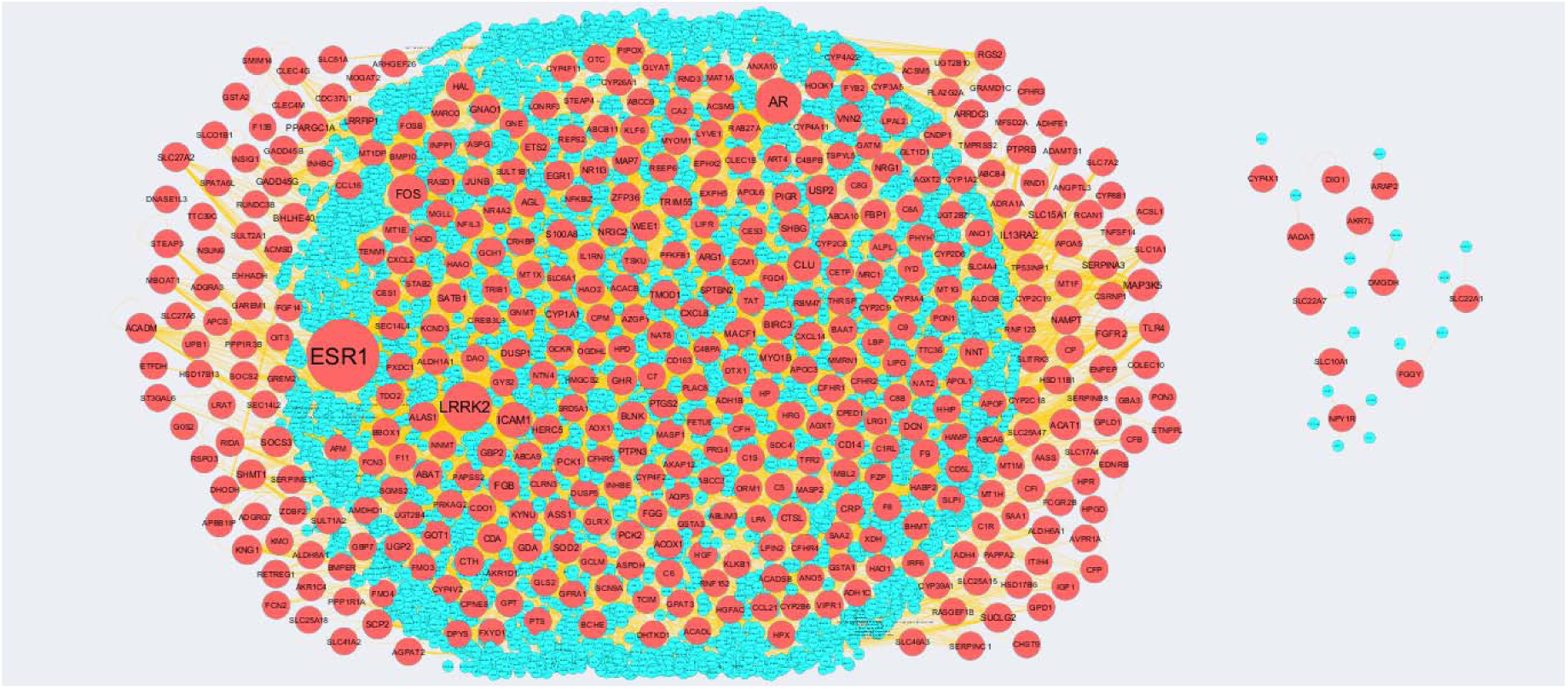
Protein–protein interaction network of down regulated genes. Red nodes denotes down regulated genes.

**Fig. 8.**
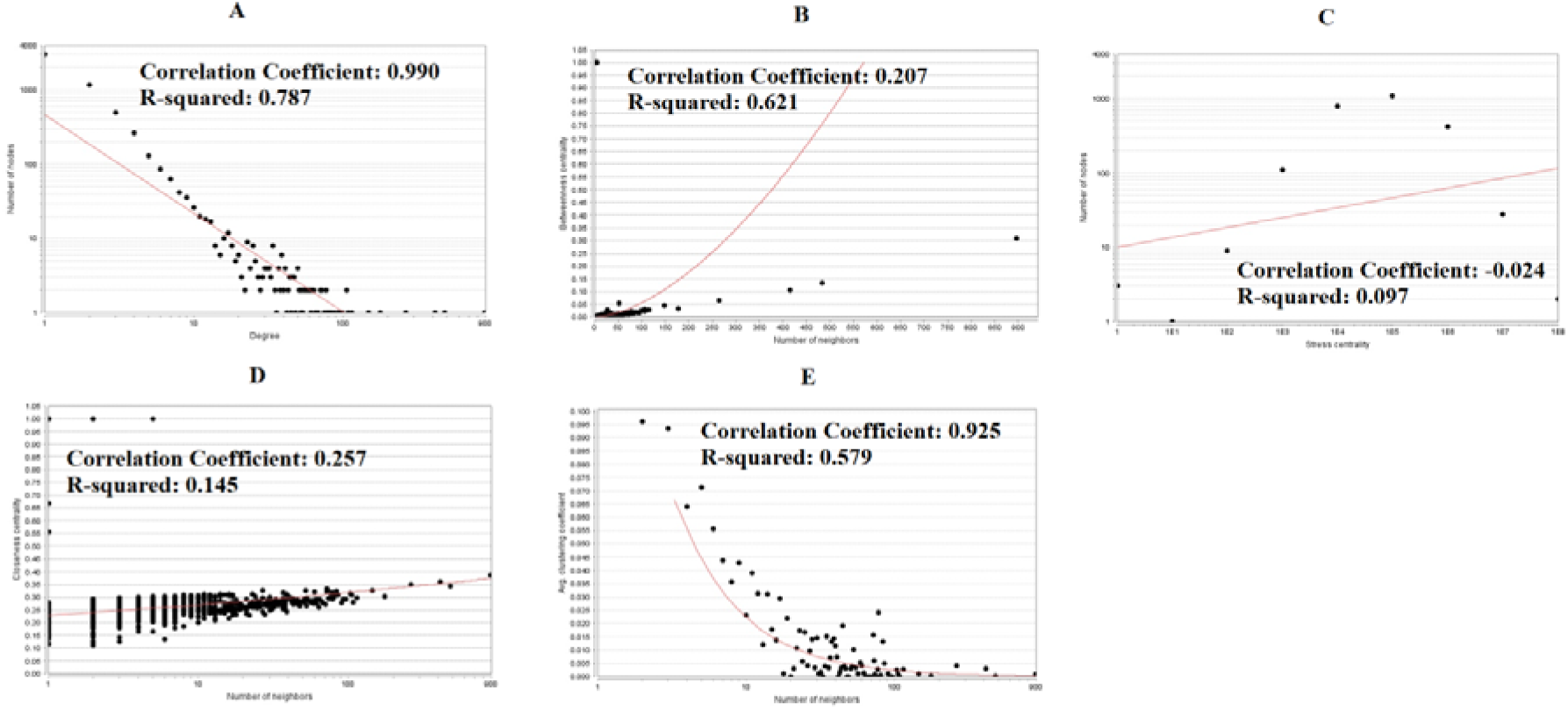
Scatter plot for down regulated genes. (A- Node degree; B- Betweenness centrality; C- Stress centrality; D-Closeness centrality; E- Clustering coefficient)

The PEWCC1 plug-in was also used to identify significant modules. As a result, 4 significant modules (up and down regulated genes) were identified. Modules (Module 6, Module 9, Module 36 and Module 43) for up regulated genes consisted of 137 nodes and 302 edges, 128 nodes and 130 edges, 55 nodes and 103 edges, and 46 nodes and 145 edges are shown in Fig. 9. These modules included up regulated hub genes such as RRM2, PBK, CHEK1, CDC25C, SKP2, TP53, KIF11, CCNA2, CCNB2, CDK1, TOP2A, CDC6, SORT1, FOXM1, CCNB1, CDC20, RACGAP1, RCN2, CAD, HJURP, HDAC11, IGF2BP3, BUB1, NDC80, SKA1, SPC25, NUF2, SPC24, PLK1, MCM7, MCM5, ASF1B, MCM4, MCM6, MCM3, FOXQ1, MCM2 and MMS22, which exhibited the highest score. These up regulated hub genes were enriched in superpathway of pyrimidine deoxyribonucleotides de novo biosynthesis, nuclear division, cell cycle, nuclear division, pathways in cancer, chromosome organization, cell division, DNA double-strand break repair, HTLV-I infection, M phase, metabolism of proteins, E2F transcription factor network, p75(NTR)- mediated signalling, FOXM1 transcription factor network, gene expression, RHO GTPase effectors, aurora B signalling, metabolic pathways, DNA conformation change, alcoholism, PLK1 signaling events, regulation of cell cycle, chromosomal region, protein-containing complex binding, spindle, CDK regulation of DNA replication, DNA metabolic process, embryo development, C-MYB transcription factor network, cellular protein-containing complex assembly, microtubule cytoskeleton, nuclear chromatin, and DNA binding, bending. Modules (Module 6, Module 7, Module 9 and Module 10) for up regulated genes consisted of 147 nodes and 158 edges, 116 nodes and 134 edges, 107 nodes and 113 edges, and 105 nodes and 118 edges are shown in Fig. 10. These modules included down regulated hub genes such as C7, FOS, C8B, PON1, CLU, FGB, CTH, FGG, LPA, USP2, RNF125, HOOK1, PLA2G2A, MAP3K5, GADD45B, SOCS3, MACF1 and GLRX, which exhibited the highest score. These down regulated hub genes were enriched in complement and coagulation cascades, ATF-2 transcription factor network, defense response, metabolic pathways, lipid metabolic process, response to metal ion, organic acid metabolic process, IL6-mediated signaling events, amb2 integrin signalling, ion transport, endoplasmic reticulum membrane, identical protein binding, glypican 1 network, p38 MAPK signaling pathway, response to other organism, golgi apparatus oxidation-reduction process.

**Fig. 9.**
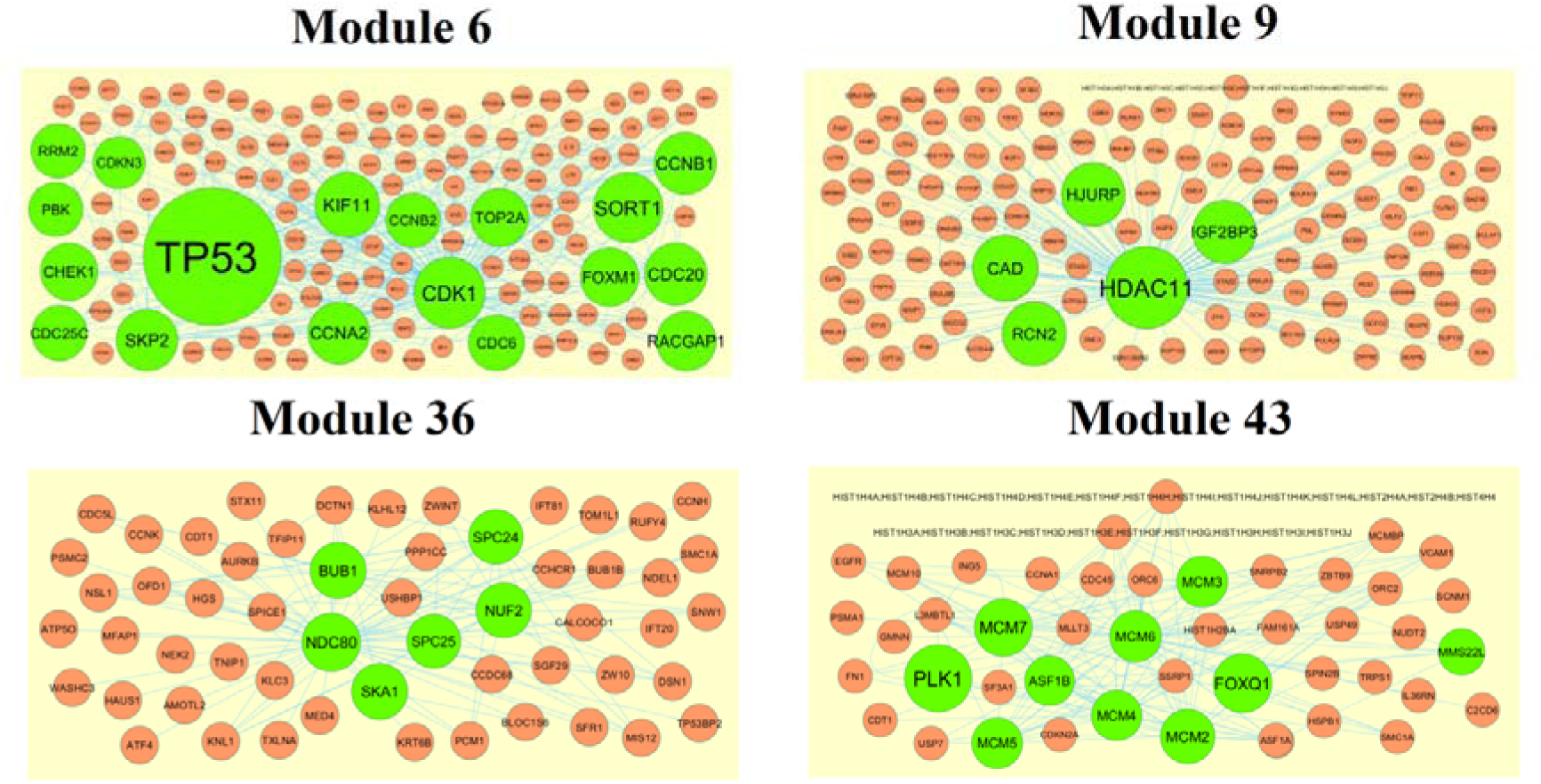
Modules in PPI network. The green nodes denote the up regulated genes

**Fig. 10.**
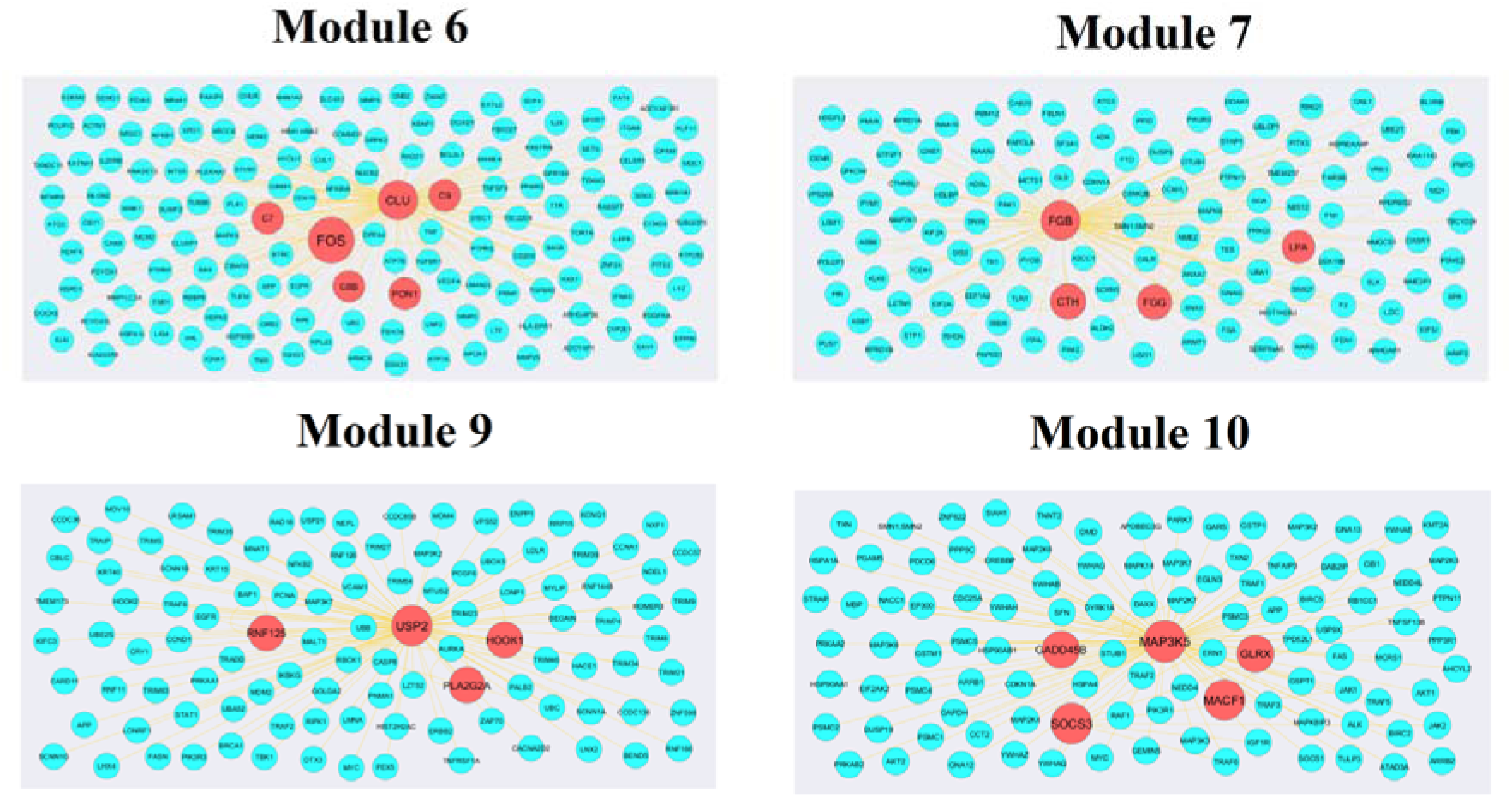
Modules in PPI network. The red nodes denote the down regulated genes

### Construction of target genes - miRNA regulatory network

We constructed a target genes - miRNA regulatory network, and the network is shown in Fig. 11 and Fig. 12, which illustrates that certain miRNAs play important roles in regulating up and down regulated genes. In Fig. 11, nodes with greater degrees tend to be up regulated target genes. In this network, PLAGL2 was targeted by the 189 miRNAs (ex, hsa-mir-4710), CDK6 was targeted by the 188 miRNAs (ex, hsa-mir-4657), CCND2 was targeted by the 179 miRNAs (ex, hsa-mir-5192), SLC1A5 was targeted by the 174 miRNAs (ex, hsa-mir-4284) and PKM was targeted by the 160 miRNAs (ex, hsa-mir-4525) are listed in Table 7. These up regulated target genes were enriched in nuclear chromatin, cell cycle, nuclear division, integral component of plasma membrane and glycolysis. In Fig. 12, nodes with greater degrees tend to be down regulated target genes. In this network, SOD2 was targeted by the 257 miRNAs (ex, hsa-mir-4803), WEE1 was targeted by the 182 miRNAs (ex, hsa-mir-4647), SULT1B1 was targeted by the 131 miRNAs (ex, hsa-mir-3163), APOL6 was targeted by the 127 miRNAs (ex, hsa-mir-4455) and DHODH was targeted by the 119 miRNAs (ex, hsa-mir-1200) are listed in Table 7. These down regulated target genes were enriched in peroxisome, biological oxidations, lipid binding and metabolic pathways.

**Fig. 11.**
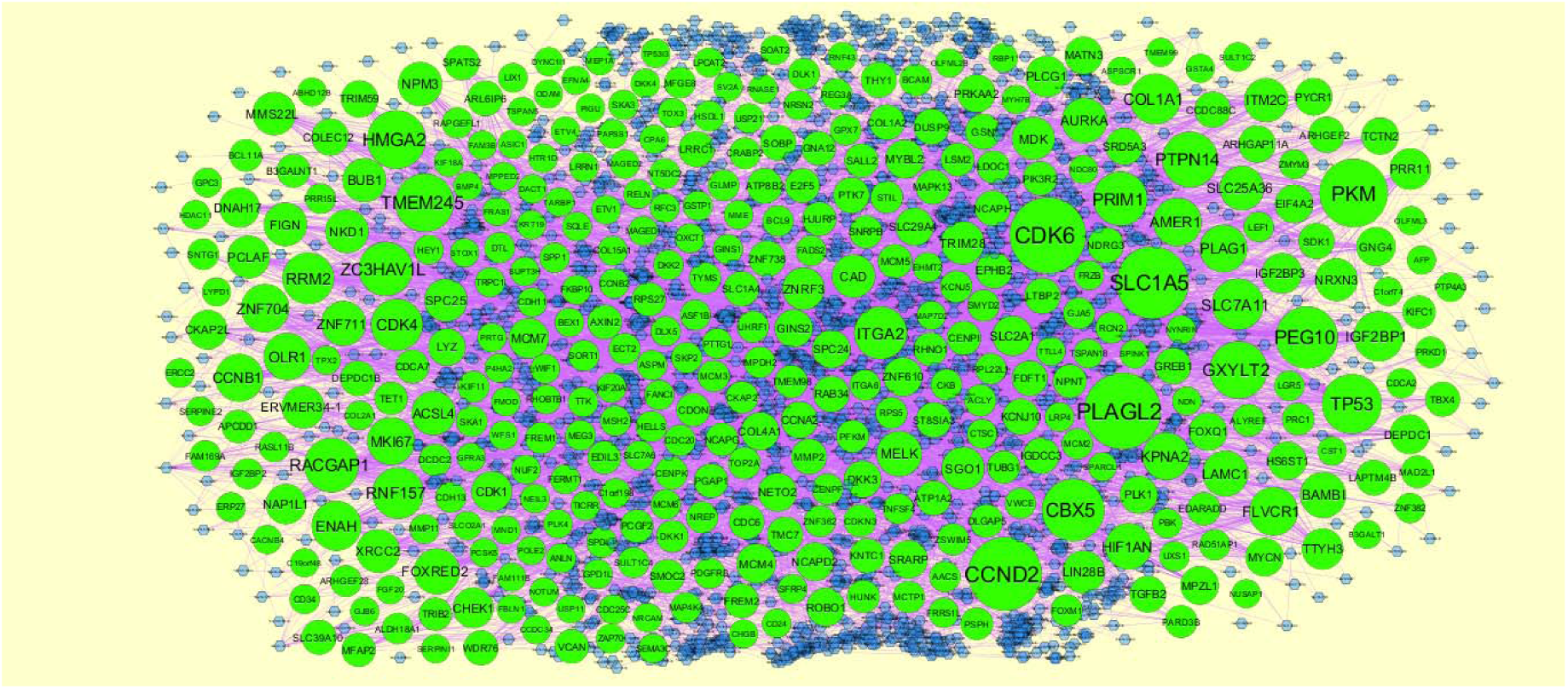
The network of up regulated genes and their related miRNAs. The green circles nodes are the up regulated genes, and blue diamond nodes are the miRNAs

**Fig. 12.**
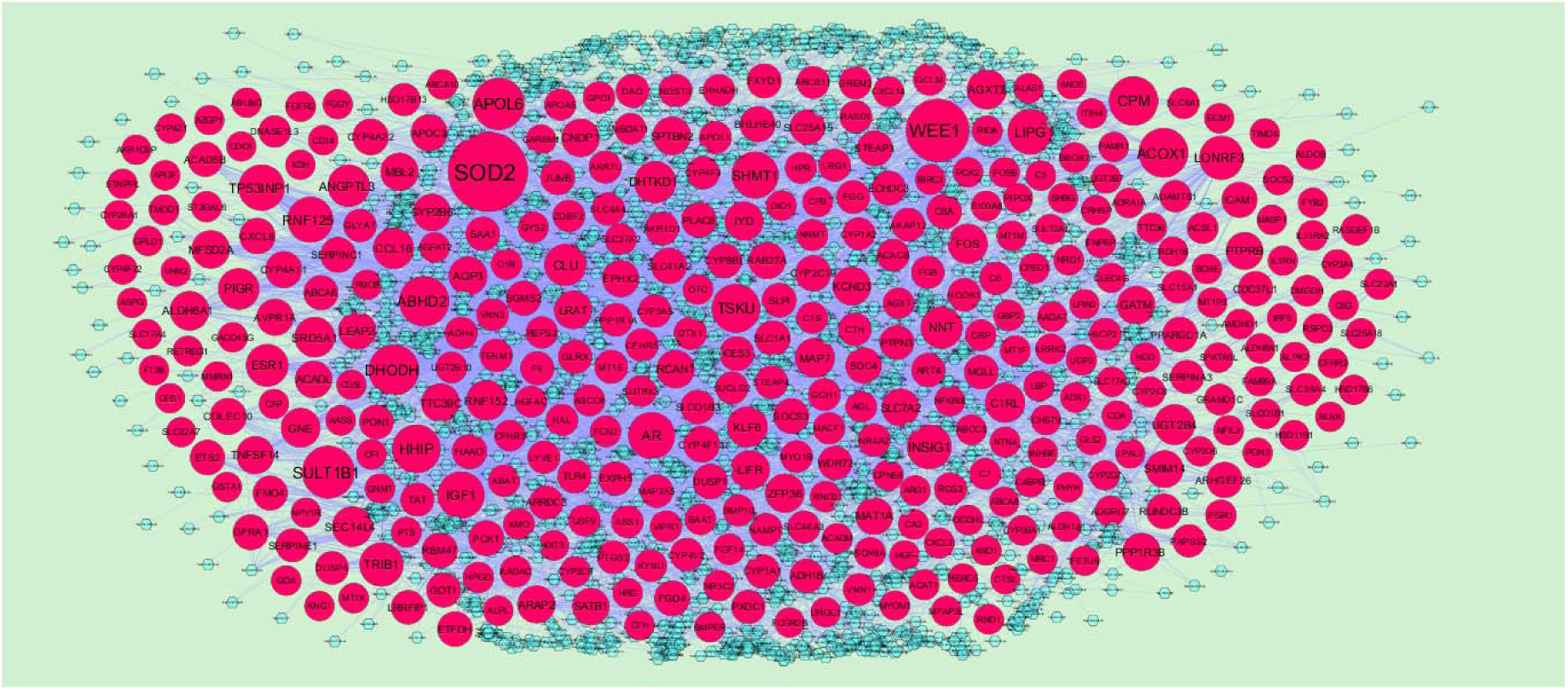
The network of down regulated genes and their related miRNAs. The pink circles nodes are the down regulated genes, and sky blue diamond nodes are the miRNAs

**Table 7.**
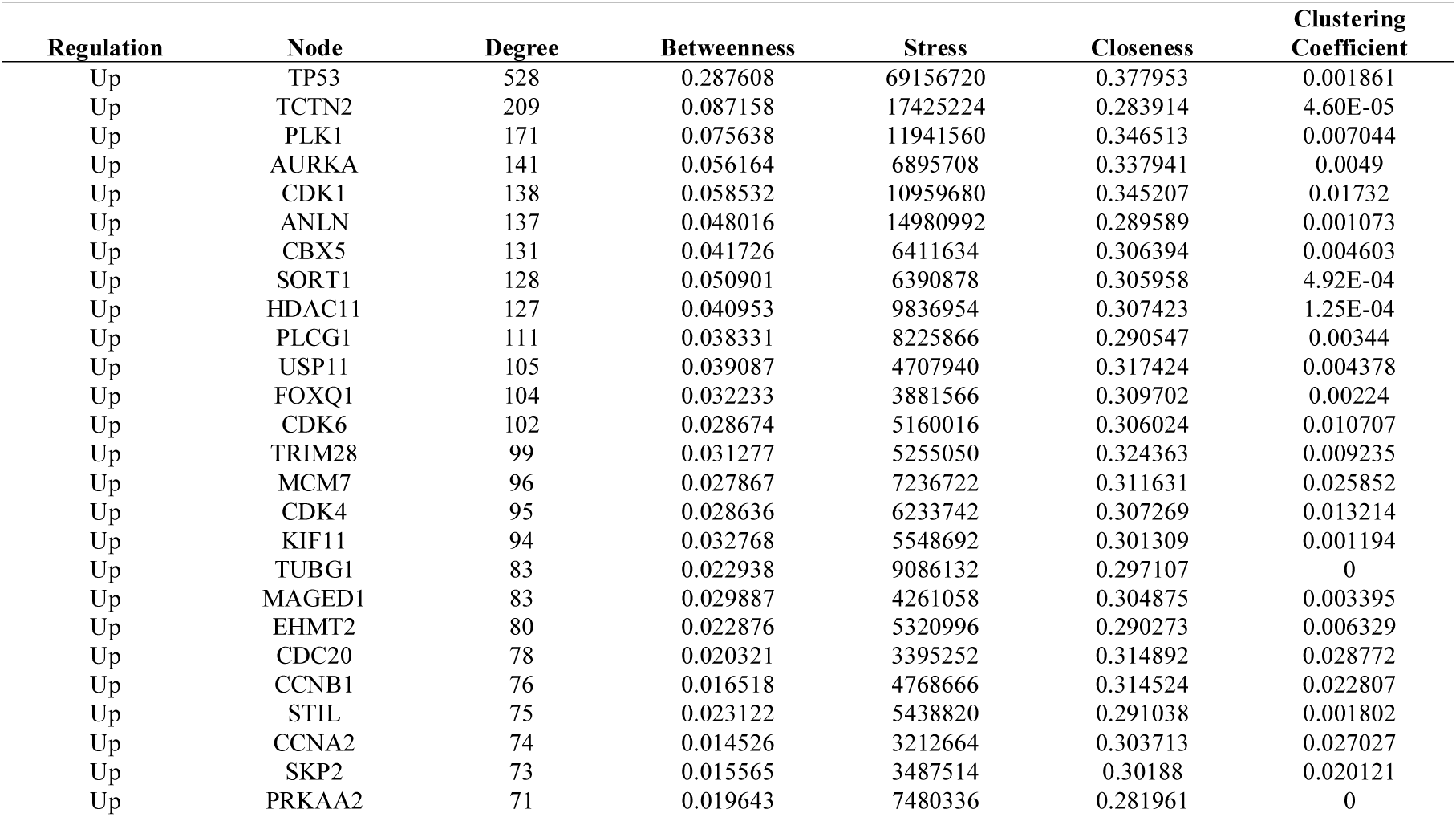
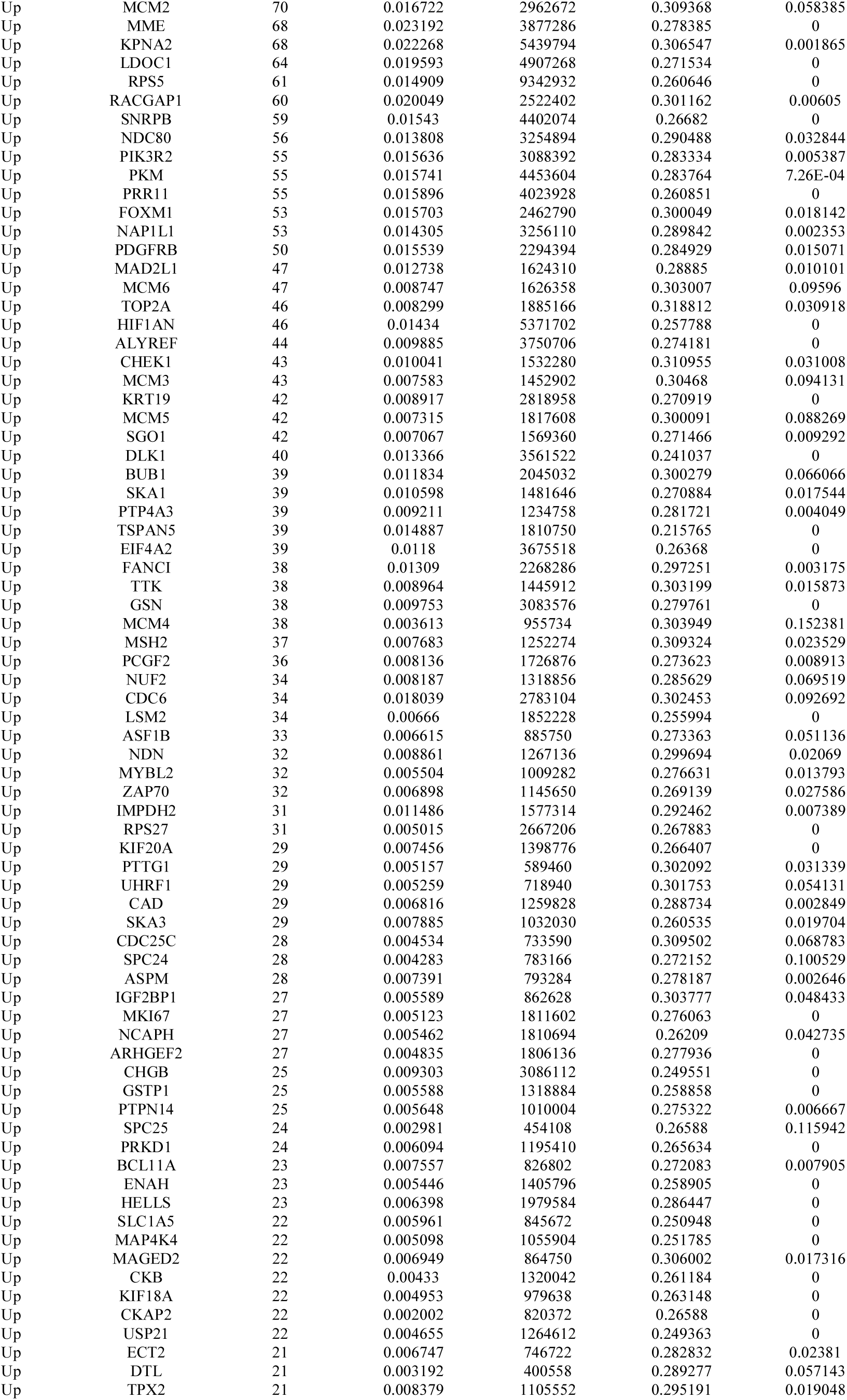
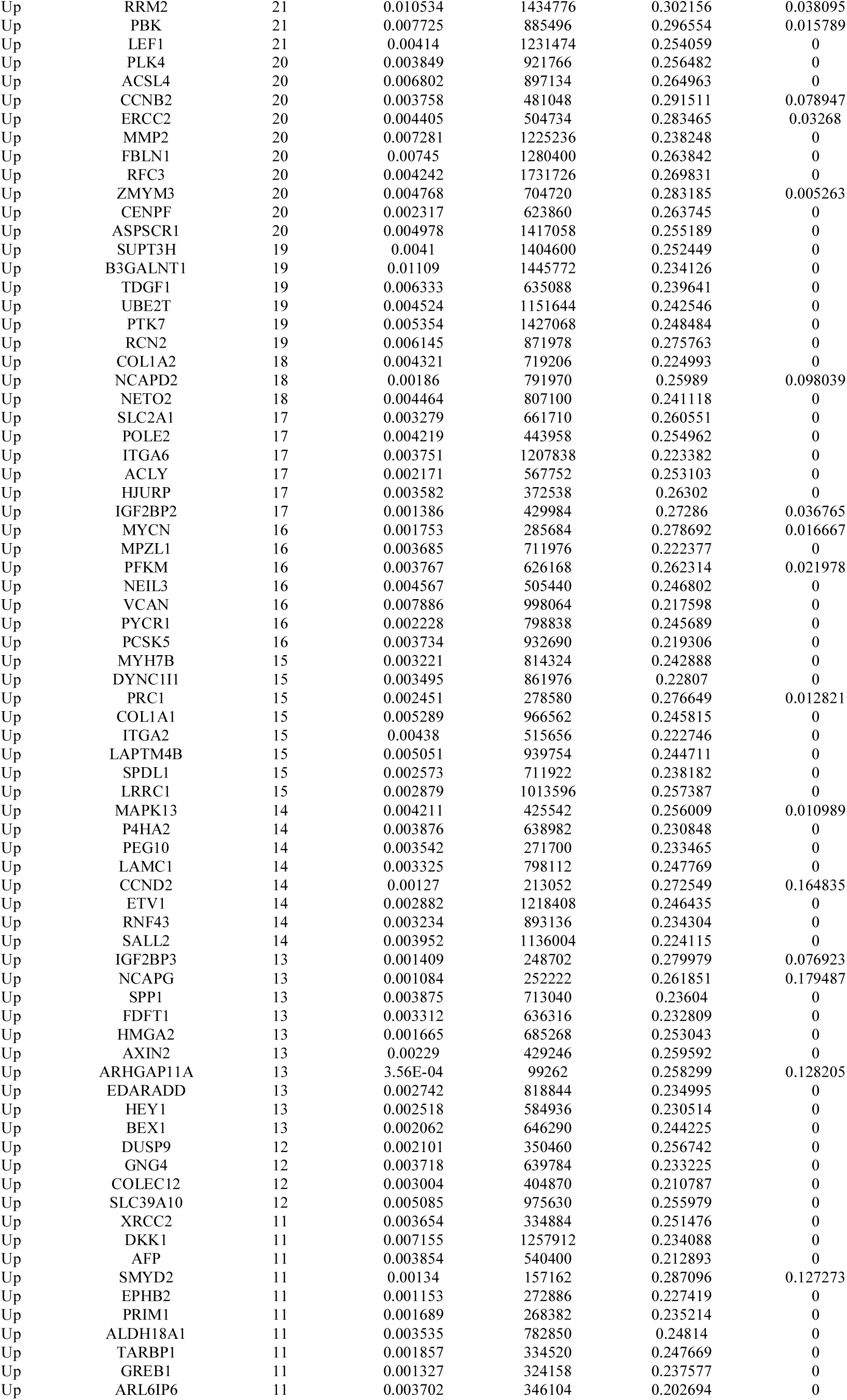
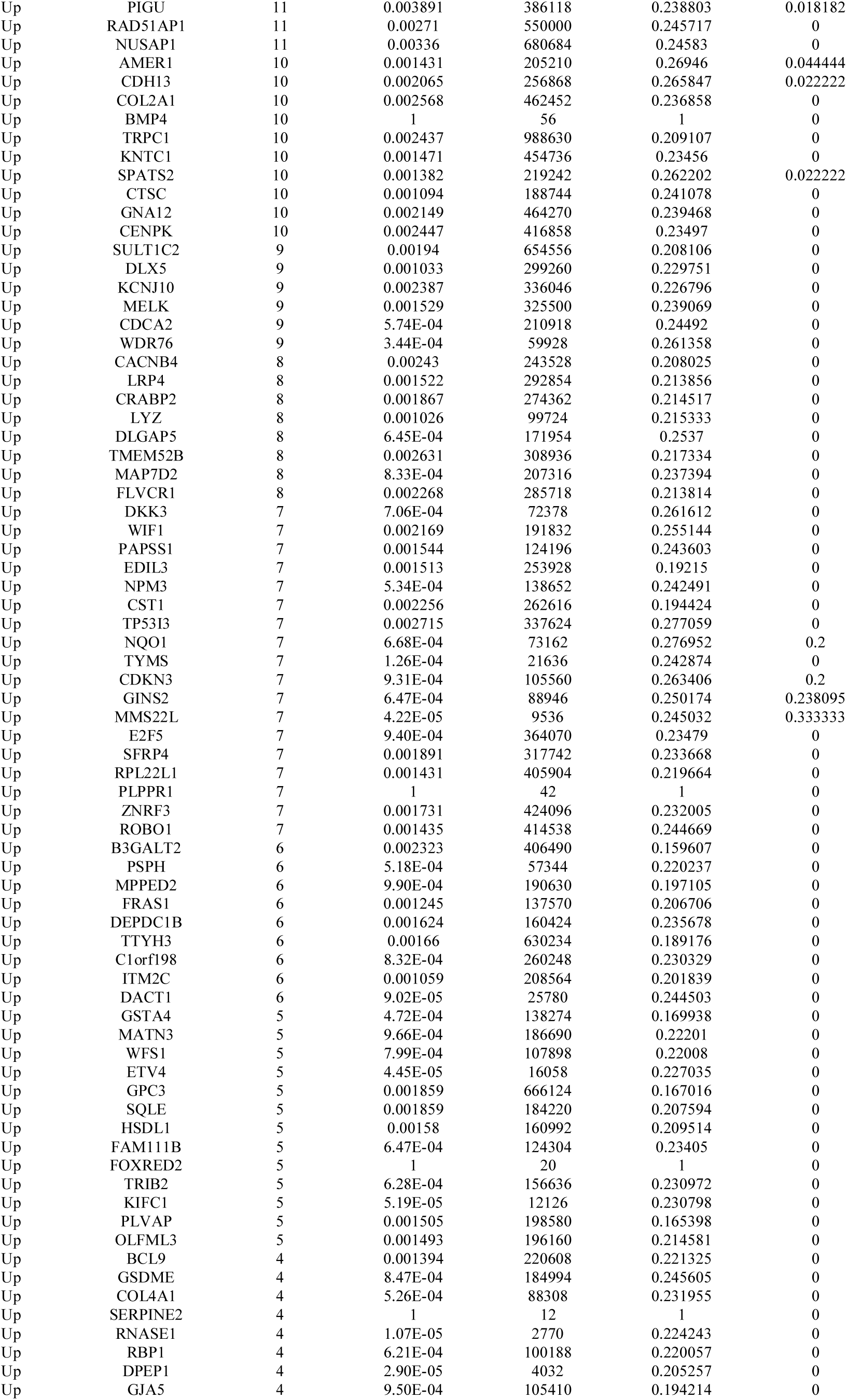
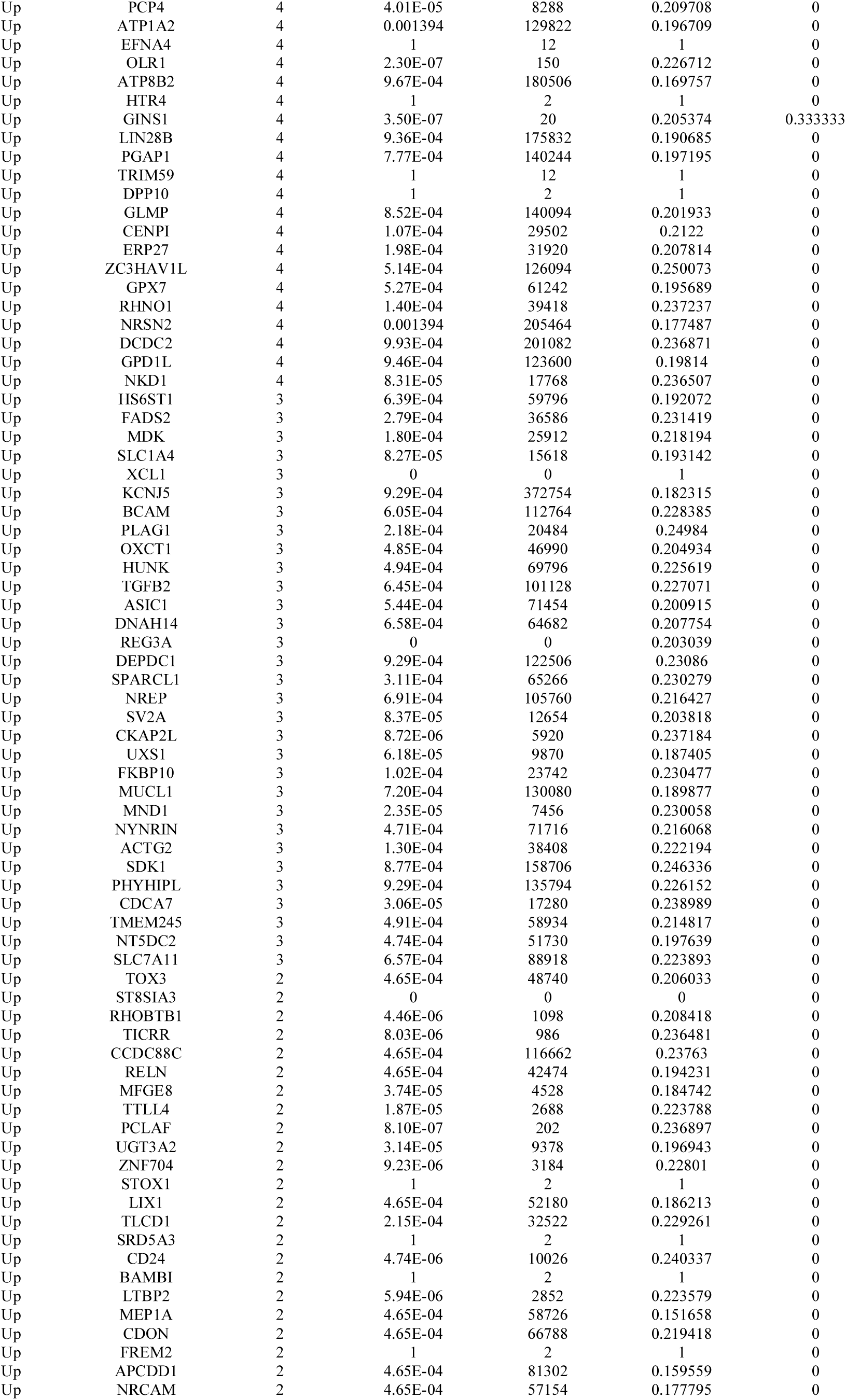
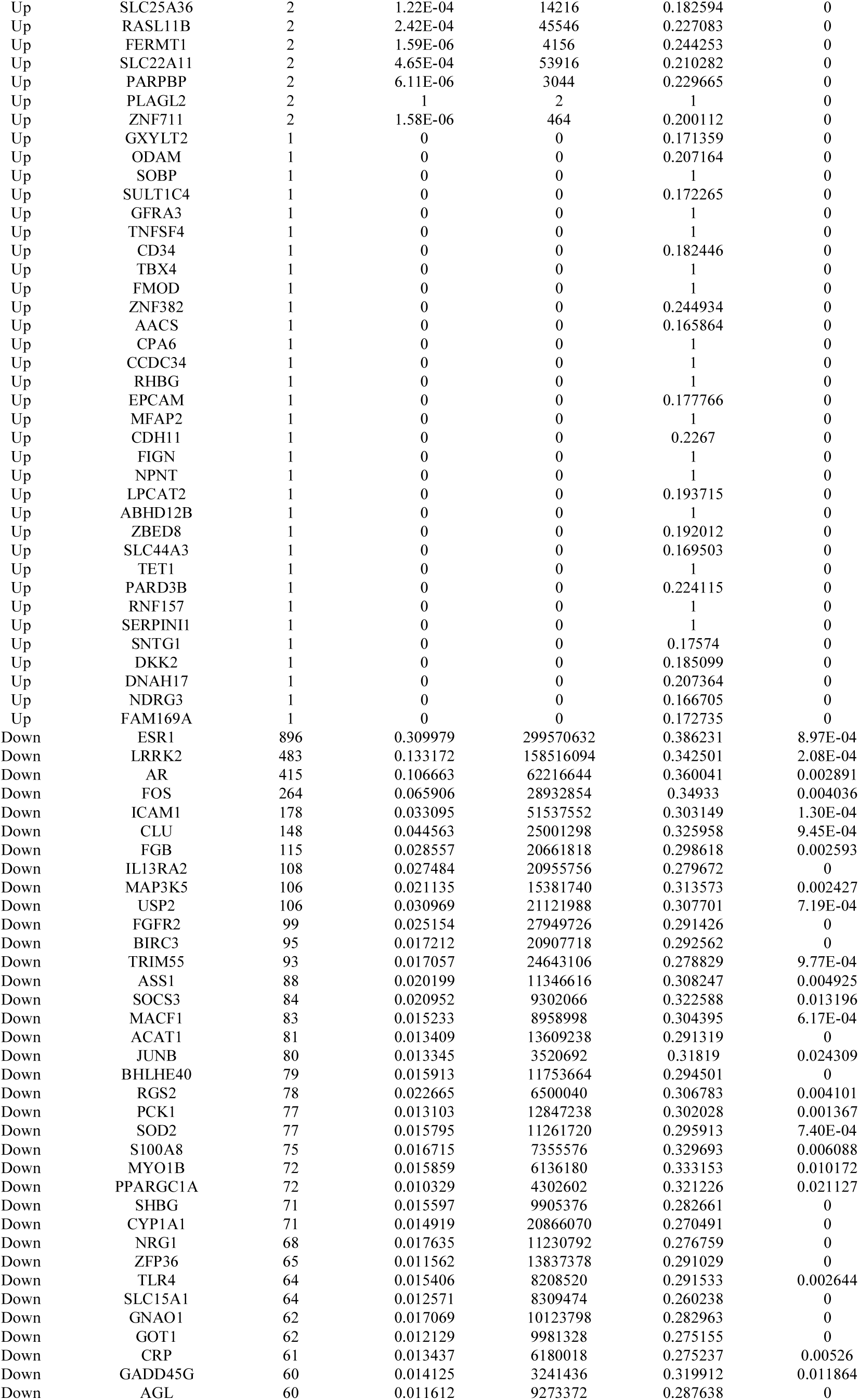
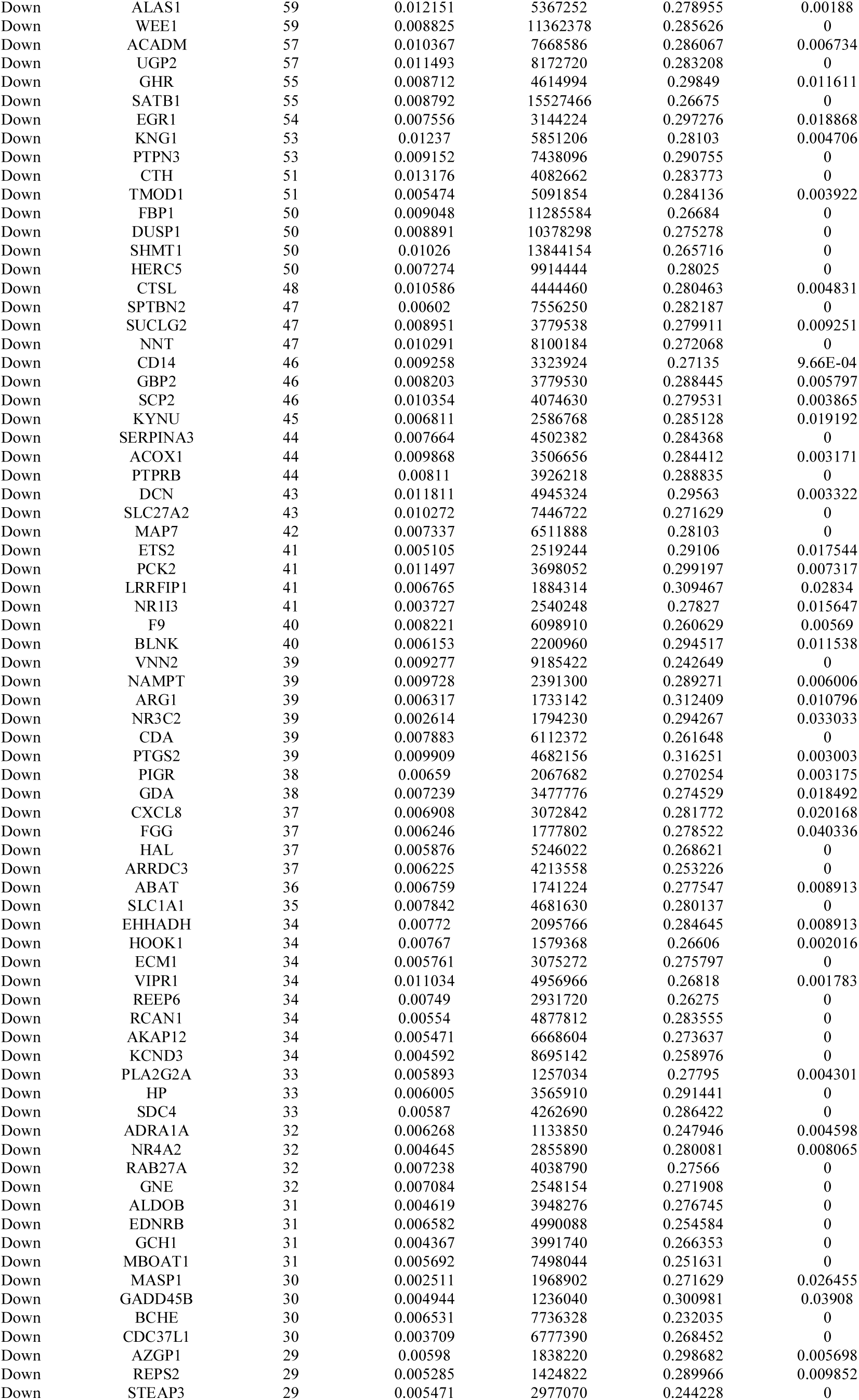
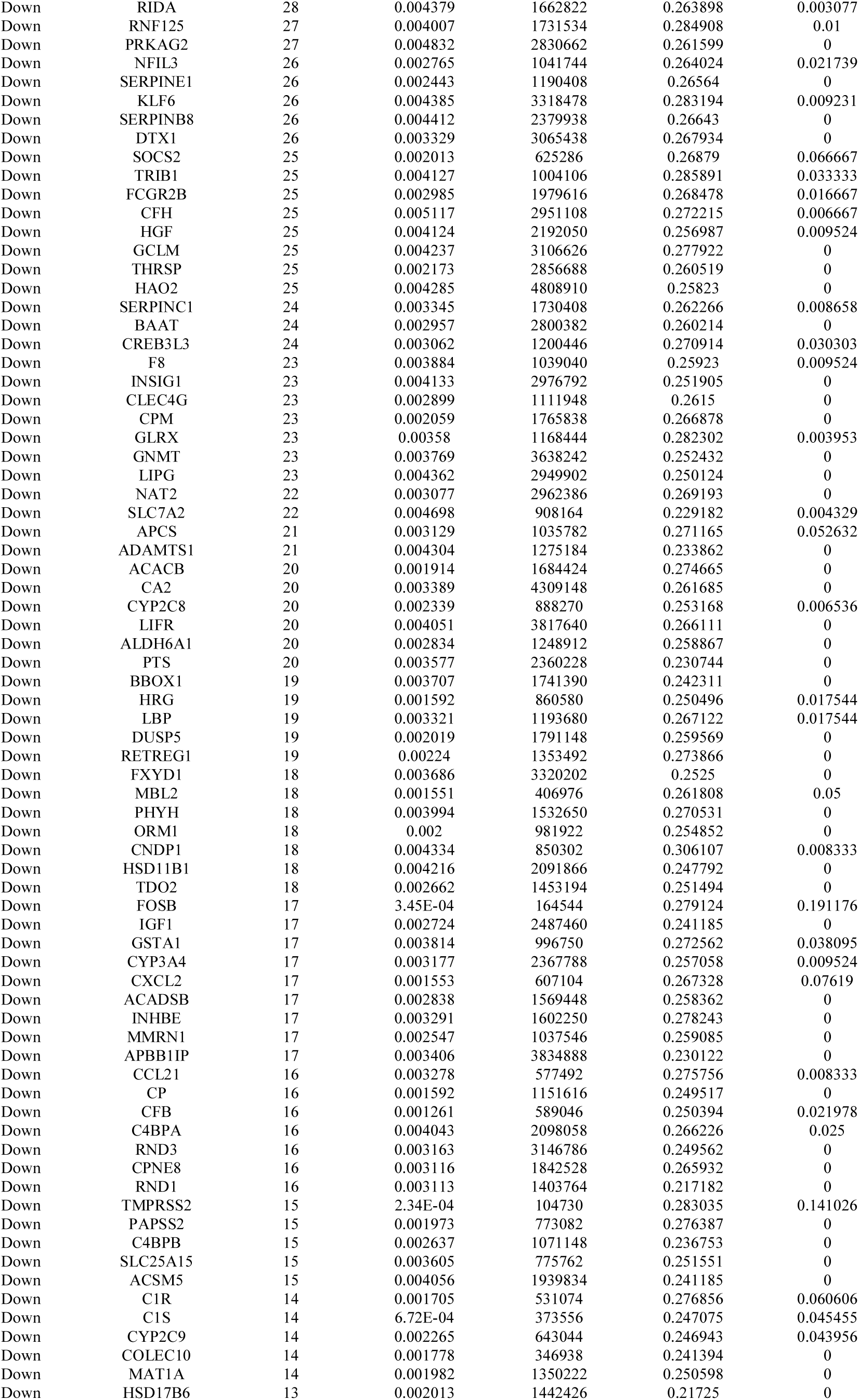
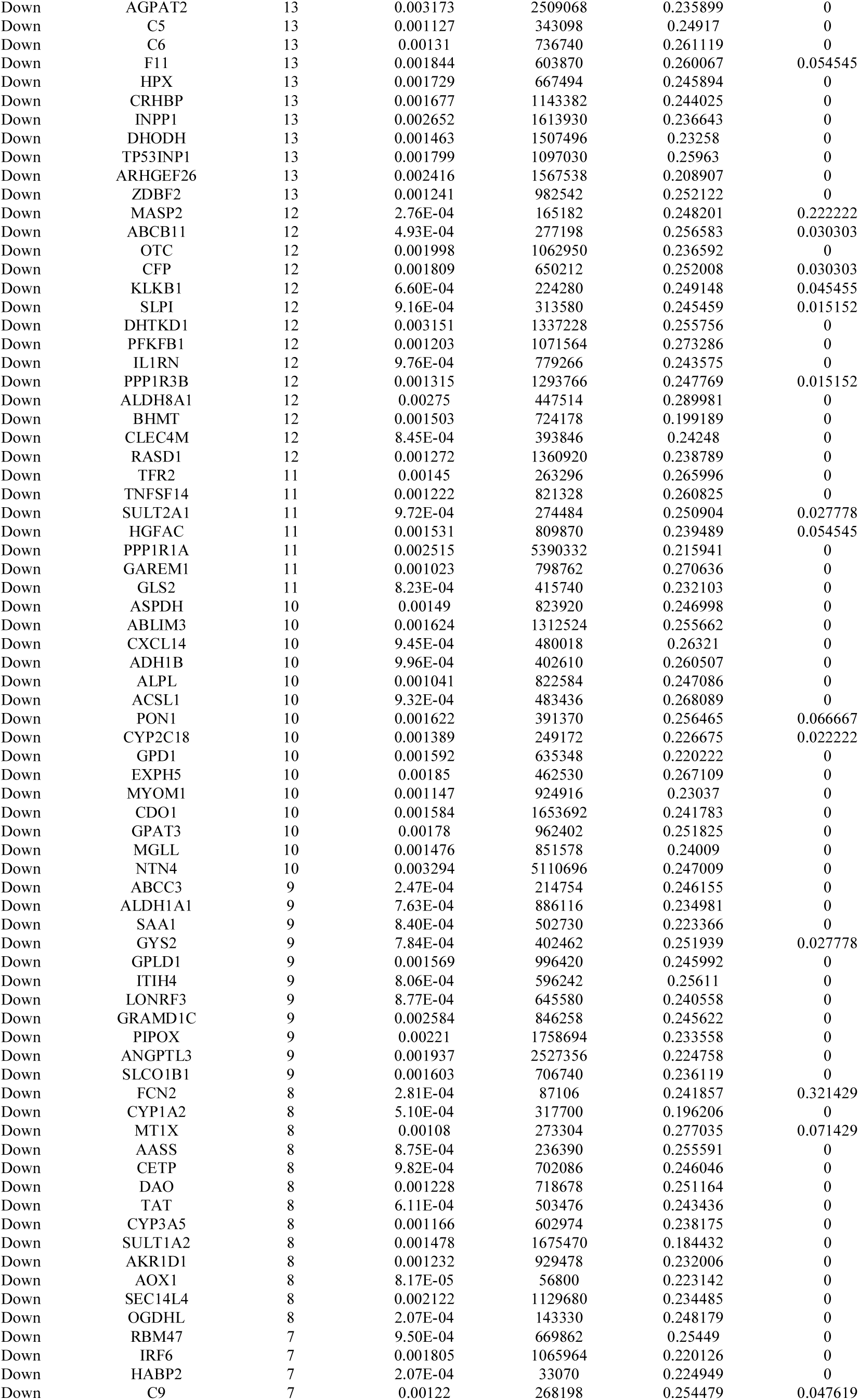
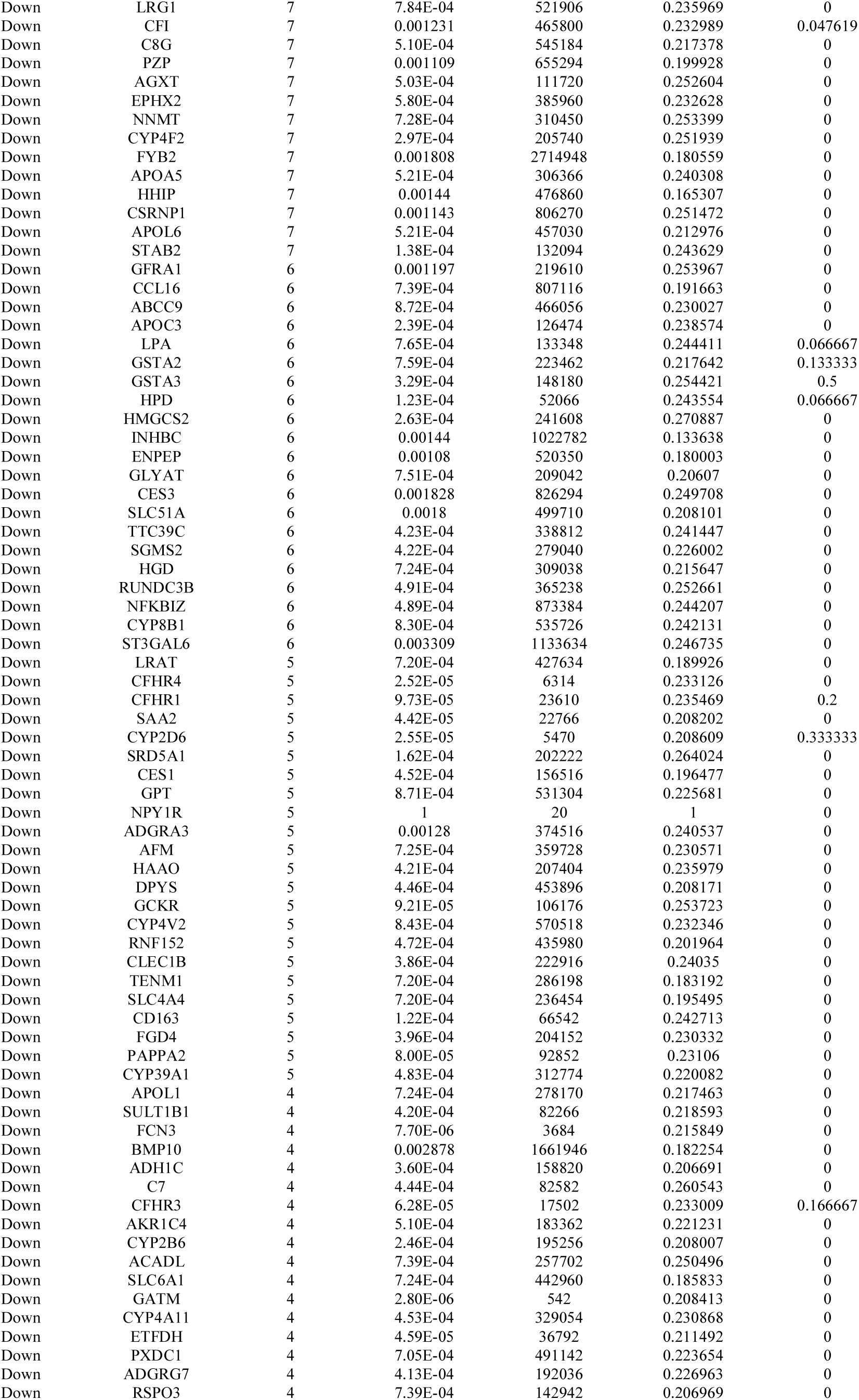
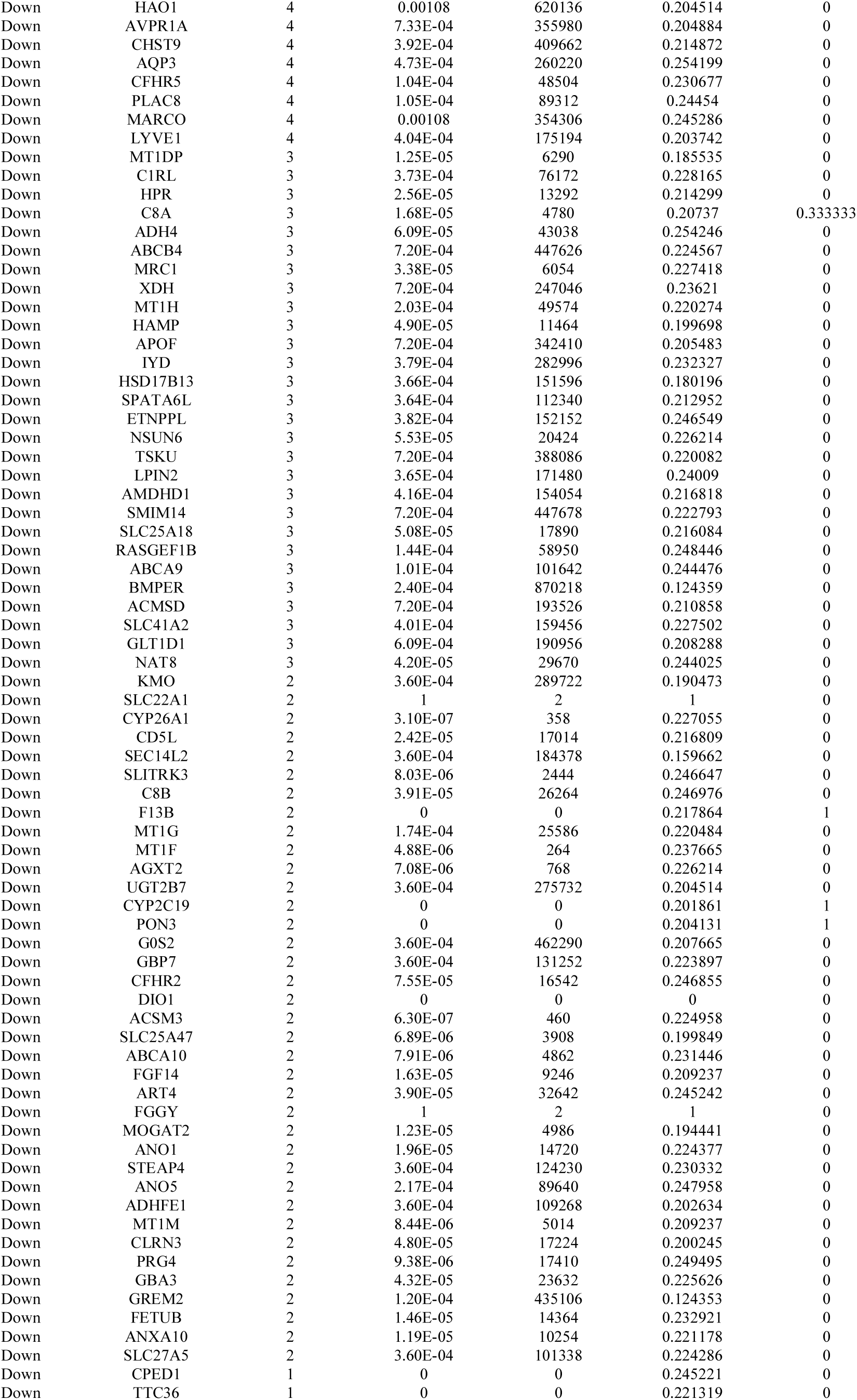
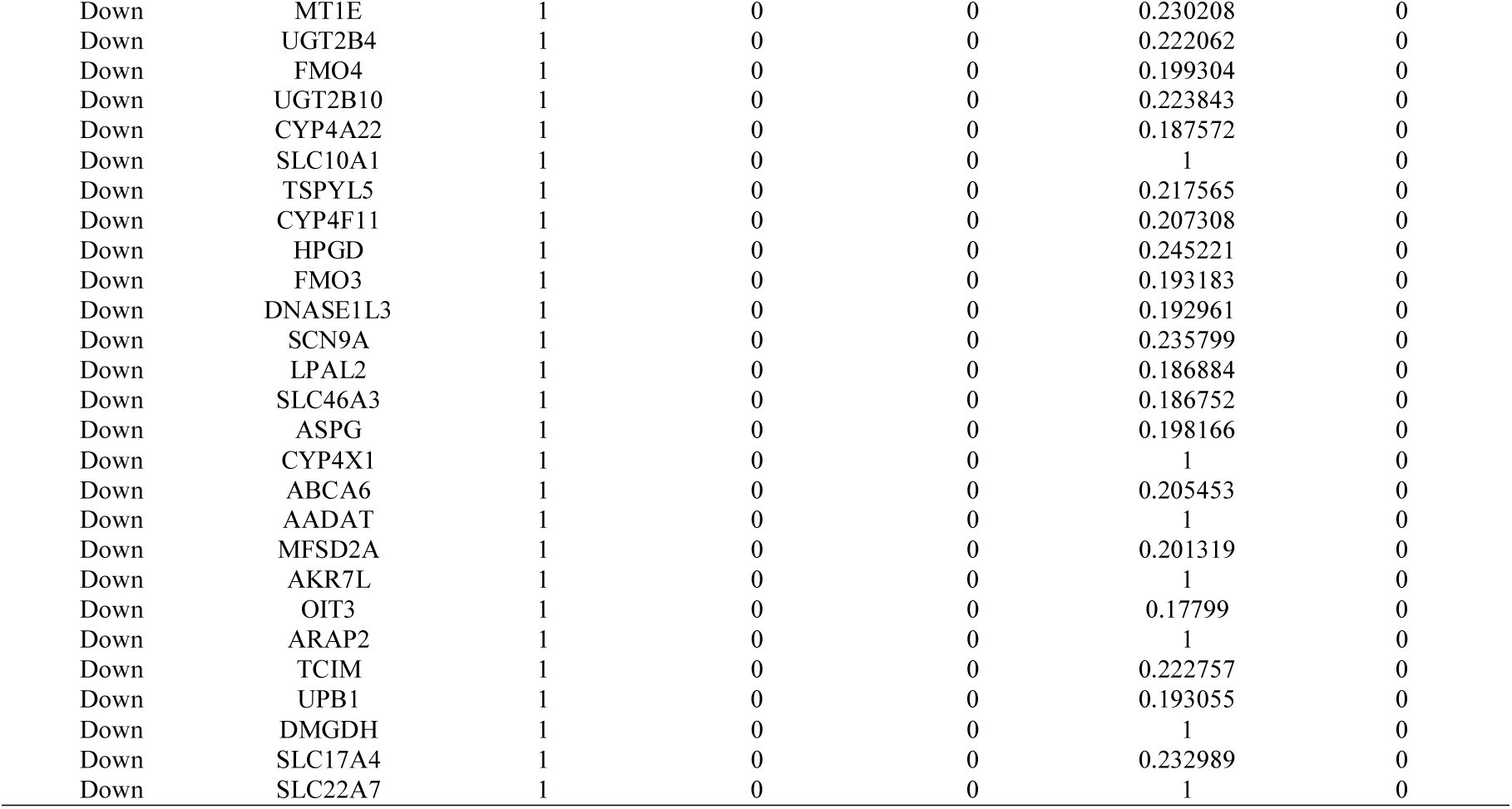
Topology table for up and down regulated genes

### Construction of target genes - TF regulatory network

We constructed a target gene - TF regulatory network, and the network is shown in Fig. 13 and Fig. 14, which illustrates that certain TFs play important roles in regulating up and down regulated genes. In Fig. 13, nodes with greater degrees tend to be up regulated target genes. In this network, DCDC2 was targeted by the 121 TFs (ex, KLF9), WFS1 was targeted by the 120 TFs (ex, TFDP1), ERCC2 was targeted by the 117 TFs (ex, ZNF580), DLX5 was targeted by the 107 TFs (ex, MXD3) and CCNB1 was targeted by the 105 TFs (ex, SMAD5) are listed in Table 8. These up regulated target genes were enriched in generation of neurons, metabolism of proteins, gene expression, embryo development and cell cycle. In Fig. 14, nodes with greater degrees tend to be down regulated target genes. In this network, MASP2 was targeted by the 115 TFs (ex, ATF3), OTC was targeted by the 82 TFs (ex, YY1), ACMSD was targeted by the 76 TFs (ex, MYBL2), AGXT was targeted by the 76 TFs (ex, MBD4) and SLC15A1 was targeted by the 76 TFs (ex, HNF4G) are listed in Table 8. These down regulated target genes were enriched in complement and coagulation cascades, metabolic pathways, superpathway of tryptophan utilization, glycine biosynthesis and ion transport.

**Fig. 13.**
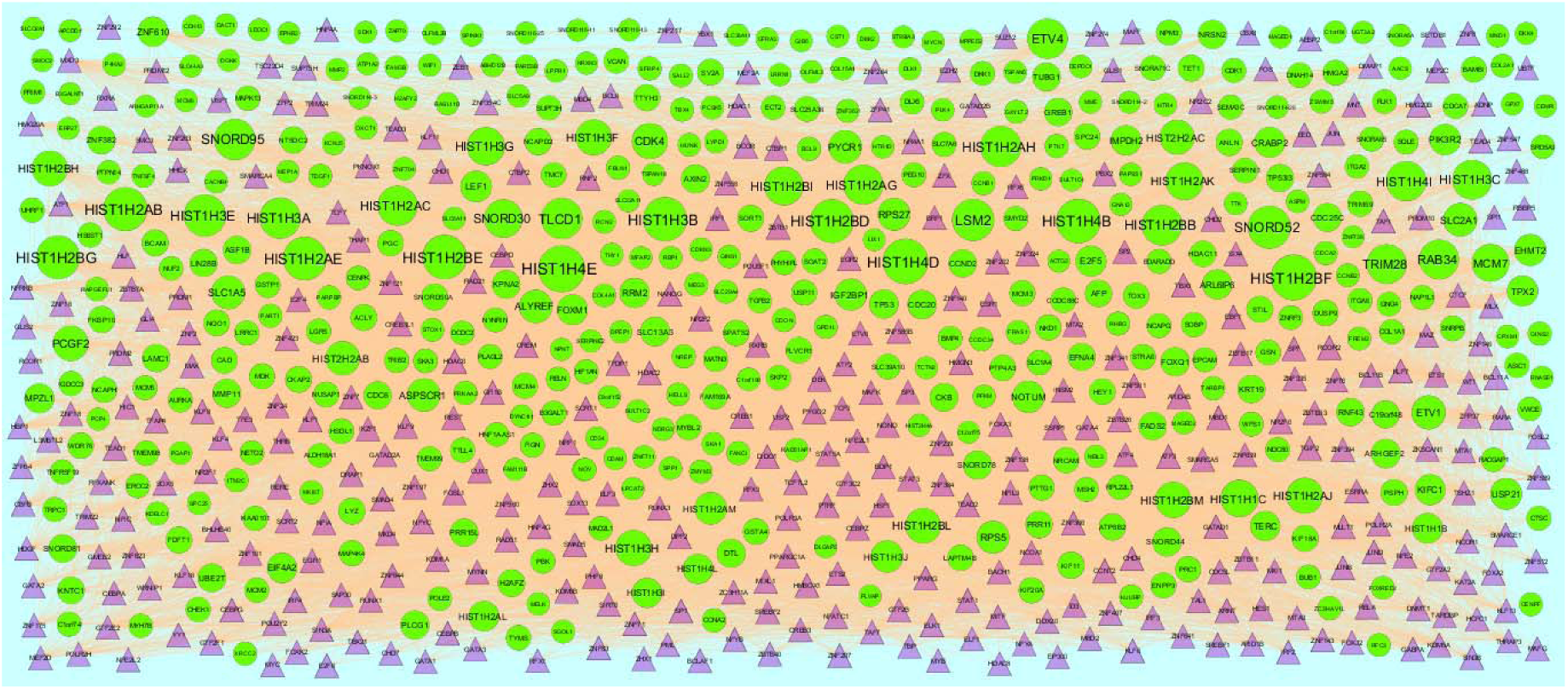
The network of up regulated genes and their related TFs. The green circles nodes are the up regulated genes, and purple triangle nodes are the TFs

**Fig. 14.**
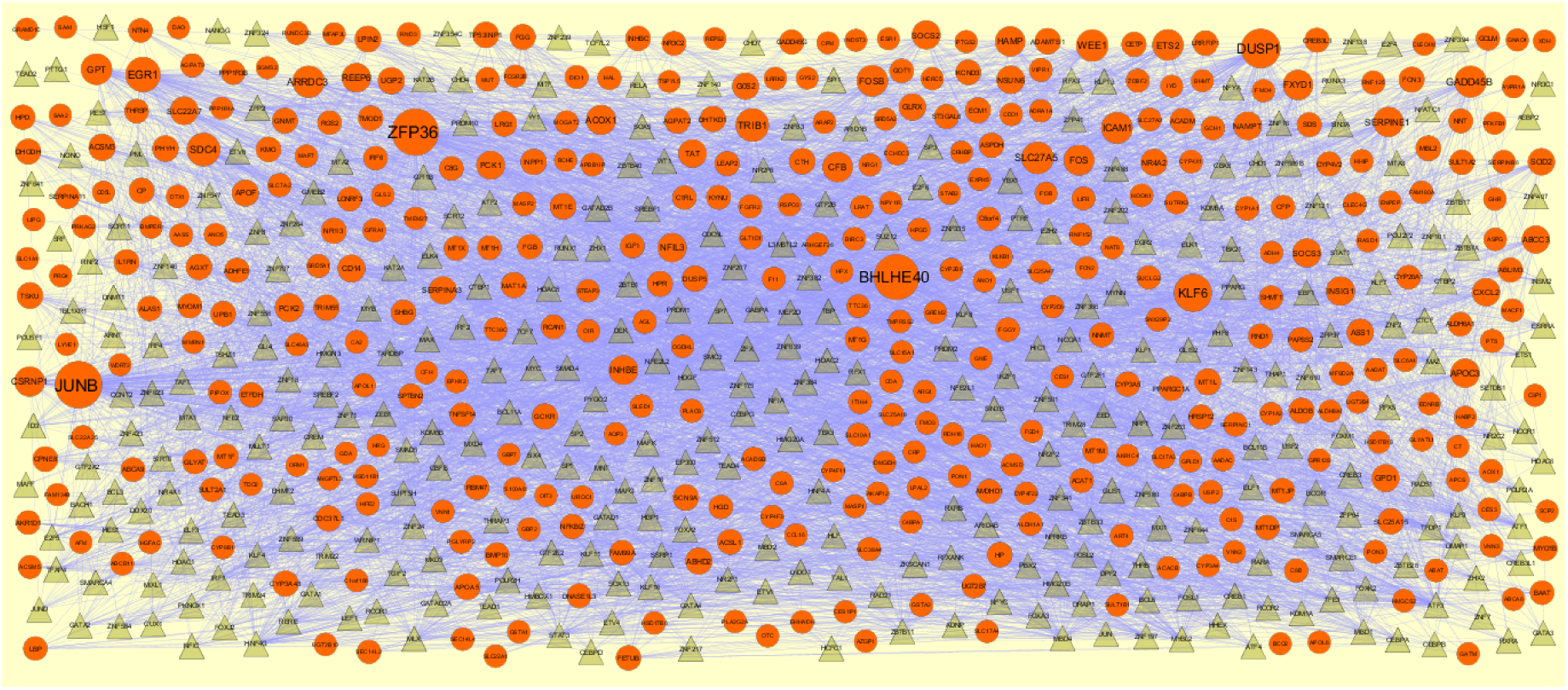
The network of down regulated genes and their related TFs. The green circles nodes are the down regulated genes, and yellow triangle nodes are the TFs.

**Table 8.**
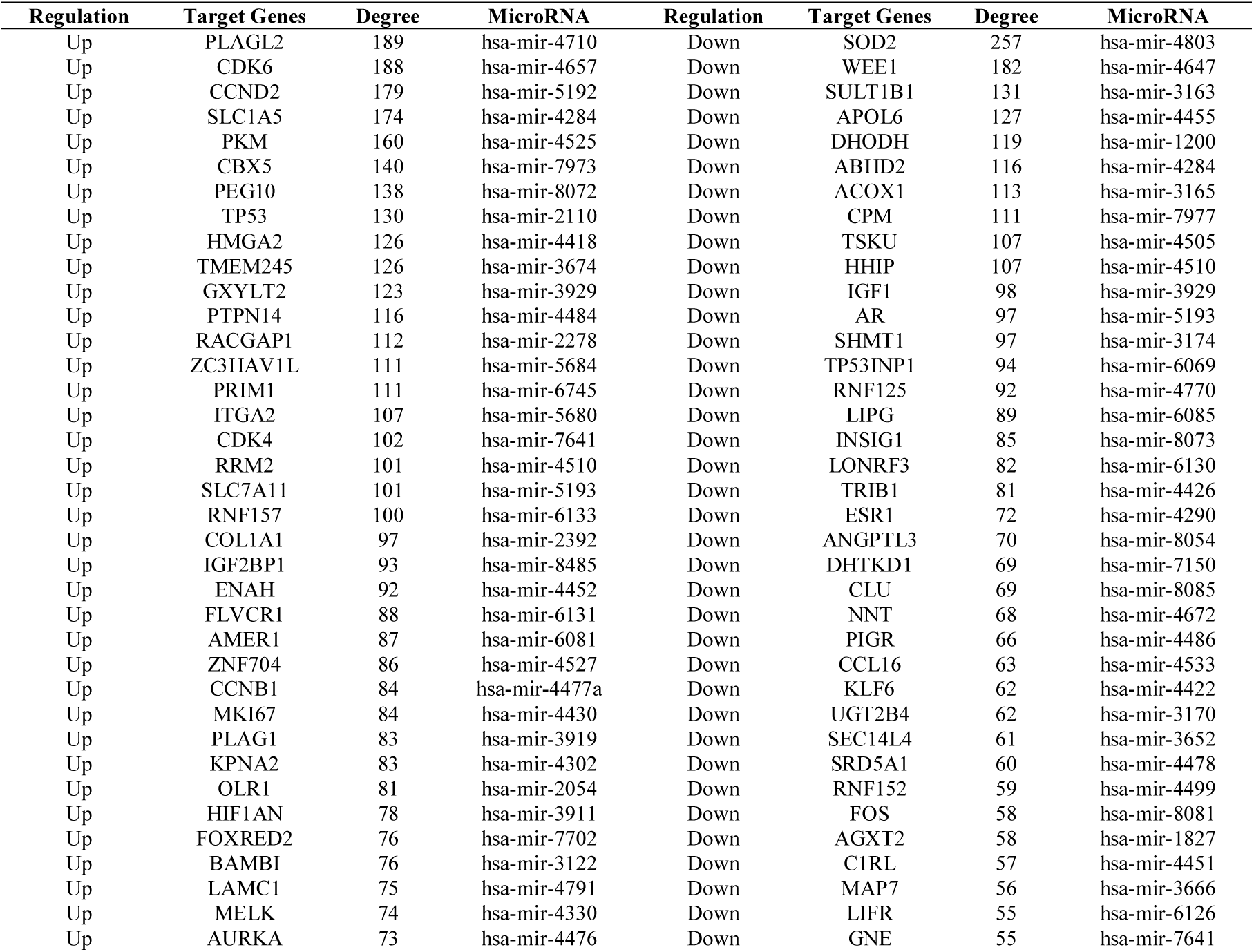
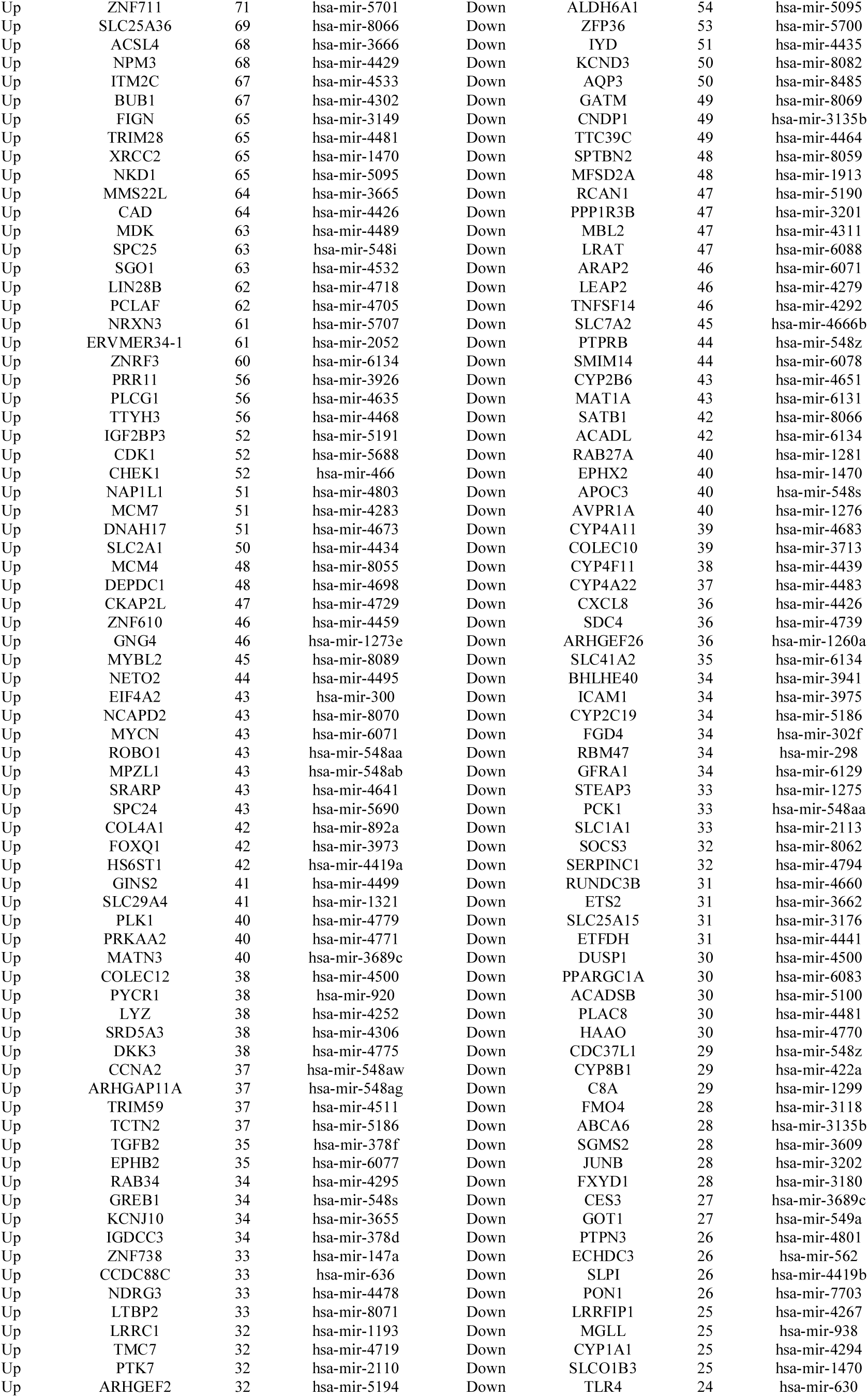
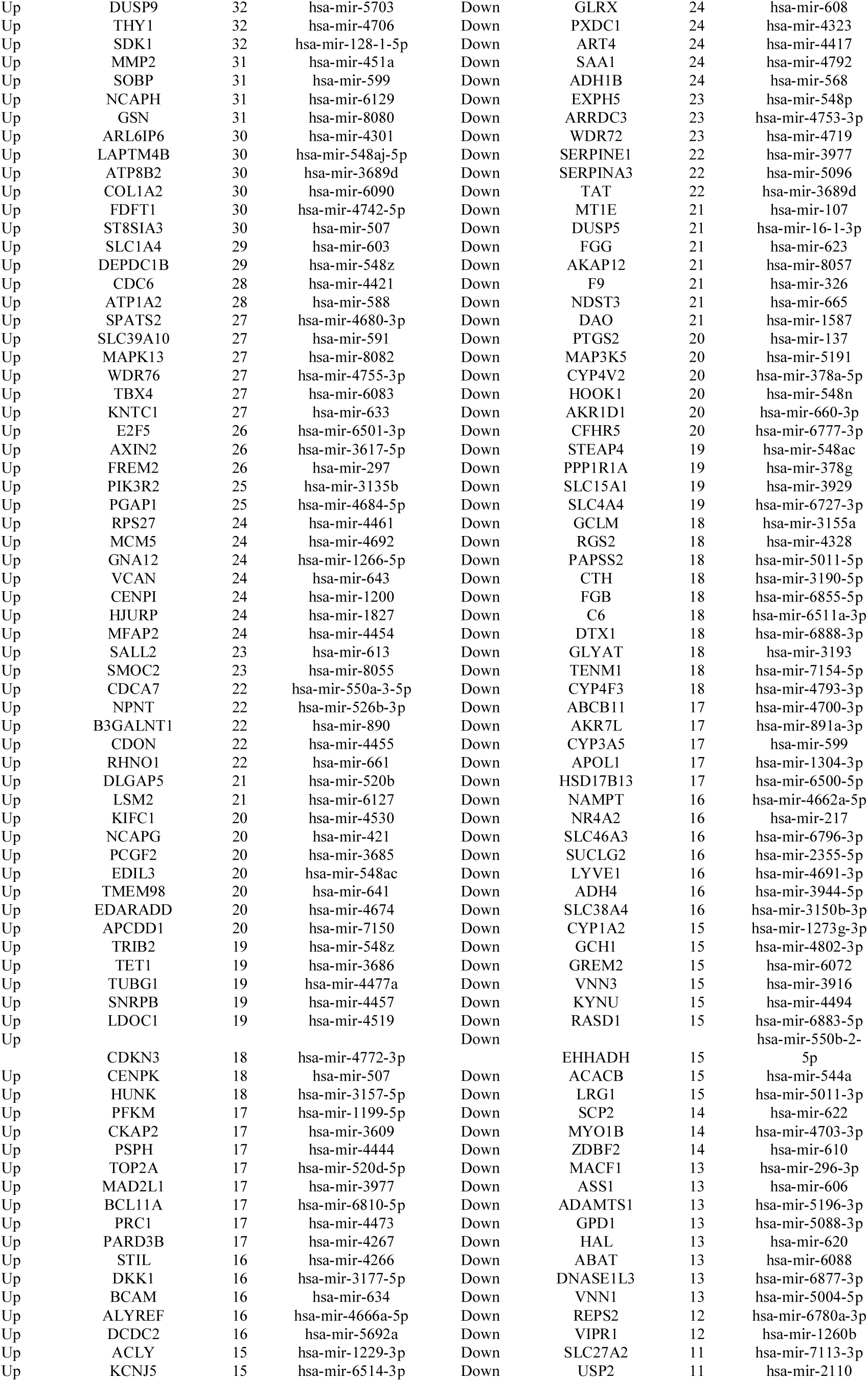
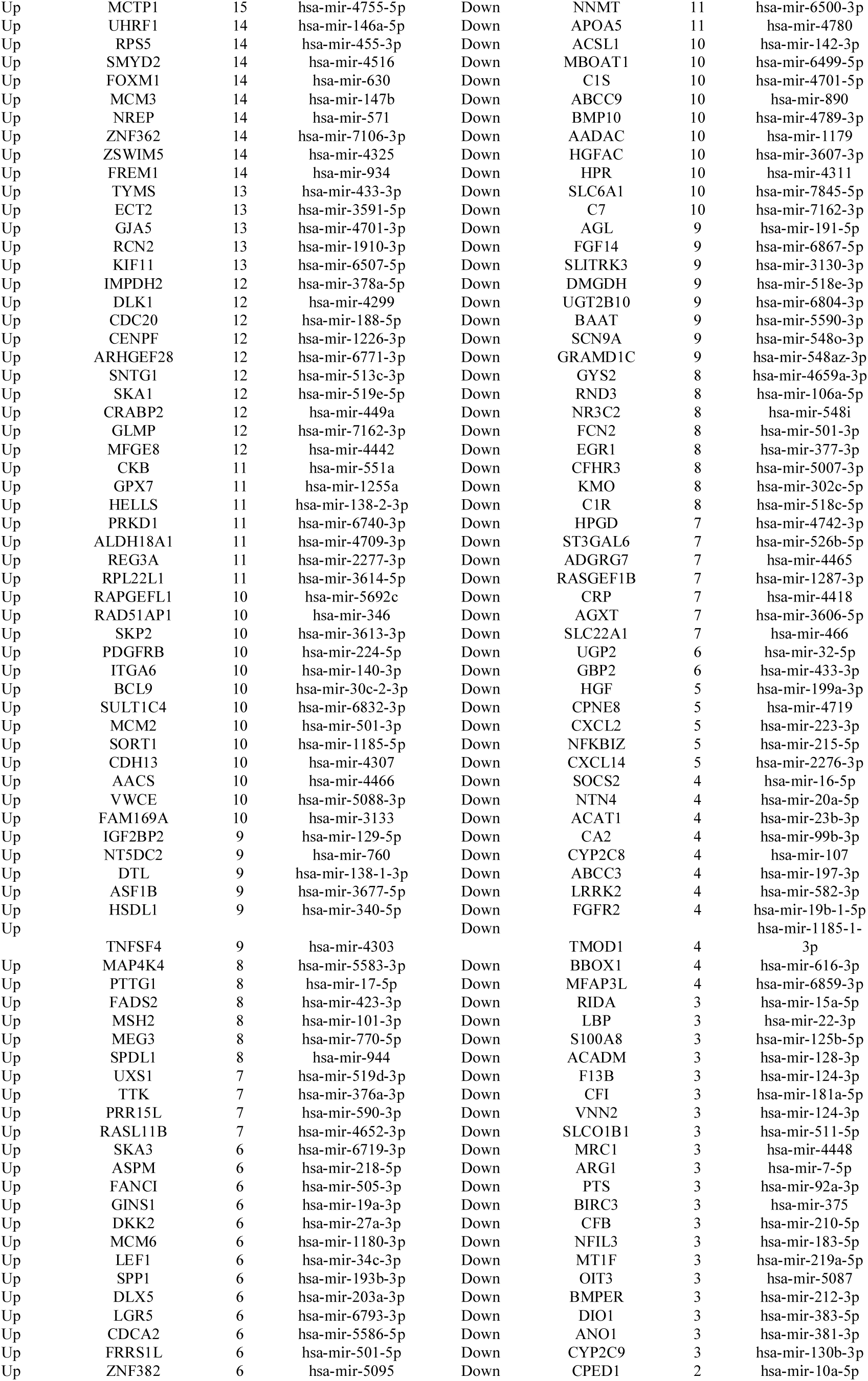
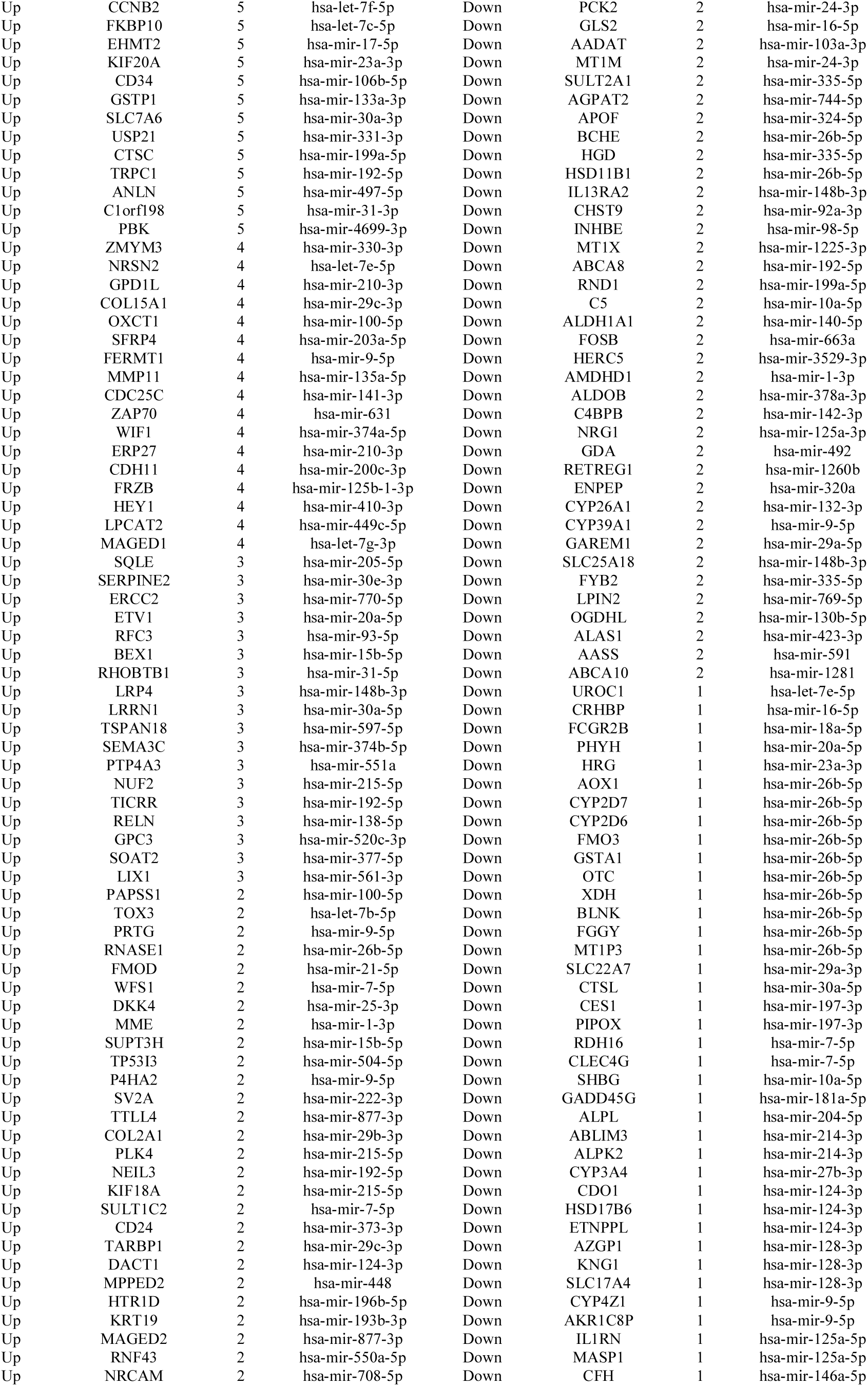
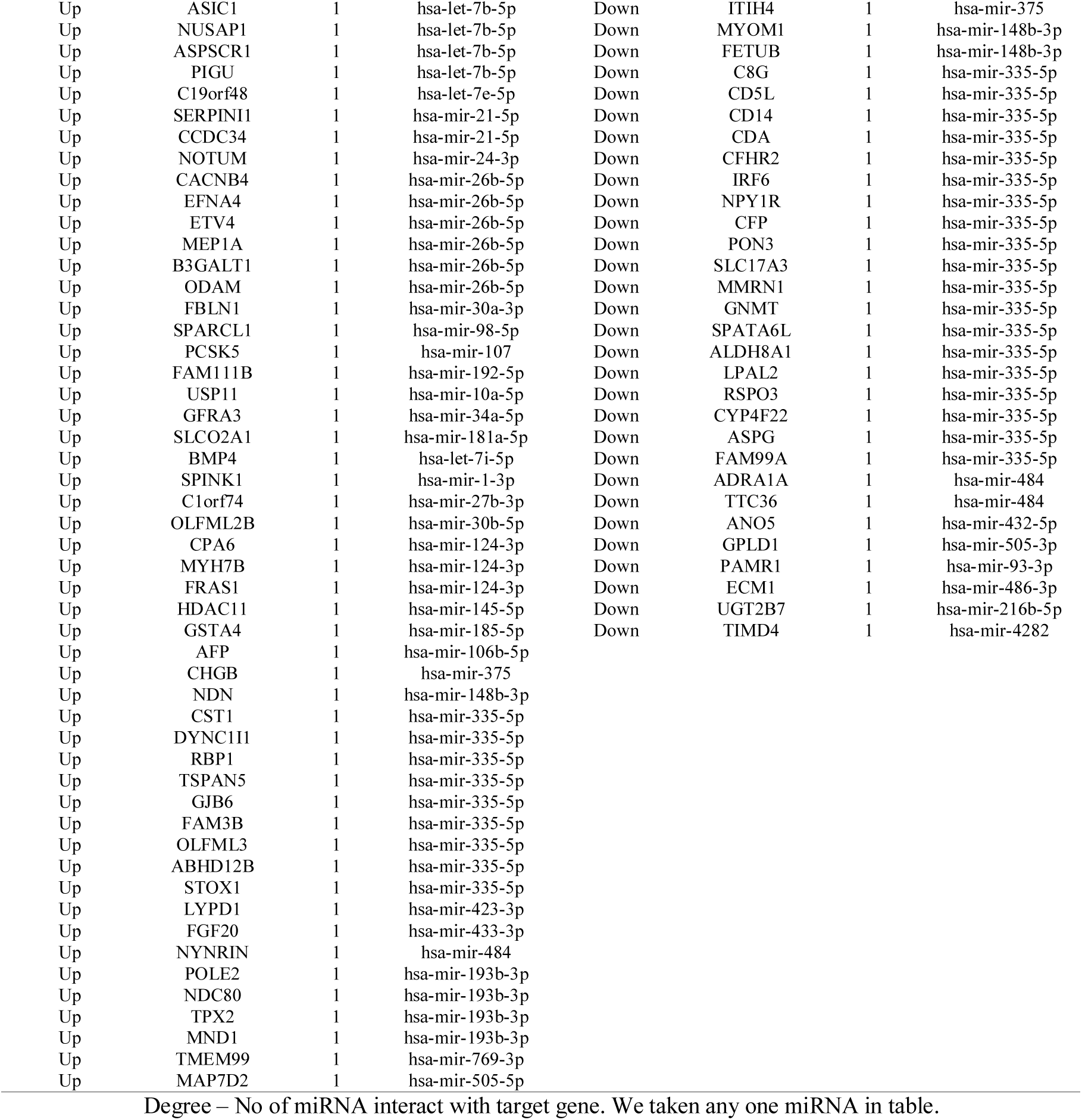
miRNA - target gene interaction table

**Table 9.**
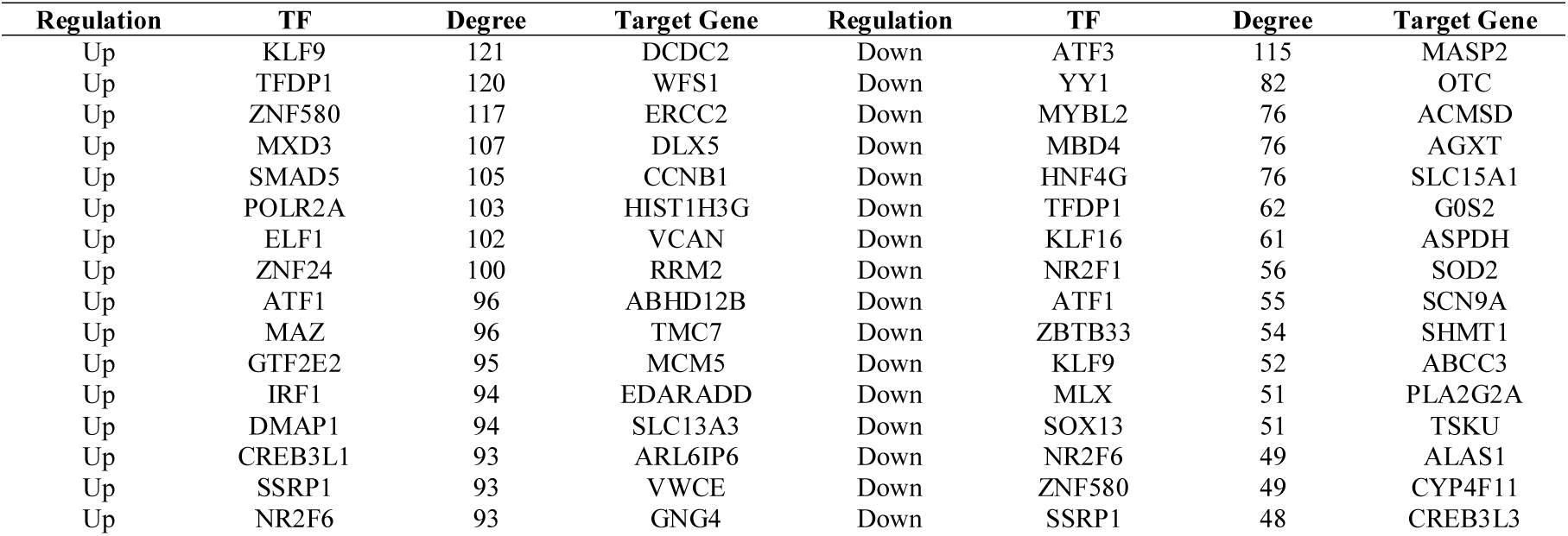
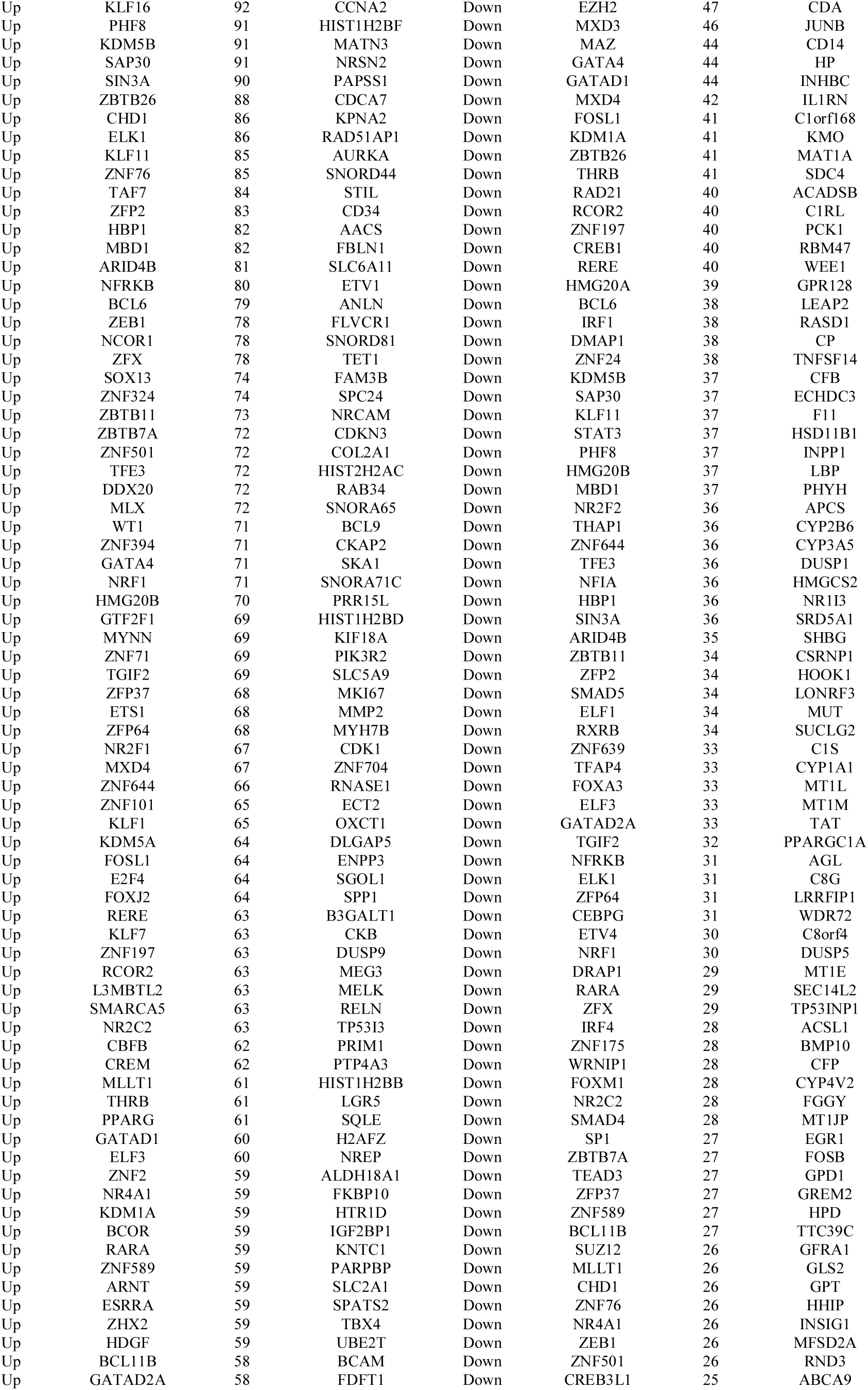
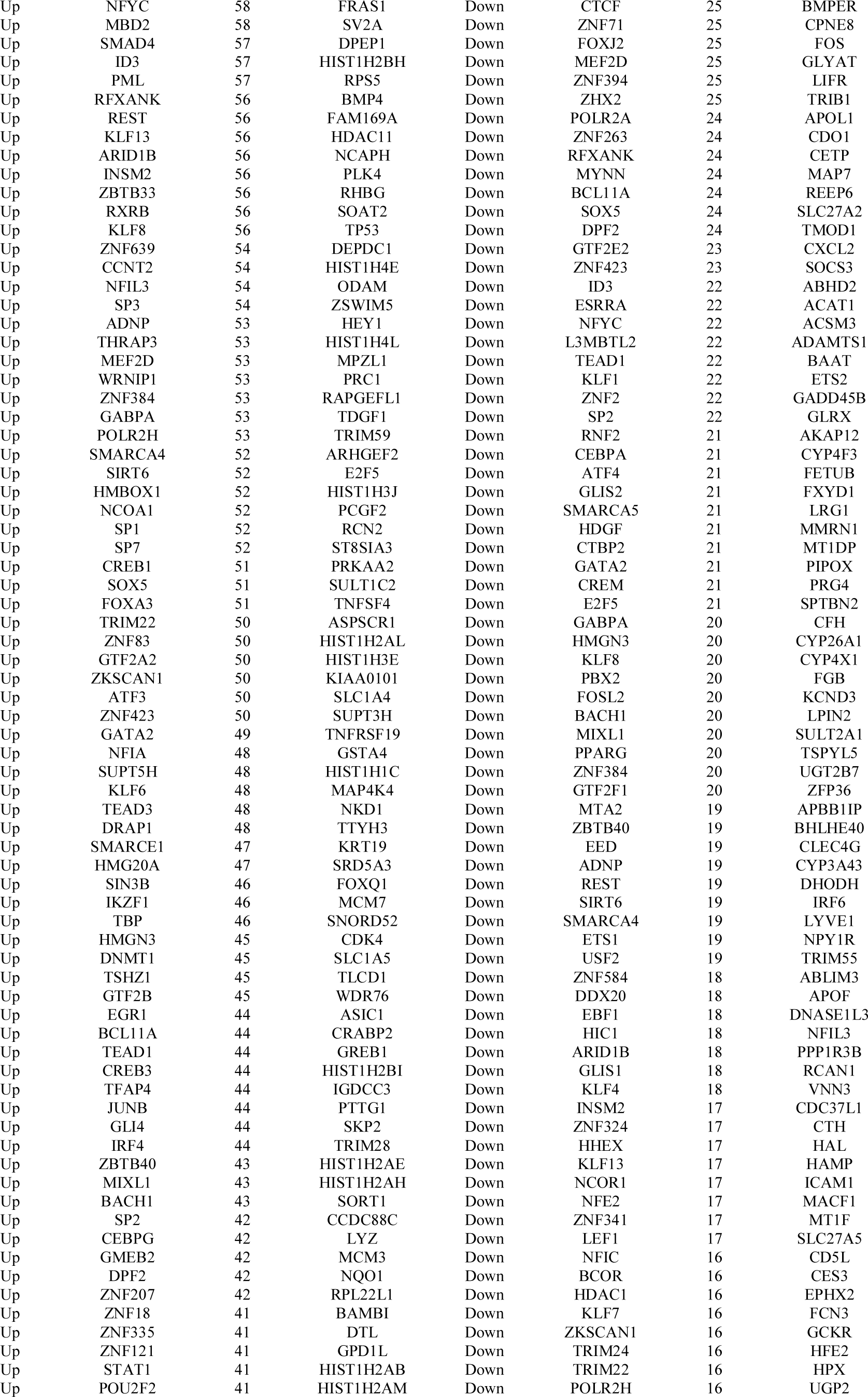
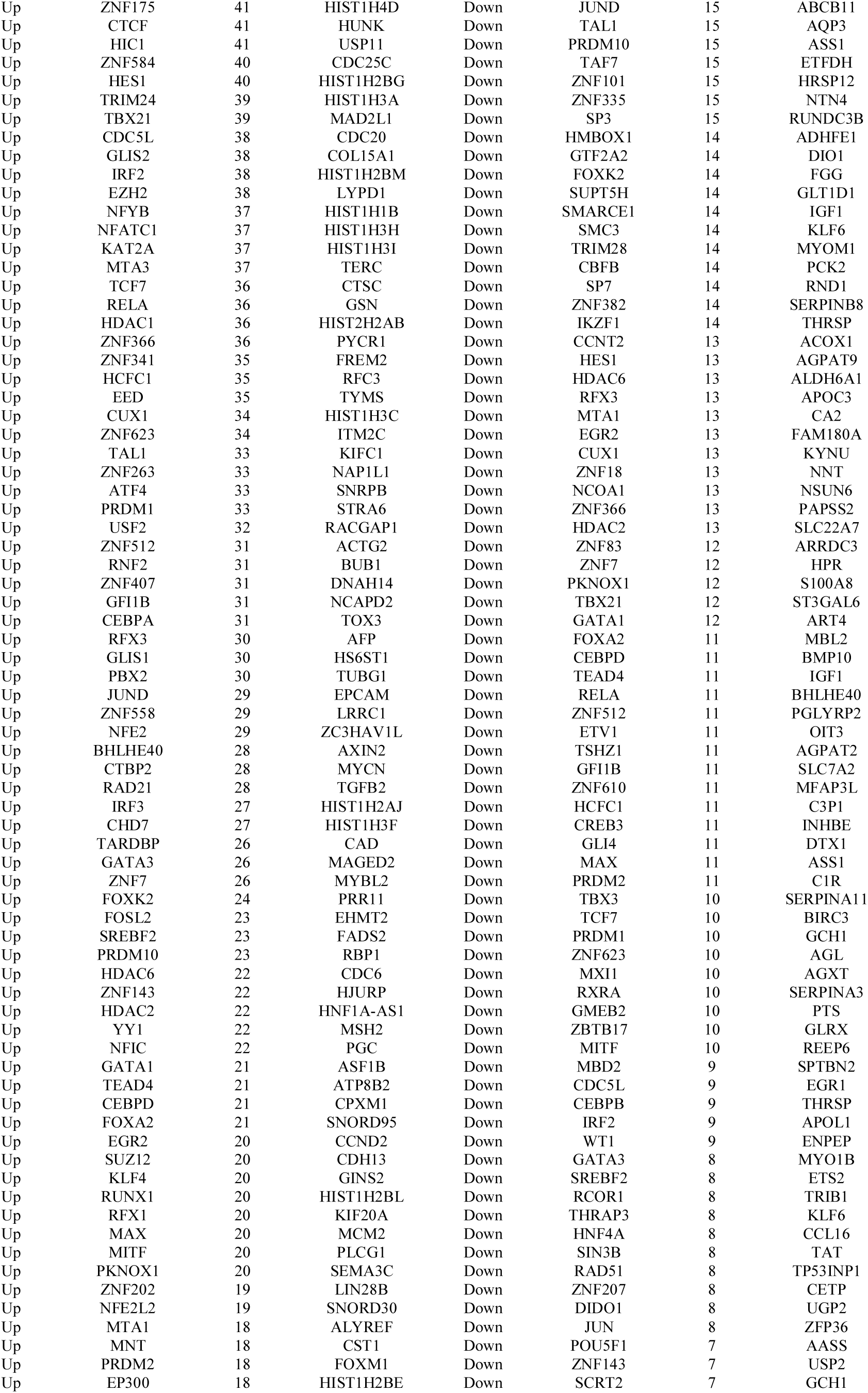
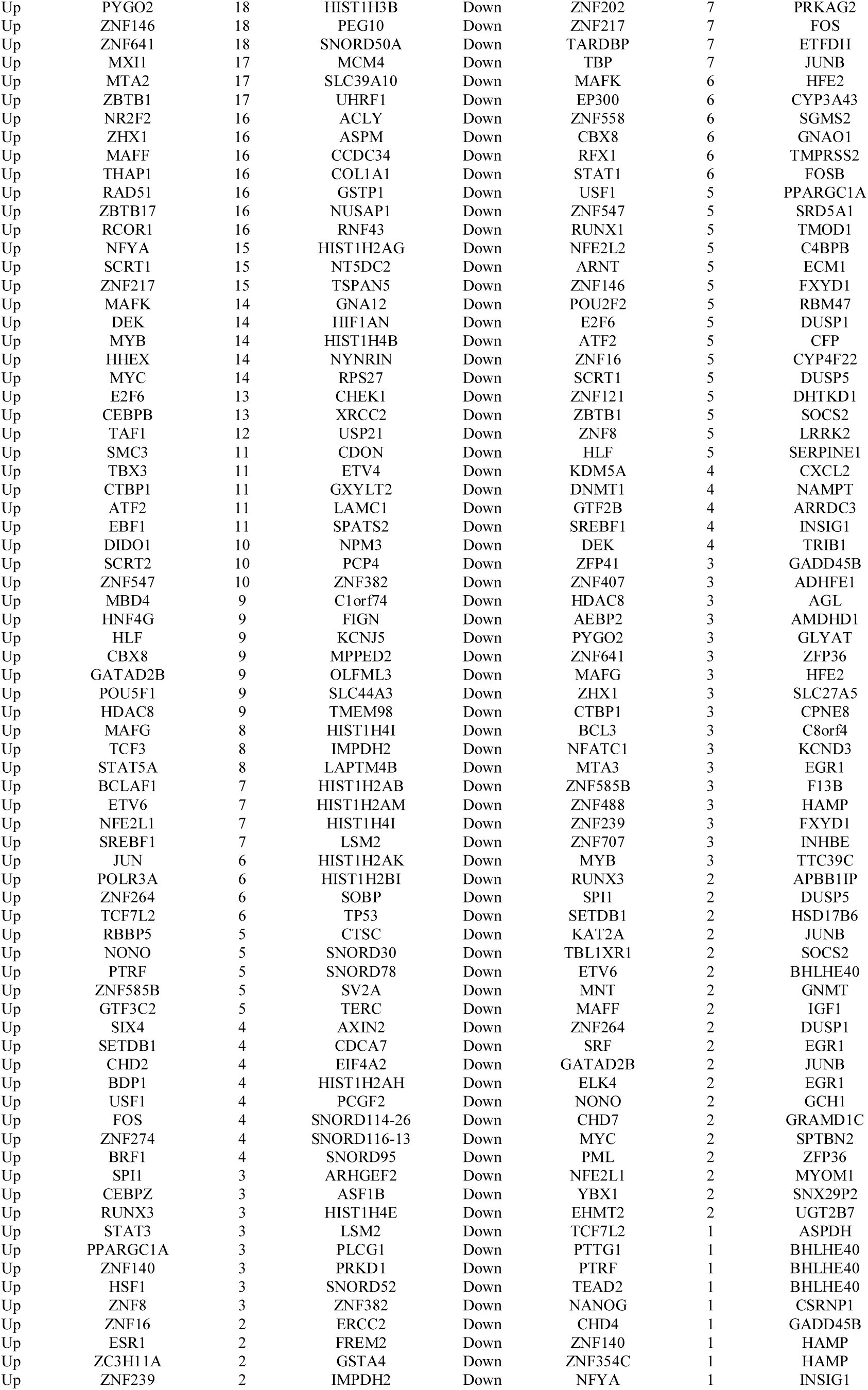
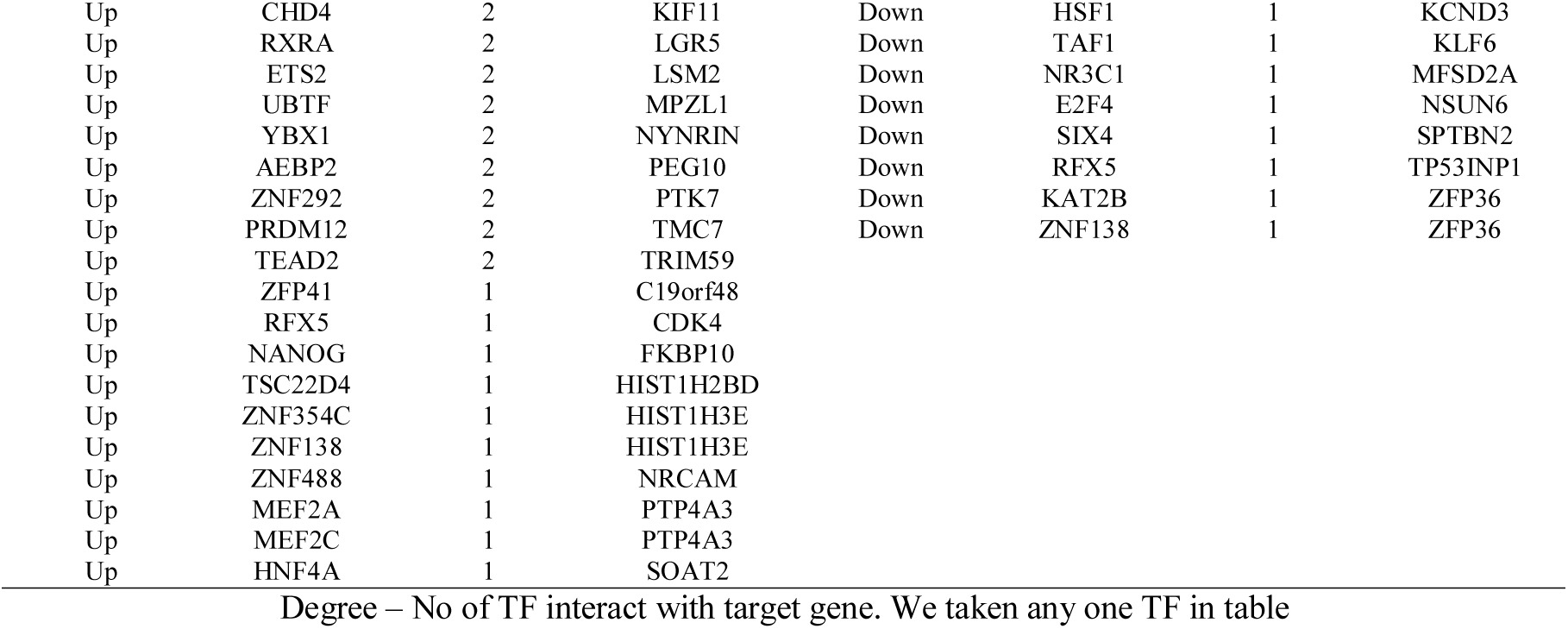
TF - target gene interaction table

### Validation of hub genes

To explore the prognostic role of up and down regulated hub genes, we analyzed the relationship between mRNA expression and overall survival (OS) using the UALCAN online tool. Interestingly, we found that high expression of TP53, PLK1, AURKA, CDK1 and ANLN were resulted in shorter OS in hepatoblastoma patients (Fig. 15), whereas low expression of ESR1, FGB, ACAT1, GOT1 and ALAS1 were resulted in shorter OS in hepatoblastoma patients (Fig. 16). To further validate the results, we examined the expression of ten hub genes using the UALCAN database. The expression levels of five up regulated hub genes (TP53, PLK1, AURKA, CDK1 and ANLN) and five down regulated hub genes (ESR1, FGB, ACAT1, GOT1 and ALAS1) were significantly different between hepatoblastoma samples and noncancerous liver samples (Fig. 17). The expression trends of the ten hub genes were consistent with the results we obtained. The stage analyses of TP53, PLK1, AURKA, CDK1, ANLN, ESR1, FGB, ACAT1, GOT1 and ALAS1 in human tissues were visualized using UALCAN online tool. The present study revealed that TP53, PLK1, AURKA, CDK1 and ANLN exhibited higher expression levels in all stage of hepatoblastoma compared with the noncancerous liver samples (Fig. 18 A-E), whereas ESR1, FGB, ACAT1, GOT1 and ALAS1 exhibited lower expression levels in all stage of hepatoblastoma compared with the noncancerous liver samples (Fig. 18 F-J). We explored the genetic mutations of up and down regulated hub genes using the cBioPortal tool in TCGA data and found that TP53 (32%), PLK1 (0.3%), AURKA (2%), CDK1 (0.6%), ANLN (1.7%), ESR1 (2.8%), FGB (2%), ACAT1 (1.1%), GOT1 (0.8%) and ALAS1 (1.1%) were the most frequently muted genes in the ten query genes, including inframe mutation (putative driver), missense mutation (putative driver), truncating mutation (putative driver), deep deletion, missense mutation (unknown significance), amplification, truncating mutation (unknown significance) and fusion (Fig. 19). Based on the immunohistochemical staining results from HPA database, the protein expression level of TP53, PLK1, AURKA, CDK1 and ANLN was consistent with their gene expression, that is, the protein levels of hub genes were also in a higher expression state in hepatoblastoma tissues compared to noncancerous liver (Fig. 20 A-E), whereas the protein expression level of ESR1, FGB, ACAT1, GOT1 and ALAS1 was consistent with their gene expression, that is, the protein levels of hub genes were also in a lower expression state in hepatoblastoma tissues compared to noncancerous liver (Fig. 20 F-J). ROC curve analysis revealed that five up regulated hub genes (TP53, PLK1, AURKA, CDK1 and ANLN) and five down regulated hub genes (ESR1, FGB, ACAT1, GOT1 and ALAS1) serve valuable biomarkers for distinguishing hepatoblastoma from those with noncancerous liver, with the AUC exceeded 0.90 (Fig. 21). For up regulated genes, the AUC was 0.934 for TP53, 0.973 for PLK1, 0.956 for AURKA, 0.976 for CDK1 and 0.942 for ANLN. For down regulated genes, the AUC was 0.950 for ESR1, 0.950 for FGB, 0.957 for ACAT1, 0.972 for GOT1 and 0.933 for ALAS1. Consistent with the bioinformatics analysis results, the RT-PCR results revealed. As shown in Fig. 22, the expression levels of TP53, PLK1, AURKA, CDK1 and ANLN were significantly higher in the hepatoblastoma than in the noncancerous liver, whereas expression levels of ESR1, FGB, ACAT1, GOT1 and ALAS1 were significantly lower in the hepatoblastoma than in the noncancerous liver. Immune infiltration analysis was found that the high expression level of TP53, PLK1, AURKA, CDK1 and ANLN were all negatively associated with tumor purity (Fig. 23 A-E), whereas low expression level of ESR1, FGB, ACAT1, GOT1 and ALAS1 were all positively associated with tumor purity (Fig. 23 F-J). This finding further confirmed the key role of these up and down regulated hub genes probably expressed not in the microenvironment, but expressed in the tumor cells. These findings further confirmed the key role of these up and down regulated hub genes in the onset of hepatoblastoma.

**Fig. 15.**
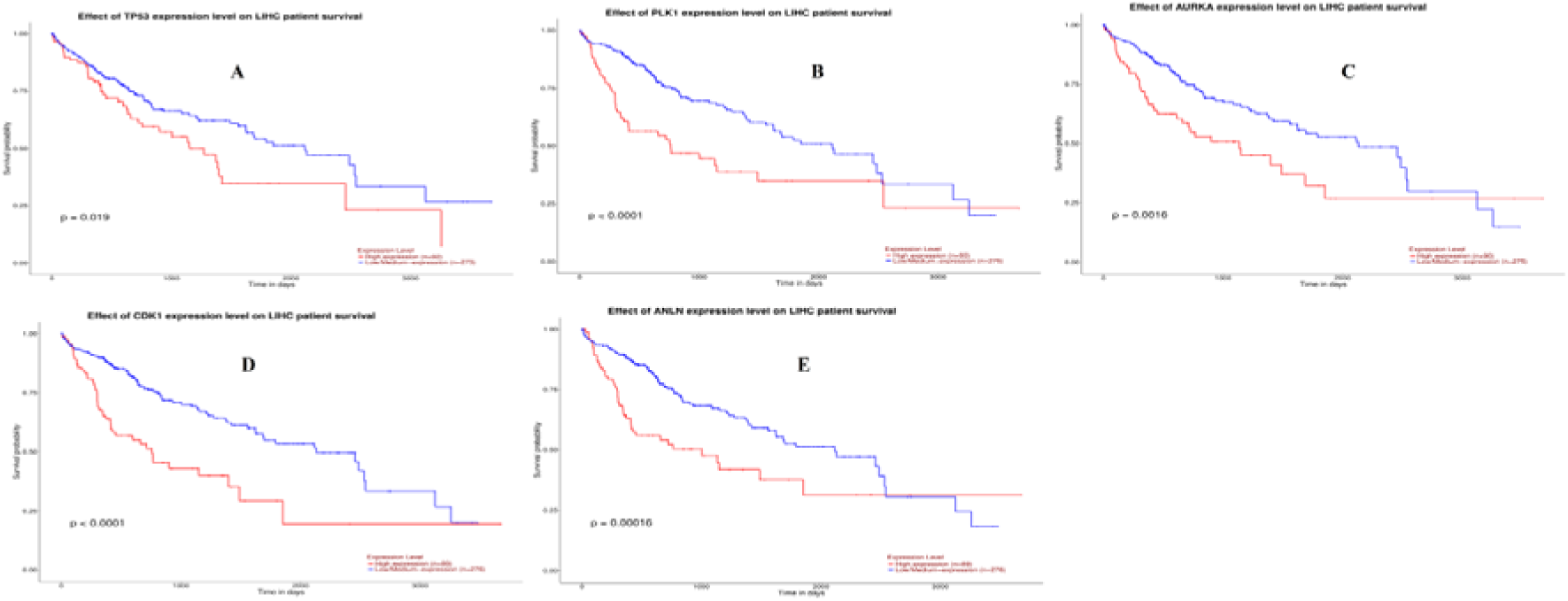
Overall survival analysis of up regulated hub genes. Overall survival analyses were performed using the UALCAN online platform. Red line denotes - high expression; Blur line denotes – low expression. A) TP53 B) PLK1 C) AURKA D) CDK1 E) ANLN

**Fig. 16.**
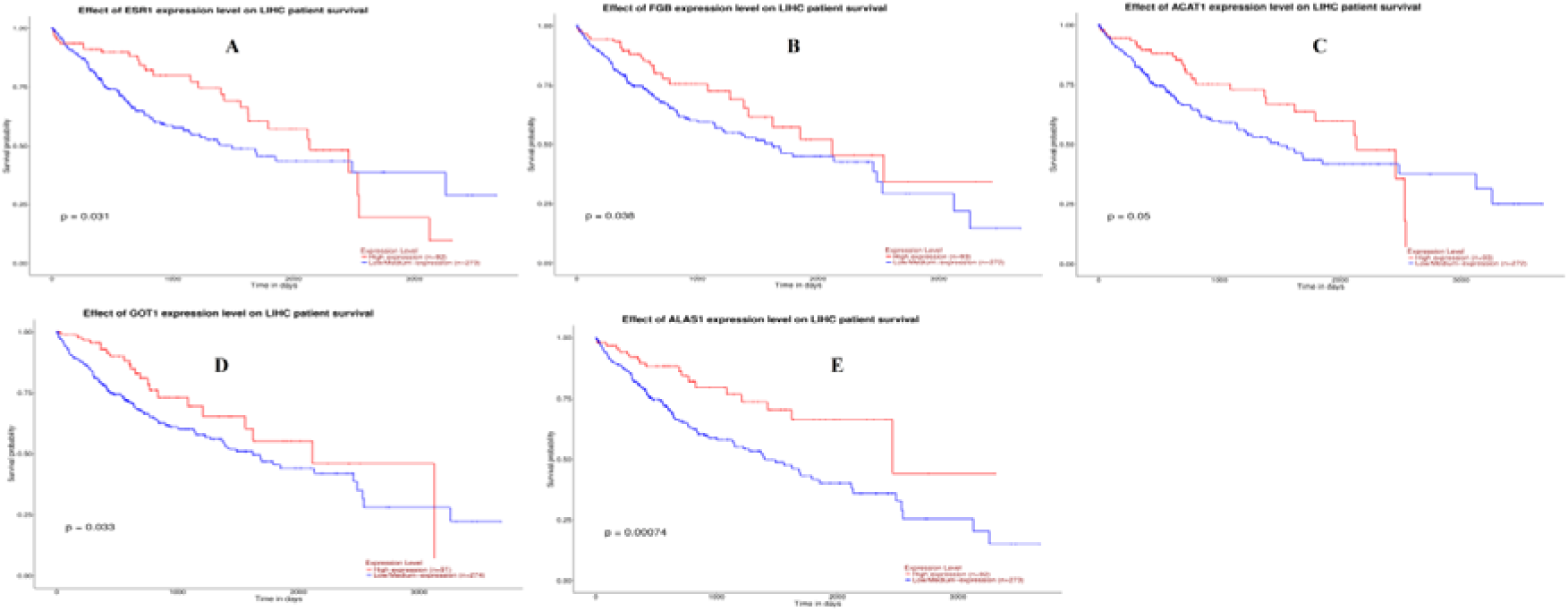
Overall survival analysis of down regulated hub genes. Overall survival analyses were performed using the UALCAN online platform. Red line denotes - high expression; Blur line denotes – low expression. A) ESR1 B) FGB C) ACAT1 D) GOT1 E) ALAS1

**Fig. 17.**
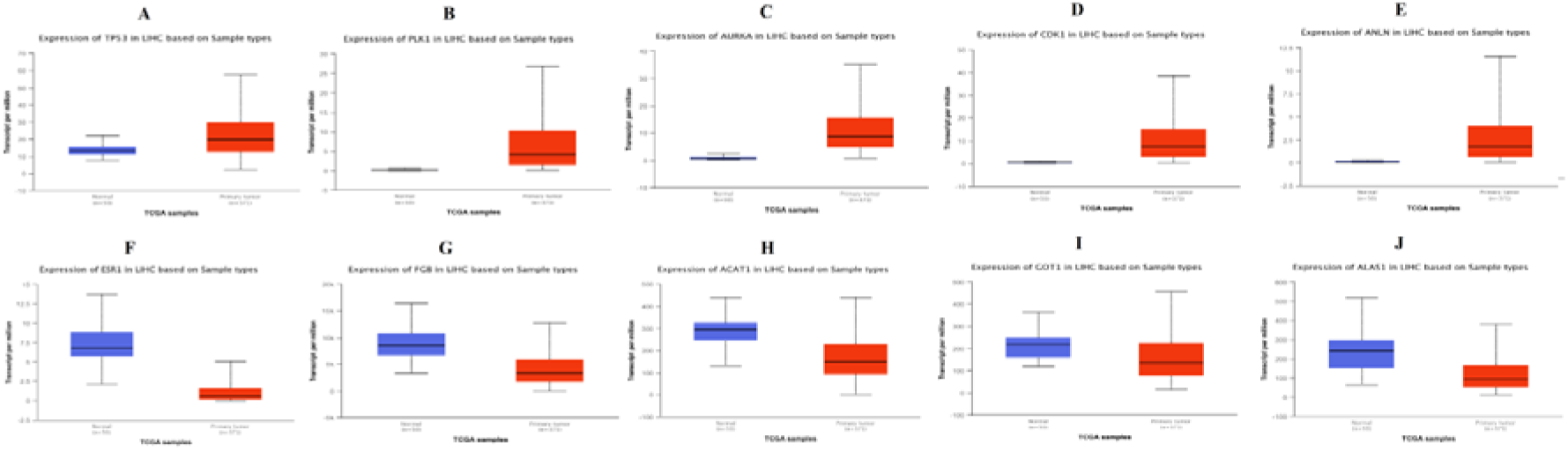
Box plots (expression analysis) hub genes (up and down regulated) were produced using the UALCAN platform. A) TP53 B) PLK1 C) AURKA D) CDK1 E) ANLN F) ESR1 G) FGB H) ACAT1 I) GOT1 G) ALAS1

**Fig. 18.**
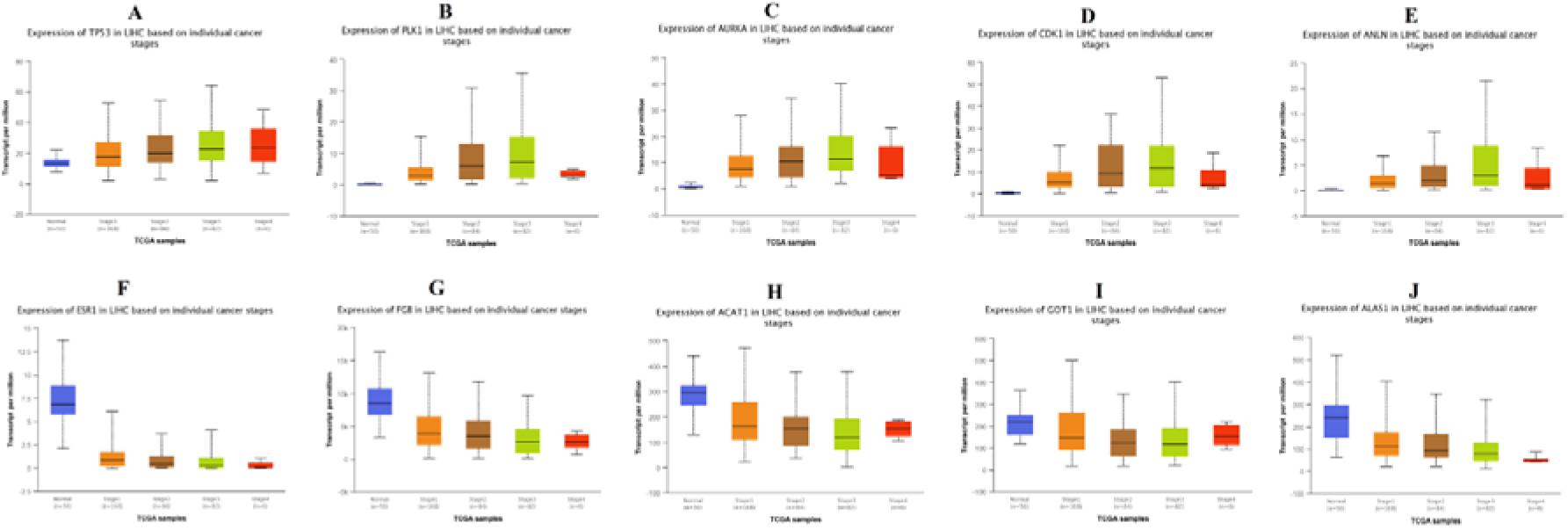
Box plots (stage analysis) of hub genes (up and down regulated) were produced using the UALCAN platform A) TP53 B) PLK1 C) AURKA D) CDK1 E) ANLN F) ESR1 G) FGB H) ACAT1 I) GOT1 G) ALAS1

**Fig. 19.**
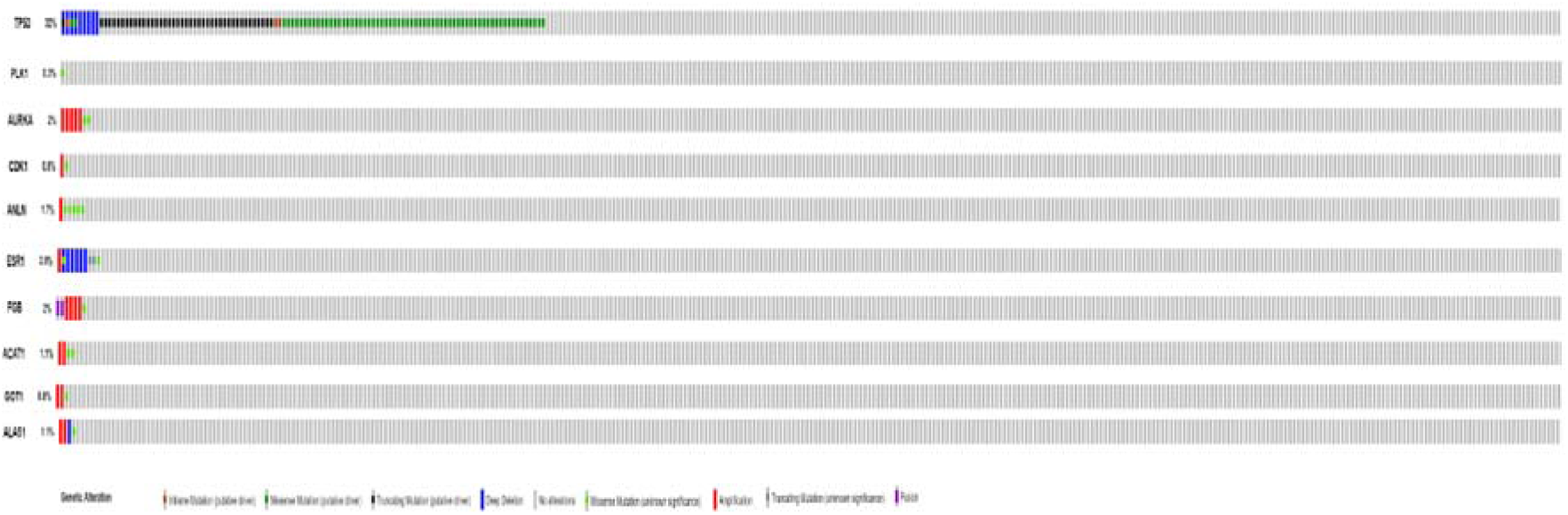
Mutation analyses of hub genes were produced using the CbioPortal online platform

**Fig. 20.**
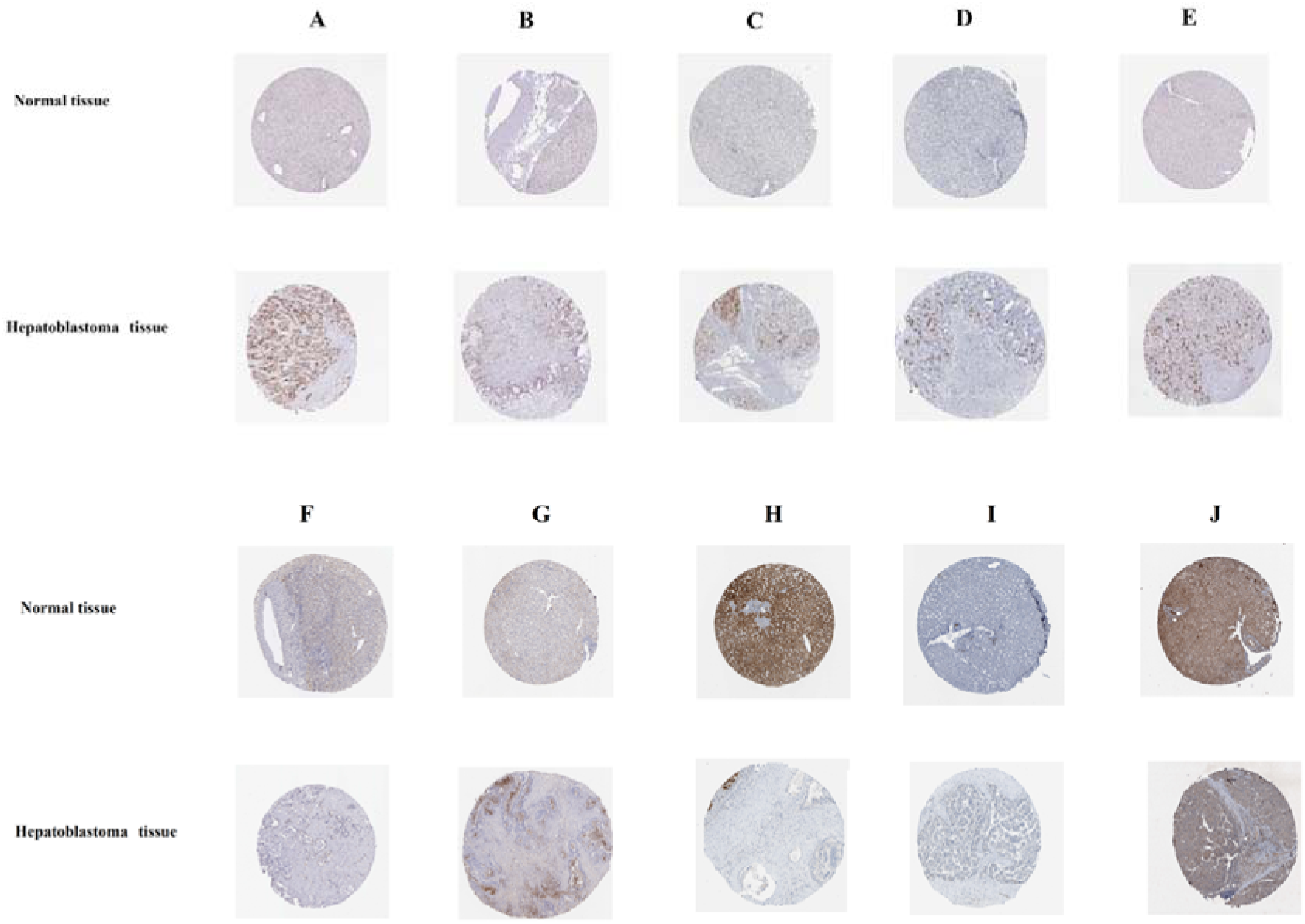
Immunohisto chemical(IHC) analyses of hub genes were produced using the human protein atlas (HPA) online platform A) TP53 B) PLK1 C) AURKA D) CDK1 E) ANLN F) ESR1 G) FGB H) ACAT1 I) GOT1 G) ALAS1

**Fig. 21.**
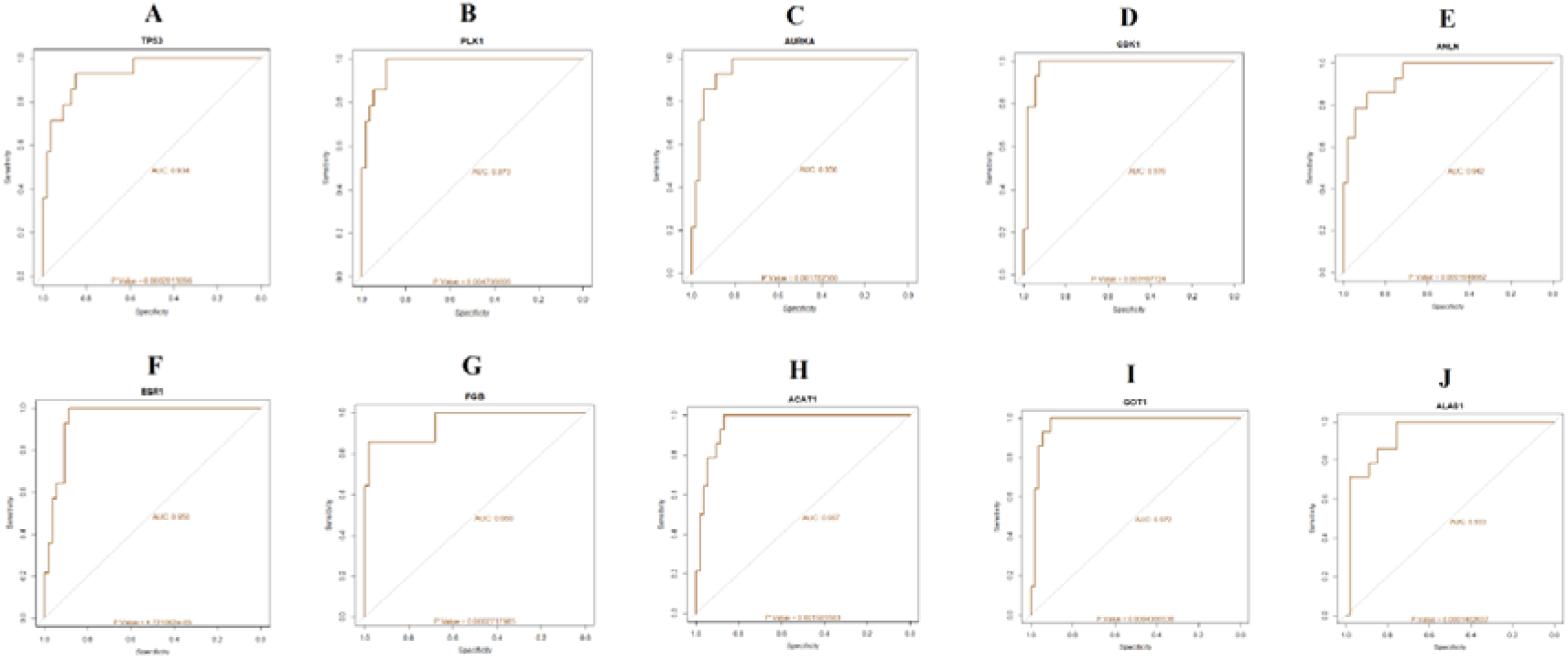
ROC curve validated the sensitivity, specificity of hub genes as a predictive biomarker for hepatoblastoma prognosis A) TP53 B) PLK1 C) AURKA D) CDK1 E) ANLN F) ESR1 G) FGB H) ACAT1 I) GOT1 G) ALAS1

**Fig. 22.**
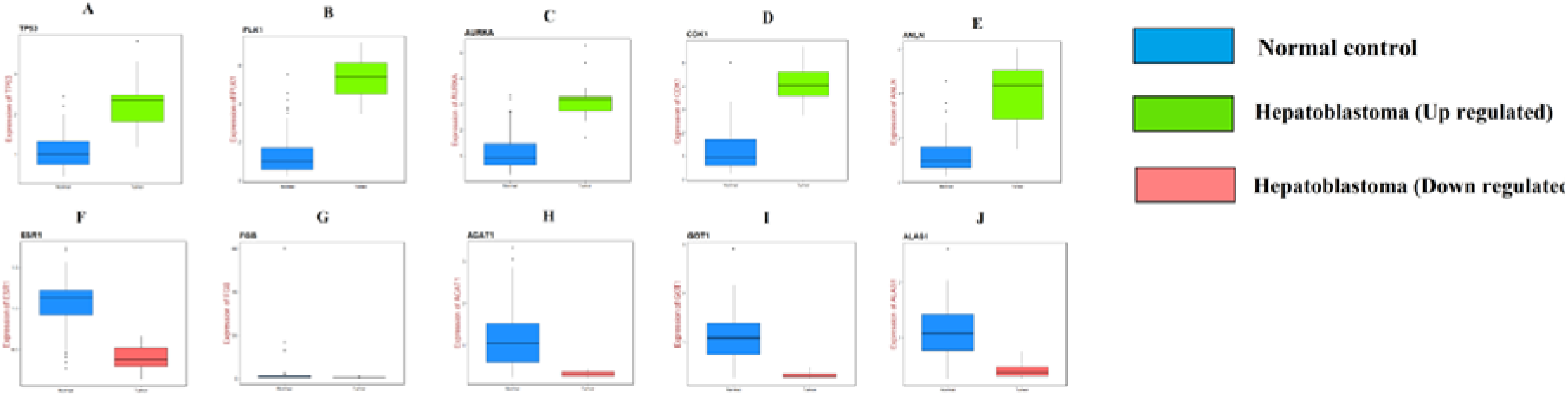
Validation of hub genes (up and down regulated) by RT- PCR. A) TP53 B) PLK1 C) AURKA D) CDK1 E) ANLN F) ESR1 G) FGB H) ACAT1 I) GOT1 G) ALAS1

**Fig. 23.**
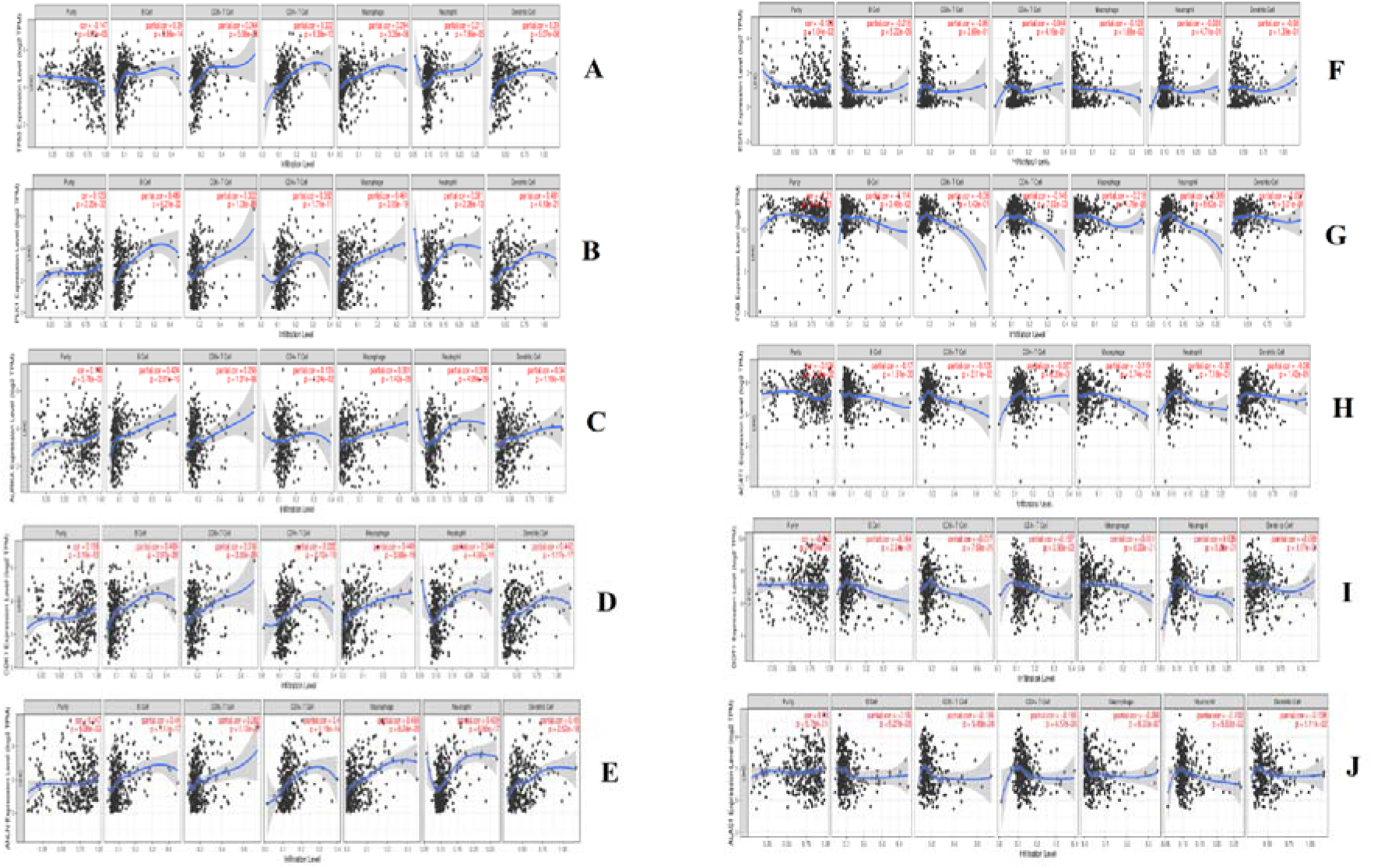
Scatter plot for immune infiltration for hub genes (up and down regulated). A) TP53 B) PLK1 C) AURKA D) CDK1 E) ANLN F) ESR1 G) FGB H) ACAT1 I) GOT1 G) ALAS1

## Discussion

Presently, the speedy forward in microarray and high-throughput technologies has spread the function biomedicine in clinical practice, such as cancer initial diagnosis, new targeted drug exploration and prognosis prediction. GEO database, as a public repository for archiving high-throughput microarray experimental gene expression data, has contributed the dynamic tools to resolve important genes and pathways linked with the molecular pathogenesis of tumors [Wu et al. 2019]. In the current investigation, based on the GEO database, single gene expression profile including 53 hepatoblastoma tissues samples and 14 noncancerous liver tissue samples were integrated for a comprehensive bioinformatics analysis. The aim of our investigation was to identify the possible small molecule drugs for the treatment of hepatoblastoma and to determine the new biomarkers correspond with the molecular pathogenesis and prognosis of hepatoblastoma. A total of 996 DEGs between hepatoblastoma tissues and corresponding noncancerous liver tissues were diagnosed, which consisted of 499 up regulated genes and 497 down regulated genes. Genes such as DLK1 [Falix et al. 2012] and EPCAM (epithelial cell adhesion molecule) [Lopez-Nunez et al. 2019] were linked with development of hepatoblastoma. Genes such as NKD1 [Wang et al. 2017] and TNFRSF19 [Guo et al. 2020] were liable for proliferation of hepatocellular carcinoma cells, but these genes may be linked with development of proliferation of hepatoblastoma cells. NPNT (nephronectin) was associated with invasion of breast cancer cells [Wang et al. 2018], but this gene may be responsible for invasion of hepatoblastoma cells. Decrease expression of tumour suppressor genes such as C3P1 [Zhong et al. 2018], SLC22A1 [Grimm et al. 2016], RDH16 [Zhu et al. 2020], HAO2 [Mattu et al. 2016] and GLS2 [Kuo et al. 2016] were linked with progression of hepatocellular carcinoma, but low expression of these genes may associated with invasion of hepatoblastoma cells.

Pathway enrichment analyses were performed using the 499 up regulated genes to elucidate the action of hepatoblastoma. PYCR1 was associated with metabolism in hepatocellular carcinoma [Zhuang et al. 2019], but this gene may be liable for linked with metabolism in hepatoblastoma. Enriched genes such as ALDH18A1 [Guo et al. 2020] and HDAC11 [Thole et al. 2017] were linked with development of neuroblastoma, but these genes may be involved in progression of hepatoblastoma. Enriched methylated enriched genes such as GNG4 [Pal et al. 2016], CCND2 [Tsutsui et al. 2010], CDH13 [Moelans et al. 2011] and HELLS (helicase, lymphoid specific) [Law et al. 2019] were liable for progression of various cancer types, but these methylated genes may be important for advancement of hepatoblastoma. Enriched genes such as KIF18A [Luo et al. 2018], MCM6 [Liu et al. 2018], SKA1 [Xiao et al. 2019] and ACTG2 [Wu et al. 2017] were involved in invasion of hepatocellular carcinoma cells, but these genes may be associated with invasion of hepatoblastoma cells. Enriched genes such as PTTG1 [Lin et al. 2019], CENPF (centromere protein F) [Li et al. 2018], HJURP (Holliday junction recognition protein) [Chen et al. 2018], MCM2 [Yang et al. 2018], MCM7 [Qu et al. 2017], RRM2 [Wang et al. 2016], SGO1 [Wang et al. 2015], NDC80 [Ju et al. 2017], FOXM1 [Yu et al. 2016], SKP2 [Wei et al. 2018], TPX2 [Chen et al. 2018], PLK4 [Bao et al. 2018], NUF2 [Wang et al. 2019], CENPK (centromere protein K) [Wang et al. 2019], NCAPG (non-SMC condensin I complex subunit G) [Zhang et al. 2018], BUB1 [Xu et al. 2017], E2F5 [Sun et al. 2016], RFC3 [Yao et al. 2015], NCAPH (non-SMC condensin I complex subunit H) [Sun et al. 2019], CCNB1 [Gu et al. 2019], CDK1 [Gao et al. 2018], CDC25C [Sun et al. 2015], MAD2L1 [Li et al. 2017], CDK6 [Zhu et al. 2016], DKK1 [Zhang et al. 2016], DKK4 [Chouhan et al. 2016], DKK2 [Lin et al. 2016] and LEF1 [Hu et al. 2018] were important for proliferation of hepatocellular carcinoma cells, but these genes may be involved in proliferation of hepatoblastoma cells. High expression of enriched genes such as SPC25 [Chen et al. 2018], MCM4 [Xie et al. 2017], TYMS (thymidylatesynthetase) [Lee et al. 2013], KNTC1 [Liu et al. 2019] and CDH11 [Chen et al. 2018] were responsible for advancement of various cancer types, but over expression these gene may be liable for progression of hepatoblastoma. Elevated expression of enriched genes such as MCM3 [Meng et al. 2019], CHEK1 [Gong et al. 2020], SPC24 [Zhu et al. 2015], MYBL2 [Guan et al. 2018], KIF20A [Lu et al. 2018], CCNB2 [Li et al. 2019], CDC20 [Zhuang et al. 2018], SQLE (squaleneepoxidase) [Liu et al. 2018], ECT2 [Yu et al. 2019], SFRP4 [Xu et al. 2015], FRZB (frizzled related protein) [Shen et al. 2015] and BCL9 [Xu et al. 2017] were associated with advancement of hepatocellular carcinoma, but over expression of these genes may be linked with development of hepatoblastoma. Enriched genes such as MCM5 [Mio et al. 2016], PRIM1 [Lee et al. 2019], GINS2 [Chi et al. 2020], GINS1 [Tang et al. 2019] and CCNA2 [Li et al. 2019] were responsible for proliferation of various cancer types, but these genes may be linked with proliferation of hepatoblastoma cells. Mutation in TUBG1 was liable for advancement of breast cancer [Blanco et al. 2015], but alteration in this gene may be liable for progression of hepatoblastoma. Enriched genes such as PLK1 [Yamada et al. 2004], AURKA (aurora kinase A) [Zhang et al. 2018], TP53 [Yang et al. 2019], CDK4 [Kim et al. 1998] and AXIN2 [Koch et al. 2004] were involved in development of hepatoblastoma. Enriched genes such as POLE2 [Li et al. 2018], CENPI (centromere protein I) [Ding et al. 2018] and SPDL1 [Kodama et al. 2019] were responsible for invasion of various cancer cell types, but these genes may be involved in invasion of hepatoblastoma cells. Single nucleotide polymorphism (SNP) in CDC6 was liable for advancement of hepatocellular carcinoma [Xiong et al. 2009], but this polymorphic gene may be involved in development of hepatoblastoma. Our study found that H4C9, H2AC15, H2AC14, H2AC16, H2AC6, H2AC4, H2AC17, H2AC20, H2BC8, H2BC13, H2BC14, H2BC7, H2BC6, H2BC9, H2BC10, H3C1, H3C3, H3C6, H3C11, H3C8, H3C12, H3C10, H3C2, H4C4, H4C2, H4C5, H4C13, H4C14, MACROH2A2, H2AC12, H2AC21, H3C7, H2AC11, H2AC8, H2AZ1, H2BC5, H2BC3, MND1, RPS27, NCAPD2, RHNO1, FDFT1, MYH7B, MYCN (MYCN proto-oncogene, bHLH transcription factor) and SLC7A6 are up regulated in hepatoblastoma and has potential as a novel diagnostic and prognostic biomarker, and therapeutic target. Similarly, pathway enrichment analyses were performed using the 497 down regulated genes to elucidate the action of hepatoblastoma. SNP in enriched genes such as ADH1B [Shih et al. 2018], NAT2 [Farker et al. 2003], CYP1A1 [Abo-Hashem et al. 2016], CYP2D6 [Agundez et al. 1996], GSTA1 [Akhdar et al. 2016], MBL2 [Wang et al. 2016] C6 [Wang et al. 2016] and SERPINE1 [Divella et al. 2015] were linked with advancement of hepatocellular carcinoma, but these polymorphic genes may be responsible for progression of hepatoblastoma. Enriched genes such as KMO (kynurenine 3-monooxygenase) [Jin et al. 2015], HMGCS2 [Wang et al. 2019], LIPG (lipase G, endothelial type) [Cadenas et al. 2019], PON3 [Cai et al. 2016], AASS (aminoadipate-semialdehyde synthase) [Xue et al. 2019], PCK2 [Liu et al. 2012], PPARGC1A [Wang et al. 2018] and JUNB (JunB proto-oncogene, AP-1 transcription factor subunit) [Kim et al. 2001] were associated with proliferation of hepatocellular carcinoma cells, but these genes may be associated with proliferation of hepatoblastoma cells. Enriched genes such as CYP1A2 [Biazi et al. 2017], ADH4 [Wei et al. 2012], ACSL1 [Cui et al. 2014], UGT2B4 [Wijayakumara et al. 2017], ALDH1A1 [Yan et al. 2016], PON1 [Shu et al. 2017], ASS1 [Frulio et al. 2019], AKR1D1 [Nikolaou et al. 2019], PTGS2 [Chen et al. 2019], NNMT (nicotinamide N-methyltransferase) [Kim et al. 2009], GPT (glutamic--pyruvic transaminase) [Shimokawa et al. 1977], NNT (nicotinamide nucleotide transhydrogenase) [Lu et al. 2017] and PCK1 [Xiang et al. 2020] were linked with metabolic activity in hepatocellular carcinoma, but these genes may be involved in metabolic activity in hepatoblastoma. Low expression of enriched genes such as CYP2C19 [Ashida et al. 2018], ABAT (4-aminobutyrate aminotransferase) [Jansen et al. 2015], ACOX1 [Chen et al. 2018], ACSM3 [Ruan et al. 2017], FBP1 [Liu et al. 2018], ST3GAL6 [Souady et al. 2011], AGXT (alanine--glyoxylate and serine-- pyruvate aminotransferase) [Sun et al. 2019], ALDOB (aldolase, fructose-bisphosphate B) [Tao et al. 2015], ALDH6A1 [Shin et al. 2020], XDH (xanthine dehydrogenase) [Chen et al. 2017], OGDHL (oxoglutarate dehydrogenase like) [Jiao et al. 2019], CYP2C8 [Li et al. 2019], CYP2C9 [Yu et al. 2015], CYP3A5 [Jiang et al. 2015], BHMT (betaine--homocysteine S-methyltransferase) [Jin et al. 2016], TAT (tyrosine aminotransferase) [Fu et al. 2010], SLC27A5 [Gao et al. 2020], GYS2 [Chen et al. 2019], GLYAT (glycine-N-acyltransferase) [Matsuo et al. 2012], CES3 [Quiroga et al. 2016], MASP2 [Ding et al. 2014], C7 [Seol et al. 2016] and F11 [Du et al. 2019] were involved in progression of hepatocellular carcinoma, but decrease expression these genes may be identified with growth of hepatoblastoma. Enriched genes such as SULT1A2 [Fernandez-Santander et al. 2013], PIK3C2G [Li et al. 2015], AGL (amylo-alpha-1, 6-glucosidase, 4-alpha-glucanotransferase) [Richmond et al. 2018], UGT2B10 [Lu et al. 2018], CTH (cystathionine gamma-lyase) [Xu et al. 2020] and CYP26A1 [Osanai and Lee, 2015] were involved in metabolic activity of various cancer types, but these genes may liable for metabolic activity of in hepatoblastoma. Enriched genes such as TDO2 [Pham et al. 2018], EPHX2 [Vainio et al. 2011], PLA2G2A [Ganesan et al. 2008], INPP1 [Li et al. 2019], GCH1 [Gao et al. 2016], GNE (glucosamine (UDP-N-acetyl)-2-epimerase/N-acetylmannosamine kinase) [Kemmner et al. 2012], PAPSS2 [Zhang et al. 2019], GLYATL1 [Eich et al. 2019], CFB (complement factor B) [Kim et al. 2019], C1R [Riihilä et al. 2020], C1S [Riihilä et al. 2020], C9 [Chong et al. 2010], FGB (fibrinogen beta chain) [Repetto et al. 2018] and KNG1 [Quesada-Calvo et al. 2017] were linked with development of various cancer types, but these genes may be culpable for advancement of hepatoblastoma. Methylation inactivation of tumor suppressor enriched genes such as HAAO (3-hydroxyanthranilate 3,4-dioxygenase) [Huang et al. 2010] and AOX1 [Vantaku et al. 2019] were identified with growth of various cancer types, but loss of these genes may be liable for progression of hepatoblastoma. Epigenetic inactivation of enriched tumor suppressor genes such as CDO1 [Choi et al. 2017], DUSP5 [Liu et al. 2017] and SOCS3 [Zhang et al. 2015] were liable for progression of hepatocellular carcinoma, but loss of these genes may be linked with development of hepatoblastoma. Enriched genes such as SGMS2 [Zheng et al. 2019], ACAT1 [Ye et al. 2016], UGP2 [Zeng et al. 2019], GOT1 [Wang et al. 2019], CDA (cytidinedeaminase) [Ye et al. 2015] and FOS (Fos proto-oncogene, AP-1 transcription factor subunit) [Liu et al. 2018] were important for proliferation of various cancer cells types, but these genes may be involved in proliferation of hepatoblastoma cells. SNP in enriched genes such as MAT1A [Mumbrekar et al. 2017], ADH1C [Hidaka et al. 2015], UGT2B7 [Sutiman et al. 2016], HSD11B1 [Wang et al. 2013], CYP2B6 [Kuo et al. 2017], CHST9 [Yuan et al. 2017], CYP39A1 [Melchardt et al. 2015], FMO3 [Bae et al. 2006], CYP3A43 [Han et al. 2015], CYP4F3 [Yin et al. 2017], CYP4F11 [Cardenas-Rodriguez et al. 2012], SULT2A1 [Wilborn et al. 2006], GSTA2 [Maekawa et al. 2011] and GSTA3 [Duan et al. 2018] were involved in progression of various cancer types, but these polymorphic genes may be linked with development of hepatoblastoma. Enriched genes such as MGLL (monoglyceride lipase) [Yang et al. 2018], DMGDH (dimethylglycine dehydrogenase) [Liu et al. 2016], SHMT1 [Dou et al. 2019], ARG1 [You et al. 2018], CXCL8 [Li et al. 2015] and FGG (fibrinogen gamma chain) [Zhang et al. 2019] were involved in invasion of hepatocellular carcinoma cells, but these genes may be responsible for invasion of hepatoblastoma cells. Enriched genes such as CYP3A4 [Holownia and Braszko, 2004], OTC (ornithine carbamoyltransferase) [Mavri-Damelin et al. 2007] and DUSP1 [Casteel et al. 2010] were associated with development of hepatoblastoma. GCLM (glutamate-cysteine ligase modifier subunit) was linked with drug resistance in lung cancer [Nishi et al. 2005], but this gene may be linked with chemoresitance in hepatoblastoma. Our study found that CYP4X1, KYNU (kynureninase), ACMSD (aminocarboxymuconatesemialdehyde decarboxylase), HGD (homogentisate 1,2-dioxygenase), ACACB (acetyl-CoA carboxylase beta), ACADL (acyl-CoA dehydrogenase long chain), ACADSB (acyl-CoA dehydrogenase short/branched chain), CES1, PIPOX (pipecolic acid and sarcosine oxidase), AKR1C4, HAO1, NDST3, HPD (4-hydroxyphenylpyruvate dioxygenase), SCP2, ALAS1, ALPL (alkaline phosphatase, biomineralization associated), AGPAT2, CYP4F2, ETNPPL (ethanolamine-phosphate phospho-lyase), GBA3, AGXT2, GDA (guanine deaminase), HSD17B6, LPIN2, MMUT (methylmalonyl-CoA mutase), UPB1, CYP2C18, MBOAT1, CYP4A11, CYP8B1, BAAT (bile acid-CoA:amino acid N-acyltransferase), GATM (glycine amidinotransferase), DAO (D-amino acid oxidase), UROC1, SUCLG2, PTS (6-pyruvoyltetrahydropterin synthase), DHODH (dihydroorotate dehydrogenase (quinone)), ACSM5, SDS (serine dehydratase), CNDP1, DPYS (dihydropyrimidinase), GPAT3, AMDHD1, NAMPT (nicotinamidephosphoribosyltransferase), EHHADH (enoyl-CoA hydratase and 3-hydroxyacyl CoA dehydrogenase), HAL (histidine ammonia-lyase), AADAT (aminoadipate aminotransferase), AADAC (arylacetamidedeacetylase), AKR7L, CYP4F22, SULT1B1, CYP4V2, CYP4A22, ACADM (acyl-CoA dehydrogenase medium chain), C5, C8A, F8, F9, F13B, SERPINC1 and KLKB1 are down regulated in hepatoblastoma and has potential as a novel diagnostic and prognostic biomarker, and therapeutic target.

GO enrichment analyses were performed using the 499 up regulated genes to elucidate the action of hepatoblastoma. Enriched genes such as NUSAP1 [Garrido-Rodríguez et al. 2019], ASF1B [Han et al. 2018] and PRKAA2 [Zhang et al. 2020] were associated with proliferation of various cancer types cells, but these genes may be linked with proliferation of hepatoblastoma cells. SNP in enriched genes such as ERCC2 [Zhao et al. 2019] and MSH2 [Zhu et al. 2018] were involved in progression of hepatocellular carcinoma, but these polymorphic genes may be linked with development of hepatoblastoma. Enriched genes such as TTK (TTK protein kinase) [Miao et al. 2016], MKI67 [Yang et al. 2017], TET1 [Lin et al. 2015], DLGAP5 [Liao et al. 2013] and SMYD2 [Zuo et al. 2018] were involved in proliferation of hepatocellular carcinoma cells, but these genes may be responsible for proliferation of hepatoblastoma cells. Over expression of USP21 was liable for progression of pancreatic cancer [Hou et al. 2019], but high expression of this gene may be linked with advancement of hepatoblastoma. Increased expression of enriched genes such as RACGAP1 [Wang et al. 2011], PRKD1 [Döppler et al. 2016], EHMT2 [Song et al. 2019], KIFC1 [Han et al. 2019], TERC (telomerase RNA component) [BaenaLDel et al. 2018], TRIM28 [Jin et al. 2017] and TOP2A [Panvichian et al. 2015] were involved in progression of hepatocellular carcinoma, but over expression these genes may be associated with advancement of hepatoblastoma. Enriched genes such as UHRF1 [Beck et al. 2018] and HMGA2 [Li et al. 2020] were responsible for advancement of hepatoblastoma. NAP1L1 was linked with chemoresistance in hepatocellular carcinoma [Le et al. 2019], but this gene may liable for drug resistance in hepatoblastoma. PRC1 was associated with invasion of hepatocellular carcinoma cell [Tang et al. 2019], but this gene may be liable for invasion of hepatoblastoma cells. Our study found that NPM3, SUPT3H, PCGF2, H1-2 (H1.2 linker histone, cluster member) and H1-5 (H1.5 linker histone, cluster member) are up regulated in hepatoblastoma and has potential as a novel diagnostic and prognostic biomarker, and therapeutic target. Similarly, GO enrichment analyses were performed using the 497 down regulated genes to elucidate the action of hepatoblastoma. Enriched genes such as HGF (hepatocyte growth factor) [Purcell et al. 2011], ABCB11 [Vilarinho et al. 2014] and TLR4 [Hsiao et al. 2015] were linked with advancement of hepatoblastoma. Enriched genes such as VNN3 [Ha et al. 2019], HPGD (15-hydroxyprostaglandin dehydrogenase) [He et al. 2016], VNN2 [Chen et al. 2018], VNN1 [Zhang et al. 2017], GPD1 [Zhou et al. 2017], CFH (complement factor H) [Campa et al. 2015], SERPINA3 [Cao et al. 2018], AFM (afamin) [Shen et al. 2016], HPR (haptoglobin-related protein) [Epelbaum et al. 1998], C4BPA [Sogawa et al. 2016], CYP4Z1 [Zheng et al. 2019], SRD5A1 [Sinreih et al. 2015] and STEAP4 [Xue et al. 2017] were linked with progression of various cancer types, but these genes may be involved in development of hepatoblastoma. Low expression of enriched genes such as ETFDH (electron transfer flavoprotein dehydrogenase) [Wu et al. 2019], GNMT (glycine N-methyltransferase) [Chen et al. 1998], DCN (decorin) [Ju et al. 2015], ITIH4 [Li et al. 2018], GHR (growth hormone receptor) [Qi et al. 2016], LYVE1 [Kitagawa et al. 2013] and HP (haptoglobin) [Mondal et al. 2016] were identified with development of hepatocellular carcinoma, but decrease expression of these genes may be associated with development of hepatoblastoma. SNP in PRKAG2 was responsible for advancement of colon and rectal cancer [Slattery et al. 2010], but this polymorphic gene may be liable for progression of hepatoblastoma. Enriched genes such as STAB2 [Wu et al. 2018], IGF1 [Shi and Teng, 2015], HRG (histidine rich glycoprotein) [Zhang et al. 2015], CFHR3 [Liu et al. 2020] and CD5L [Aran et al. 2018] were linked with proliferation of hepatocellular carcinoma cells, but these genes may be involved in proliferation of hepatoblastoma cells. Methylation inactivation of ADHFE1 was identified with growth of colorectal cancer [Hu et al. 2019], but loss this gene may be responsible for progression of hepatoblastoma. Enriched genes such as ANGPTL3 [El-Shal et al. 2017], CLU (clusterin) [Zheng et al. 2016], FCN2 [Yang et al. 2016], HPX (hemopexin) [Kobayashi et al. 2012], CP (ceruloplasmin) [Pousset et al. 2001] and BCHE (butyrylcholinesterase) [Yokoyama et al. 1982] were involved in invasion of hepatocellular carcinoma cells, but these genes may be associated invasion of hepatoblastoma cells. Enriched genes such as SLC27A2 [Chen et al. 2018] and ABHD2 [Yamanoi et al. 2016] were important for drug resistance in various cancer types, but these genes may be involved in chemo resistance in hepatoblastoma. Our study found that RIDA (reactive intermediate imine deaminase A homolog), ALDH8A1, PFKFB1, PHYH (phytanoyl-CoA 2-hydroxylase), DHTKD1, IYD (iodotyrosinedeiodinase), APOC3, SLC7A2, ART4, CYP2D7, SLC17A3, AVPR1A, INSIG1, GCKR (glucokinase regulator), APOA5, ASPG (asparaginase), BCO2, CFHR1, APCS (amyloid P component, serum), APOL1, FCN3, C8G, PZP (PZP alpha-2-macroglobulin like), ORM1, ASPDH (aspartate dehydrogenase domain containing) and FMO4 are down regulated in hepatoblastoma and has potential as a novel diagnostic and prognostic biomarker, and therapeutic target.

In PPI network and module analysis, we found out the top up regulated hub genes. Over expression of hub genes such as ANLN (anillin actin binding protein) [Zhang et al. 2018], CHEK1 [Yang et al. 2017], IGF2BP3 [Li et al. 2015] and FOXQ1 [Huang et al. 2015] were linked with progression of hepatocellular carcinoma, but these genes may be involved in the advancement of hepatoblastoma. Hub genes such as BMP4 [Deng et al. 2018] and RCN2 [Ding et al. 2017] were responsible for proliferation of hepatocellular carcinoma cells, but this gene may be associated with proliferation of hepatoblastoma cells. Elevated expression of hub genes such as SERPINE2 [Stępień et al. 2017] and KIF11 [Schneider et al. 2017] were liable for progression of thyroid cancer, but this gene may be identified with growth of hepatoblastom. Our study found that TCTN2, PLPPR1, FOXRED2, EFNA4, SORT1, CAD (carbamoyl-phosphate synthetase 2, aspartate transcarbamylase, and dihydroorotase) and MMS22 are up regulated in hepatoblastoma and has potential as a novel diagnostic and prognostic biomarker, and therapeutic target. Similarly, PPI network and module analysis, we found out the top down regulated hub genes. Methylation inactivation tumor suppressor ESR1 was linked with development of hepatocellular carcinoma [Dou et al. 2016], but loss of this gene may be responsible for progression of hepatoblastom. Low expression of hub genes such as AR (androgen receptor) [Shi et al. 2017] and NPY1R [Lv et al. 2016] were liable for progression of hepatocellular carcinoma, but decrease expression of this gene may be identified with growth of hepatoblastom. Hub genes such as ICAM1 [Guo et al. 2016], LPA (lipoprotein(a)) [Malaguarnera et al. 2017], HOOK1 [Sun et al. 2017] and GADD45B [Xia et al. 2017] were responsible for invasion of hepatocellular carcinoma cells, but this gene may be involved in invasion of hepatoblastom cells. Hub genes such as FGGY (FGGY carbohydrate kinase domain containing) [Zhang et al. 2019], USP2 [Davis et al. 2016] and MAP3K5 [Stark et al. 2011] were involved inprogression of various cancer types, but these gene may be involved in development of hepatoblastom. SNP in SLC10A1 was responsible for advancement of hepatocellular carcinoma [Hu et al. 2016], but this polymorphic gene may associated with development of hepatoblastom. Our study found that LRRK2, MYO1B, C8B, RNF125, MACF1 and GLRX (glutaredoxin) are down regulated in hepatoblastoma and has potential as a novel diagnostic and prognostic biomarker, and therapeutic target.

In target genes - miRNA regulatory network analysis, we found out the top up regulated target genes. High expression of target genes such as PLAGL2 [Xu et al. 2018] and SLC1A5 [Toda et al. 2017] were involved in progression of various cancer types, but elevated expression of these genes may be associated with development of hepatoblastom. Our study found that PKM (pyruvate kinase M1/2) is up regulated in hepatoblastoma and has potential as a novel diagnostic and prognostic biomarker, and therapeutic target. Similarly, target genes - miRNA regulatory network analysis, we found out the top down regulated target genes. Decreased expression of target genes such as SOD2 [Wang et al. 2016] and WEE1 [Cuneo et al. 2016] were liable for advancement of hepatocellular carcinoma, but reduced expression these genes may be involved in development of hepatoblastoma. Our study found that APOL6 is down regulated in hepatoblastoma and has potential as a novel diagnostic and prognostic biomarker, and therapeutic target.

In target genes - TF regulatory network analysis, we found out the top up regulated target genes. Methylation inactivation of DCDC2 was responsible for progression of hepatocellular carcinoma [Inokawa et al. 2013], but loss of this gene may be liable for development of hepatoblastoma. Increase expression of DLX5 was important for proliferation of ovarian cancer cells [Tan et al. 2010], but this gene may be associated with proliferation of hepatoblastoma cells. Our study found that WFS1 is up regulated in hepatoblastoma and has potential as a novel diagnostic and prognostic biomarker, and therapeutic target. Similarly, target genes - TF regulatory network analysis, we found out the top down regulated target genes. Our study found that SLC15A1 is down regulated in hepatoblastoma and has potential as a novel diagnostic and prognostic biomarker, and therapeutic target.

Ten candidate genes (TP53, PLK1, AURKA, CDK1, ANLN, ESR1, FGB, ACAT1, GOT1 and ALAS1) that were closely related to the survival rate of patients with hepatoblastoma were diagonised by analyzing the corresponding total survival information from patients with hepatoblastoma from TCGA program. Survival analysis, expression verification, stage analysis, mutation analysis, immunio infiltration analysis and immunio histochemical analysis of these candidate genes were analyzed according to the TCGA database and the human protein atlas database. ROC analysis of these candidate genes were analyzed according to the GSE131329 dataset for prognostic and diagonsitic value of candidate genes. Semelateniously, these ten candidate genes were further verified by RTL using hepatoblastoma patient samples. Overall, the survival characteristics of the ten candidate genes may suggest the prognosis and diagnosis of hepatoblastoma, and thus may be a promising biomarker and therapeutic target for hepatoblastoma.

In summary, the current study used the GEO database, TCGA database and the human protein atlas database to diagonise ten candidate genes as possible biomarkers for patients with hepatoblastoma and documented the existence of the ten candidate genes by RTL CR. The results of the current study provide a potential biomarker for hepatoblastoma screening and diagnosis, therapeutic targets, and prognosis. However, further invedtigations and follow□up of clinical cases are required to expose the molecular pathogenesis and prognosis of hepatoblastoma.

## Data Availability

The datasets supporting the conclusions of this article are available in the GEO (Gene Expression Omnibus) (https://www.ncbi.nlm.nih.gov/geo/) repository. [(GSE131329) (https://www.ncbi.nlm.nih.gov/geo/query/acc.cgi?acc=GSE131329]

## Acknowledgement

I thank Eiso Hiyama, Hiroshima University, Natural Science Center for Basic Research and Deve, LIfe Science, Hiroshima, Japan, very much, the author who deposited their microarray dataset, GSE131329, into the public GEO database.

## Ethical approval

This article does not contain any studies with human participants or animals performed by any of the authors.

## Informed consent

No informed consent because this study does not contain human or animals participants.

## Availability of data and materials

The datasets supporting the conclusions of this article are available in the GEO (Gene Expression Omnibus) (https://www.ncbi.nlm.nih.gov/geo/) repository. [(GSE131329)

(https://www.ncbi.nlm.nih.gov/geo/query/acc.cgi?acc=GSE131329]

## Consent for publication

Not applicable.

## Author Contributions

B. V. - Writing original draft, and review and editing

C. V. - Software and investigation

I. K. - Supervision and resources

